# Impact of COVID-19 pandemic on rates of congenital heart disease procedures among children: Prospective cohort analyses of 26,270 procedures in 17,860 children using CVD-COVID-UK consortium record linkage data

**DOI:** 10.1101/2024.05.20.24307597

**Authors:** Arun Karthikeyan Suseeladevi, Rachel Denholm, Sonya Babu-Narayan, Shubhra Sinha, Serban Stoica, Tim Dong, Gianni Angelini, Cathie Sudlow, Venexia Walker, Katherine Brown, Massimo Caputo, Deborah A Lawlor, the CVD-COVID-UK/COVID-IMPACT Consortium

## Abstract

**Background:** The COVID-19 pandemic necessitated major re-allocation of health care services. Our aim was to assess the impact on paediatric congenital heart disease procedures during different pandemic periods compared to the pre-pandemic period, to inform appropriate responses to future major health services disruptions.

**Methods and Results:** We analysed 26,270 procedures from 17,860 children between 01-Jan-2018 and 31-Mar-2022 in England, linking them to primary/secondary care data. The study period included pre-pandemic and pandemic phases, with latter including three restriction periods and corresponding relaxation periods. We compared procedure characteristics and outcomes between each pandemic periods and the pre-pandemic period. There was a reduction in all procedures across all pandemic period with the largest reductions during the first, most severe restriction period (23-Mar-2020 to 23-Jun-2020), and the relaxation period following second restrictions (03-Dec-2020 to 04-Jan-2021) coinciding with winter pressures. During the first restrictions, median procedures per week dropped by 51 compared with the pre-pandemic period(80 vs 131 per week, p = 4.98×10^-08^). Elective procedures drove these reductions, falling from 96 to 44 per week, (p = 1.89×10^-06^), while urgent (28 vs 27 per week, p = 0.649) and life-saving/emergency procedures (7 vs 6 per week, p = 0.198) remained unchanged. Cardiac surgery rates increased, and catheter-based procedure rates reduced during the pandemic. Procedures for children under 1-year were prioritized, especially during the first four pandemic periods. No evidence was found for differences in post procedure complications (age adjusted odds ratio 1.1 (95%CI: 0.9, 1.4) or post procedure mortality (age and case mix adjusted odds ratio 0.9 (0.6, 1.3)).

**Conclusions:** Prioritization of urgent, emergency and life-saving procedures during the pandemic, particularly in infants, did not impact paediatric CHD post procedure complications or mortality. This information is valuable for future major health services disruptions, though longer-term follow-up of the effects of delaying elective surgery is needed.

## Introduction

The pandemic caused by the novel coronavirus, severe acute respiratory coronavirus type 2 (SARS-CoV-2), necessitated major re-allocation of health care service resources to respond to the need for hospitalization of patients with COVID-19.^1–3^ Children born with congenital heart diseases (CHD) commonly require repeat cardiac catheterization and surgical procedures (hereafter referred to as procedures) across childhood to ensure they maintain healthy cardiac structure and function as they grow.^4–6^ Several studies from different countries including China,^7^ India,^8^ Mexico,^9^ Turkey,^10^ Italy,^11^ and the UK,^12^ have explored the impact of the pandemic on procedures for children with CHD. These have compared the initial period, commonly the first 4 to 6 months, of the pandemic, with a pre-pandemic period, and report marked reductions in elective procedures. These have all been from selected regions or cities, with number of procedures ranging from 29 to ∼8000.^8^ ^10^ None explored effects of varying population restrictions over time, and few examined post procedure complications and mortality.

Learning from the COVID pandemic experiences is crucial for preparing for future disruptions to healthcare services, whether caused by other pandemics, or factors such as extreme weather, wars, or social disruptions like industrial action. Prioritizing services for vulnerable populations during such disruptions is essential, while understanding their consequences is also necessary.

The aim of this study was to assess the impact of the COVID-19 pandemic on paediatric procedures for CHD in England. Specifically, we aimed to describe differences in overall, elective, urgent, emergency and life-saving procedures, and in post procedure complications and mortality during various periods of pandemic restrictions and relaxations compared to the pre-pandemic period. We also explored whether the results varied by the child’s age, neighbourhood deprivation and ethnicity. **Table 1** presents the different phases of pandemic restrictions and relaxations in England.

**Table 1:**
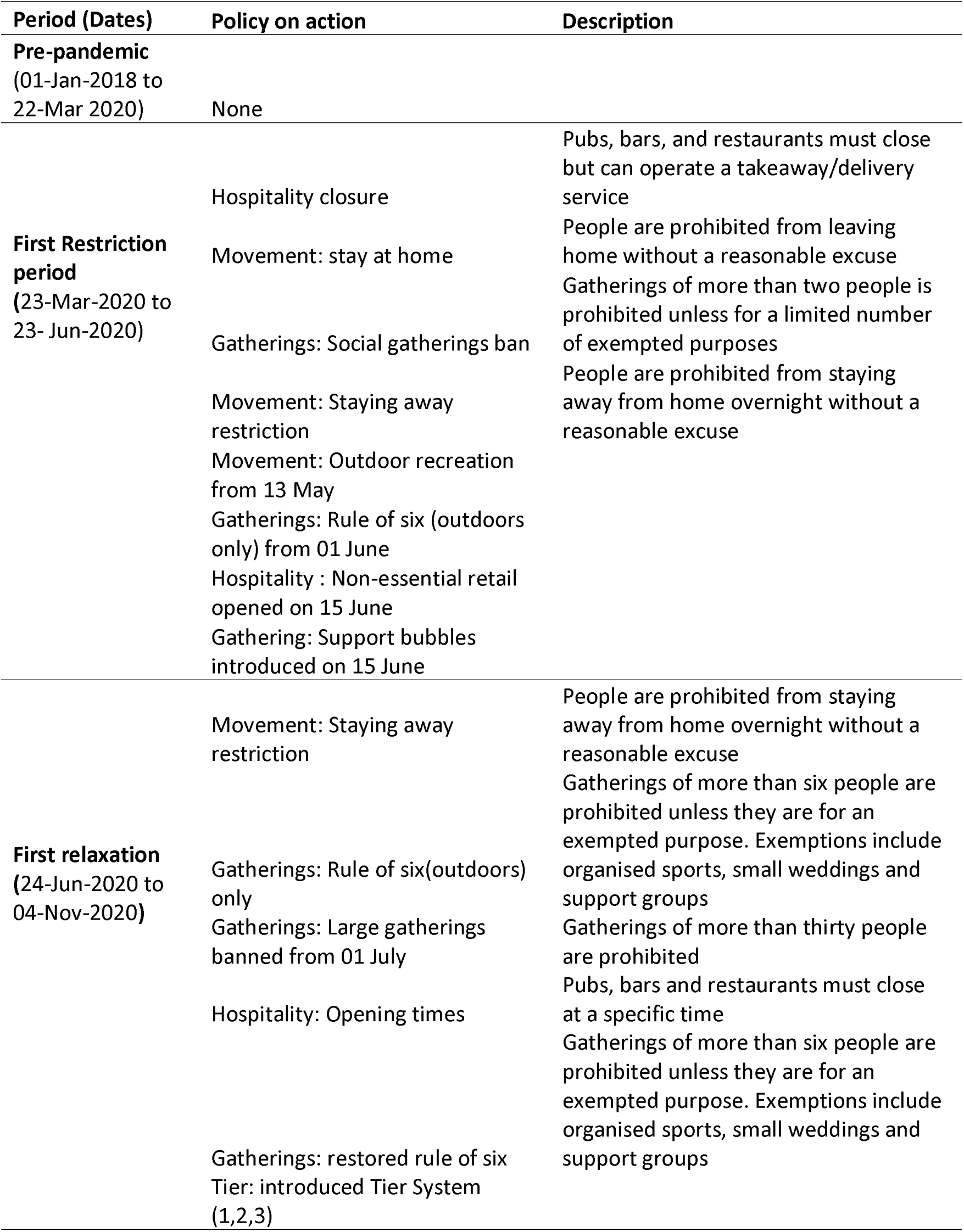
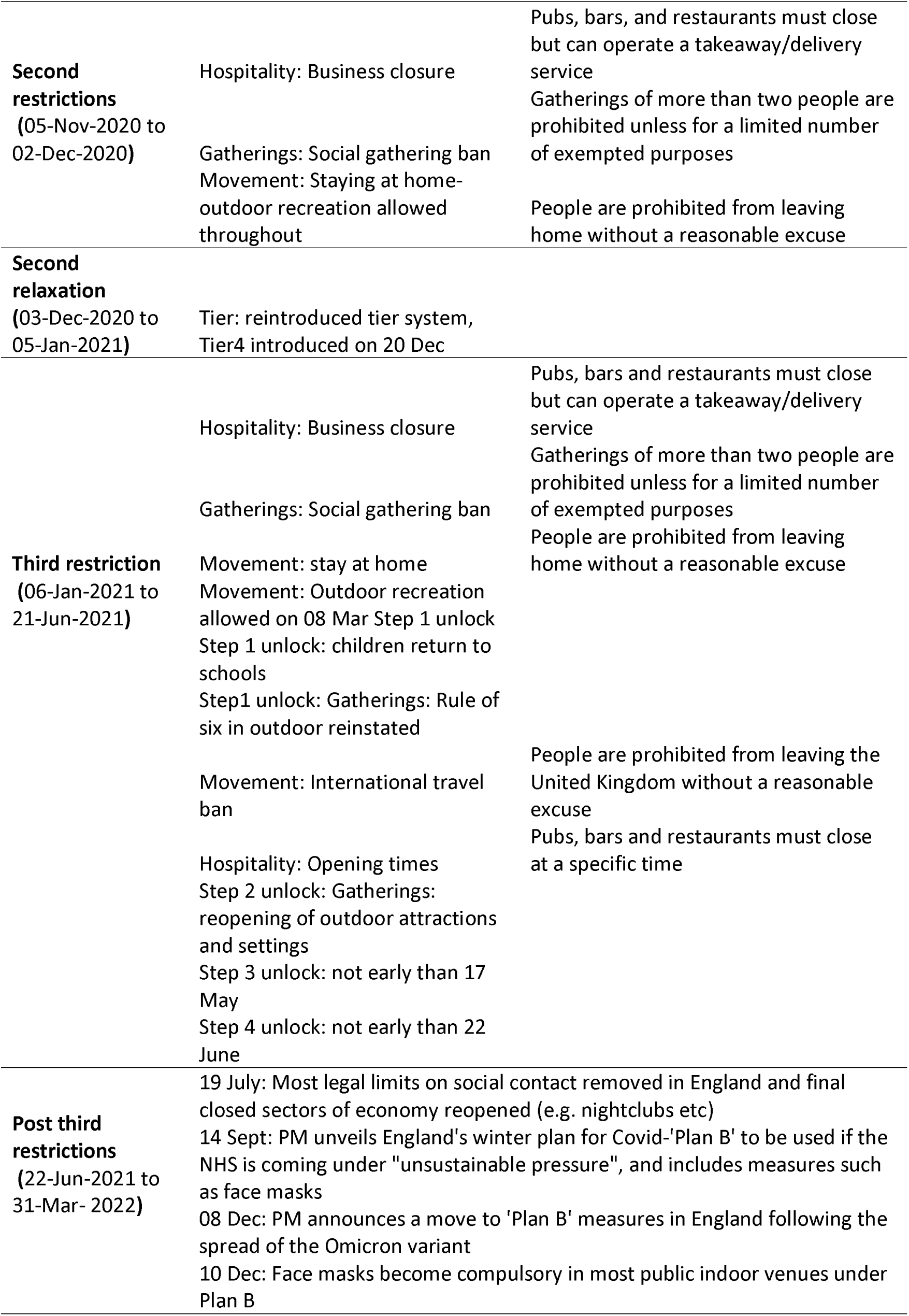
Key restrictions during the different phases of pandemic in England.

## Methods

### Data sources

We used the National Congenital Heart Disease Audit database (NCHDA) as the central dataset. Established in 2000, the NCHDA evaluates outcomes of paediatric and congenital cardiovascular procedures in the UK. Data submission is mandatory for all centres performing these procedures, requiring information on diagnoses, procedures, urgency, and outcomes up to 30-days post procedure.^13^ The NCHDA data undergo validation tests for accuracy and completeness (Supplementary-text).^14–16^

NCHDA data were linked to electronic health records from General Practice Extraction Service (GPES) Data for Pandemic Planning and research (GDPPR), Hospital Episode Statistics (HES), and the Office of National Statistics (ONS) death registry (**Figure 1**; Supplementary-text) Procedures performed between 01-Jan-2018 and 31-Mar-2022, among children under 16 in England were analysed.

**Figure 1:**
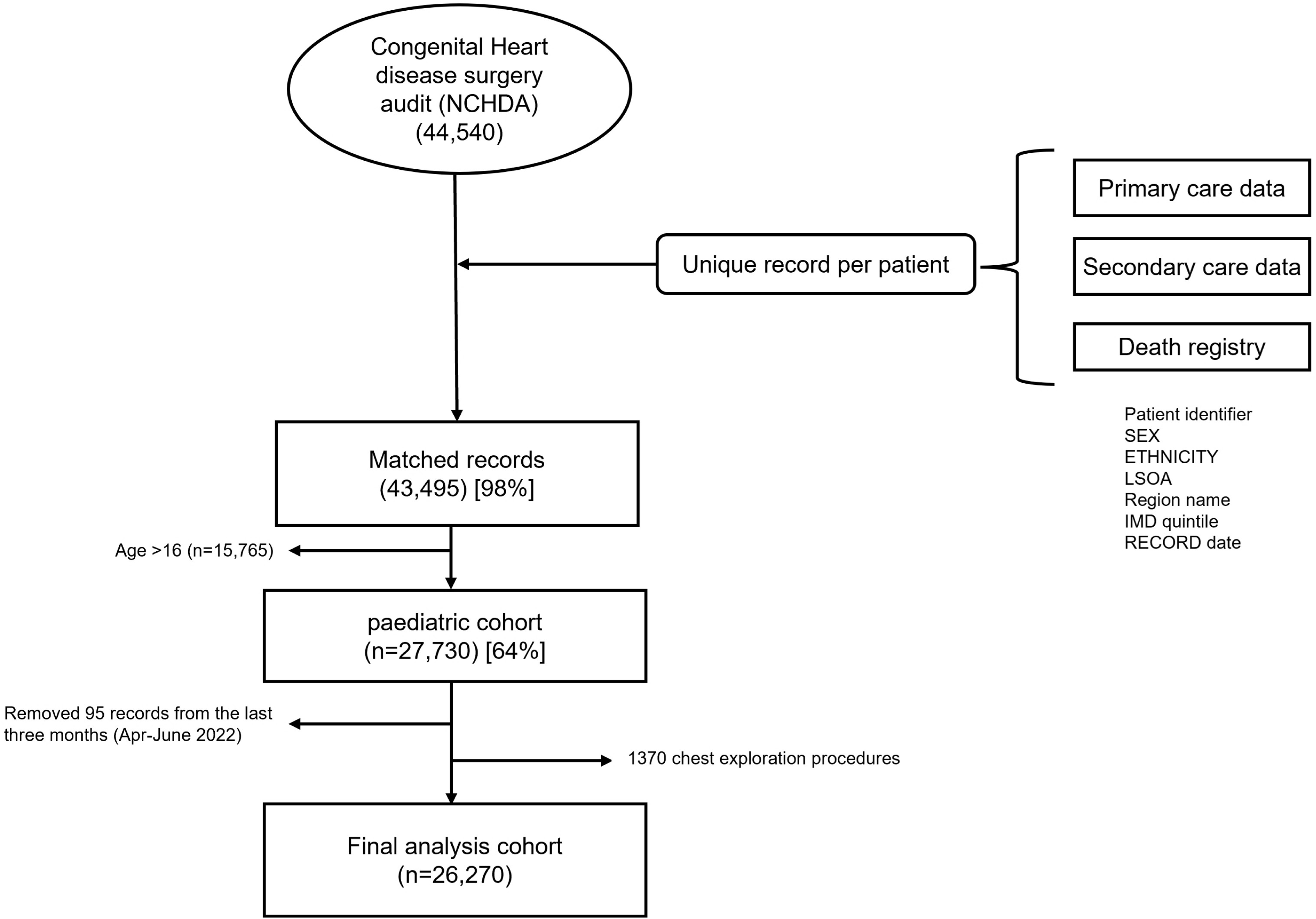
Flow chart of study participants from record linkage to the final analysis sample.

The de-identified data were accessed within NHS England’s Secure Data Environment service^17^ via the BHF Data Science Centre. Ethics and governance details are provided in the **Supplementary-text**.

### Exposure

The exposure time periods reflect the UK’s COVID-19 responses (**Table 1**).^18^

- Pre-pandemic (reference) period (01-Jan-2018 to 22-Mar-2020)
- First restriction period (23-Mar-2020 to 23-Jun-2020)
- First relaxation period (24-Jun-2020 to 04-Nov-2020)
- Second restriction period (05-Nov-2020 to 02-Dec-2020)
- Second relaxation period (03-Dec-2020 to 05-Jan-2021)
- Third restriction period (06-Jan-2021 to 21-Jun-2021)
- Third relaxation period (22-Jun-2021 to 31-Mar-2022).

### Outcomes

The key outcomes were procedure urgency status, post procedure complications and post procedure mortality. Urgency status classifies procedures as elective, urgent, emergency and life-saving. Post procedure complications were defined as any operative or procedure complication occurring within 30 days after the procedure,^13^ ^14^ (full list **Supplementary-Table S1**). Post procedure mortality was defined as deaths within 30-days of the procedure.^13^ ^14^

### Covariates

Mortality following paediatric cardiac surgeries is compared across institutions using the Partial Risk Adjustment in Surgery 2 (PRAIS2) score.^19^ The PRAiS 2 score is estimated using factors such as activity group, specific procedure, primary diagnosis, ventricular physiology, child’s age and weight, and comorbidity, which are specific for cardiac surgeries.^14^ ^20–23^ To adjust for case mix in morbidity for all procedures included in this study, we used individual risk factors. Please refer to the **Supplementary text for** details on the derivation of the variables from the NCHDA dataset and the estimation of PRAiS 2 score, as well as **Supplementary Table S2, S3, and S4** for the full list of primary diagnoses, specific procedures, and risk factors used for adjustment.

### Analyses

The unit of analysis was each procedure, with children undergoing multiple procedures contributing more than once. Robust standard errors were used to account for this. We described the distribution of procedures and children’s sociodemographic characteristics using counts (%), median (interquartile range (IQR)), and mean (standard deviation (SD)).

We present the median (IQR) number of overall, elective, urgent, and emergency/life-saving procedures per week for the pre-pandemic and pandemic periods, using the Wilcoxon rank sum test to compare differences between pandemic periods and the pre-pandemic period. Emergency and life-saving procedures were combined due to low numbers.

We used the Z-test to estimate the difference in mean percentage (95% confidence interval (95% CI) of procedures by (1) urgency: elective, urgent, or emergency/life-saving, (2) type of procedure: cardiac surgery, intervention catheter, or other, and (3) age group: <1-year, 1 to <5-years, 5 to <10-years, 0r 10 to <16-years.

We used age-adjusted logistic regression to estimate the odds ratios for (1) undergoing an urgent, emergency, or life-saving procedure vs elective procedure, (2) post procedure complications (yes vs no), and (3) post procedure mortality within 30-days (yes vs no), comparing each pandemic period to the pre-pandemic period. For the mortality analysis, we additionally adjusted for case mix using PRAIS2 risk factors (**Supplementary-text and Table S4**).

### Sensitivity analyses

We assessed whether using individual PRAIS2 risk factors, rather than the weighted score, influenced our main results by comparing logistic regression outcomes for mortality with three adjustments: age only, age plus individual risk factors, and age plus PRAIS2 score, specifically for cardiac surgeries.

### Exploratory subgroup analyses

We repeated the logistic regression analyses for subgroups based on age, ethnicity, and deprivation quintiles, testing for statistical difference by including interaction terms between these variables and the pandemic periods. The **Supplementary-text** provides justification and details on the characteristics adjusted for in the subgroup analyses.

### Dealing with missing data

No data were missing in the main analysis. Subgroup analyses for ethnicity (missing n = 1,385 (5.3%)) and area deprivation (missing n=1,405 (5.3%)) were limited to complete cases.

This analysis was performed according to a pre-specified analysis plan published on GitHub, along with the phenotyping and analysis code (https://github.com/BHFDSC/CCU007_01).

## Results

The linkage of the NCHDA dataset with routine healthcare data was achieved for 43,495 (98%), with data from primary care, secondary care, or ONS death registry data (91 % linked to primary care, 99% to secondary care, and 90% linked to both sources). After excluding the last low-reporting months (95 records from April to June 2022), and chest closure and exploration procedures (1370 records), the final analysis included 26,270 procedures performed on 17,860 children under 16 years of age, from 01-Jan-2018 to 31-Mar-2022 **(Figure 1** & **Table 2)**.

**Table 2:**
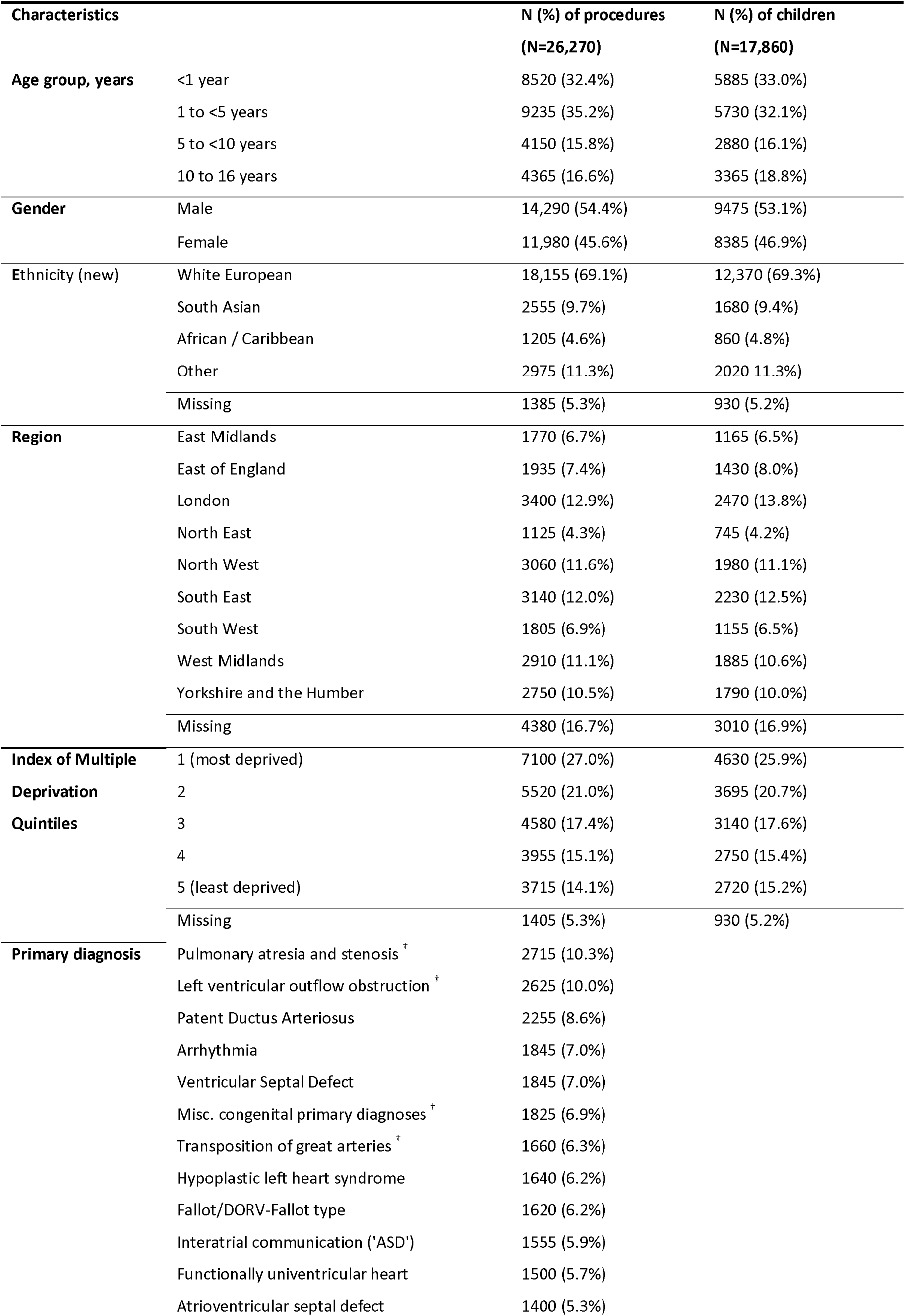
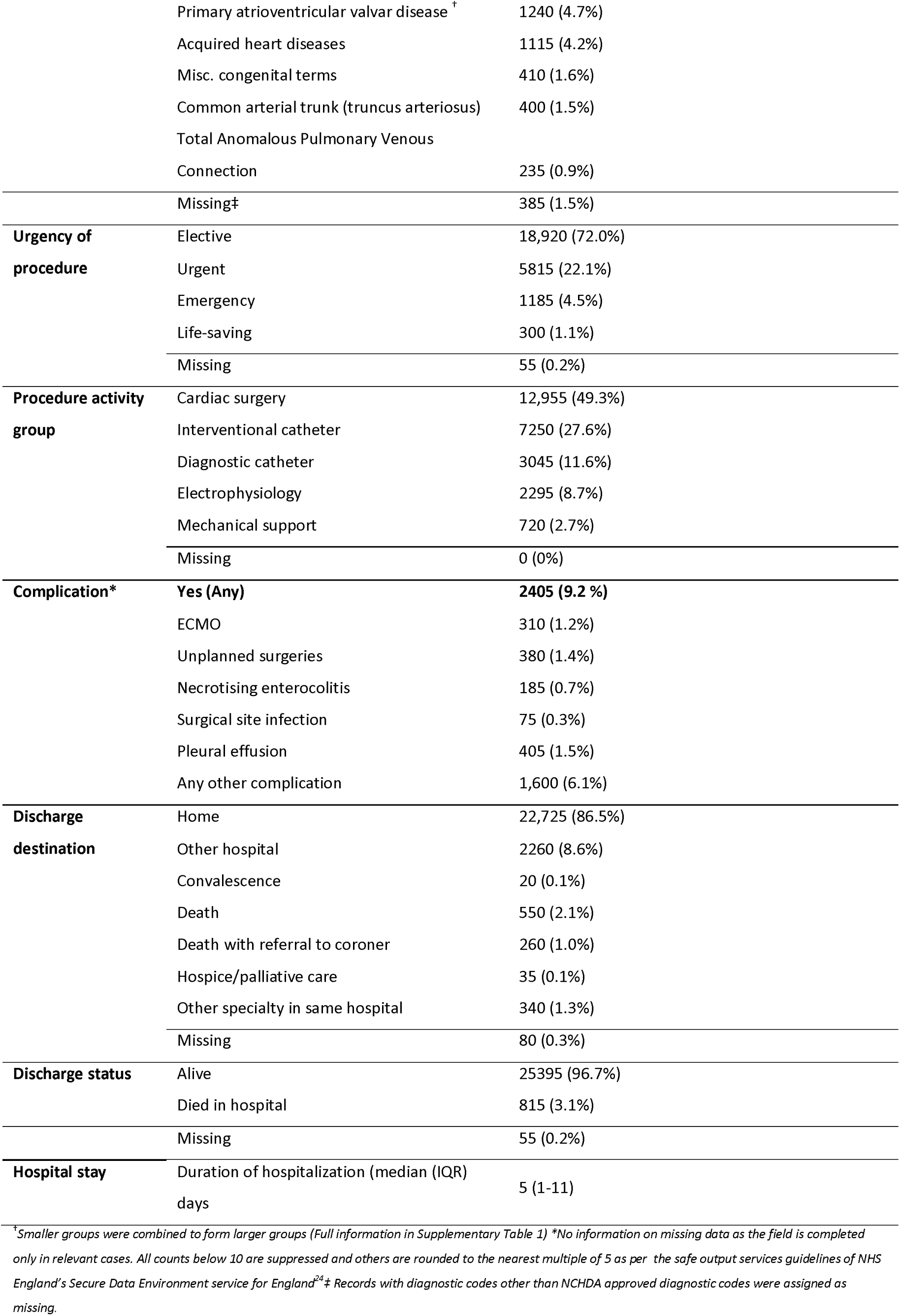
Characteristics of children (< 16-years) who underwent congenital heart disease surgical procedures in England between 01-Jan-2018 and 31-Mar-2022.

| Characteristics |  | N (%) of procedures<br>(N=26,270) | N (%) of children<br>(N=17,860) |
| --- | --- | --- | --- |
| Age group, years | <1 year | 8520 (32.4%) | 5885 (33.0%) |
|  | 1 to <5 years | 9235 (35.2%) | 5730 (32.1%) |
|  | 5 to <10 years | 4150 (15.8%) | 2880 (16.1%) |
|  | 10 to 16 years | 4365 (16.6%) | 3365 (18.8%) |
| Gender | Male | 14,290 (54.4%) | 9475 (53.1%) |
|  | Female | 11,980 (45.6%) | 8385 (46.9%) |
| Ethnicity (new) | White European | 18,155 (69.1%) | 12,370 (69.3%) |
|  | South Asian | 2555 (9.7%) | 1680 (9.4%) |
|  | African / Caribbean | 1205 (4.6%) | 860 (4.8%) |
|  | Other | 2975 (11.3%) | 2020 (11.3%) |
|  | Missing | 1385 (5.3%) | 930 (5.2%) |
| Region | East Midlands | 1770 (6.7%) | 1165 (6.5%) |
|  | East of England | 1935 (7.4%) | 1430 (8.0%) |
|  | London | 3400 (12.9%) | 2470 (13.8%) |
|  | North East | 1125 (4.3%) | 745 (4.2%) |
|  | North West | 3060 (11.6%) | 1980 (11.1%) |
|  | South East | 3140 (12.0%) | 2230 (12.5%) |
|  | South West | 1805 (6.9%) | 1155 (6.5%) |
|  | West Midlands | 2910 (11.1%) | 1885 (10.6%) |
|  | Yorkshire and the Humber | 2750 (10.5%) | 1790 (10.0%) |
|  | Missing | 4380 (16.7%) | 3010 (16.9%) |
| Index of Multiple Deprivation Quintiles | 1 (most deprived) | 7100 (27.0%) | 4630 (25.9%) |
|  | 2 | 5520 (21.0%) | 3695 (20.7%) |
|  | 3 | 4580 (17.4%) | 3140 (17.6%) |
|  | 4 | 3955 (15.1%) | 2750 (15.4%) |
|  | 5 (least deprived) | 3715 (14.1%) | 2720 (15.2%) |
|  | Missing | 1405 (5.3%) | 930 (5.2%) |
| Primary diagnosis | Pulmonary atresia and stenosis <sup>†</sup> | 2715 (10.3%) |  |
|  | Left ventricular outflow obstruction <sup>†</sup> | 2625 (10.0%) |  |
|  | Patent Ductus Arteriosus | 2255 (8.6%) |  |
|  | Arrhythmia | 1845 (7.0%) |  |
|  | Ventricular Septal Defect | 1845 (7.0%) |  |
|  | Misc. congenital primary diagnoses <sup>†</sup> | 1825 (6.9%) |  |
|  | Transposition of great arteries <sup>†</sup> | 1660 (6.3%) |  |
|  | Hypoplastic left heart syndrome | 1640 (6.2%) |  |
|  | Fallot/DORV-Fallot type | 1620 (6.2%) |  |
|  | Interatrial communication ('ASD') | 1555 (5.9%) |  |
|  | Functionally univentricular heart | 1500 (5.7%) |  |
|  | Atrioventricular septal defect | 1400 (5.3%) |  |

|  |  |  |
| --- | --- | --- |
|  | Primary atrioventricular valvar disease <sup>†</sup> | 1240 (4.7%) |
|  | Acquired heart diseases | 1115 (4.2%) |
|  | Misc. congenital terms | 410 (1.6%) |
|  | Common arterial trunk (truncus arteriosus) | 400 (1.5%) |
|  | Total Anomalous Pulmonary Venous Connection | 235 (0.9%) |
|  | Missing‡ | 385 (1.5%) |
| **Urgency of procedure** | Elective | 18,920 (72.0%) |
|  | Urgent | 5815 (22.1%) |
|  | Emergency | 1185 (4.5%) |
|  | Life-saving | 300 (1.1%) |
|  | Missing | 55 (0.2%) |
| **Procedure activity group** | Cardiac surgery | 12,955 (49.3%) |
|  | Interventional catheter | 7250 (27.6%) |
|  | Diagnostic catheter | 3045 (11.6%) |
|  | Electrophysiology | 2295 (8.7%) |
|  | Mechanical support | 720 (2.7%) |
|  | Missing | 0 (0%) |
| **Complication\*** | **Yes (Any)** | **2405 (9.2 %)** |
|  | ECMO | 310 (1.2%) |
|  | Unplanned surgeries | 380 (1.4%) |
|  | Necrotising enterocolitis | 185 (0.7%) |
|  | Surgical site infection | 75 (0.3%) |
|  | Pleural effusion | 405 (1.5%) |
|  | Any other complication | 1,600 (6.1%) |
| **Discharge destination** | Home | 22,725 (86.5%) |
|  | Other hospital | 2260 (8.6%) |
|  | Convalescence | 20 (0.1%) |
|  | Death | 550 (2.1%) |
|  | Death with referral to coroner | 260 (1.0%) |
|  | Hospice/palliative care | 35 (0.1%) |
|  | Other specialty in same hospital | 340 (1.3%) |
|  | Missing | 80 (0.3%) |
| **Discharge status** | Alive | 25395 (96.7%) |
|  | Died in hospital | 815 (3.1%) |
|  | Missing | 55 (0.2%) |
| **Hospital stay** | Duration of hospitalization (median (IQR) days | 5 (1-11) |

<sup>†</sup>Smaller groups were combined to form larger groups (Full information in Supplementary Table 1) \*No information on missing data as the field is completed only in relevant cases. All counts below 10 are suppressed and others are rounded to the nearest multiple of 5 as per the safe output services guidelines of NHS England's Secure Data Environment service for England<sup>24</sup> ‡ Records with diagnostic codes other than NCHDA approved diagnostic codes were assigned as missing.

**Table 2** presents the distributions of sociodemographic and clinical characteristics for all procedures throughout the analysis period. The predominant ethnic group was White European, and the London region had the highest proportion of cases. Pulmonary atresia and stenosis and left ventricular outflow obstruction were the most common primary diagnosis, while total anomalous pulmonary venous connection was the least common. Of all the procedures, 72% (n=18,920) were elective, with the most being cardiac surgeries (n=12,955, 49%) or intervention catheters (n=7250, 28%). During the study period, post procedure complication were below 10% and 2.1% children died within 30 days.

There was a reduction in median (IQR) number of overall procedures per week across all pandemic periods in comparison to the pre-pandemic period. (**Figure 2**) The largest declines occurred during the first and most severe pandemic restriction and the relaxation following the second restriction, while the smallest differences were during relaxations following the first and second restriction periods. Elective procedures drove these reductions in elective procedures, decreasing from 96 per week to 44 per week during the first restriction (p = 1.89×10^-06^). Urgent procedures showed no change (27 vs 28 per week, p = 0.649) nor did life-saving/emergency procedures (6 vs 7 per week, p = 0.198). Differences in mean percentage of urgent and emergency/life-saving procedures between pandemic and pre-pandemic period followed similar patterns **(Supplementary Figure S1)**.

**Figure 2:**
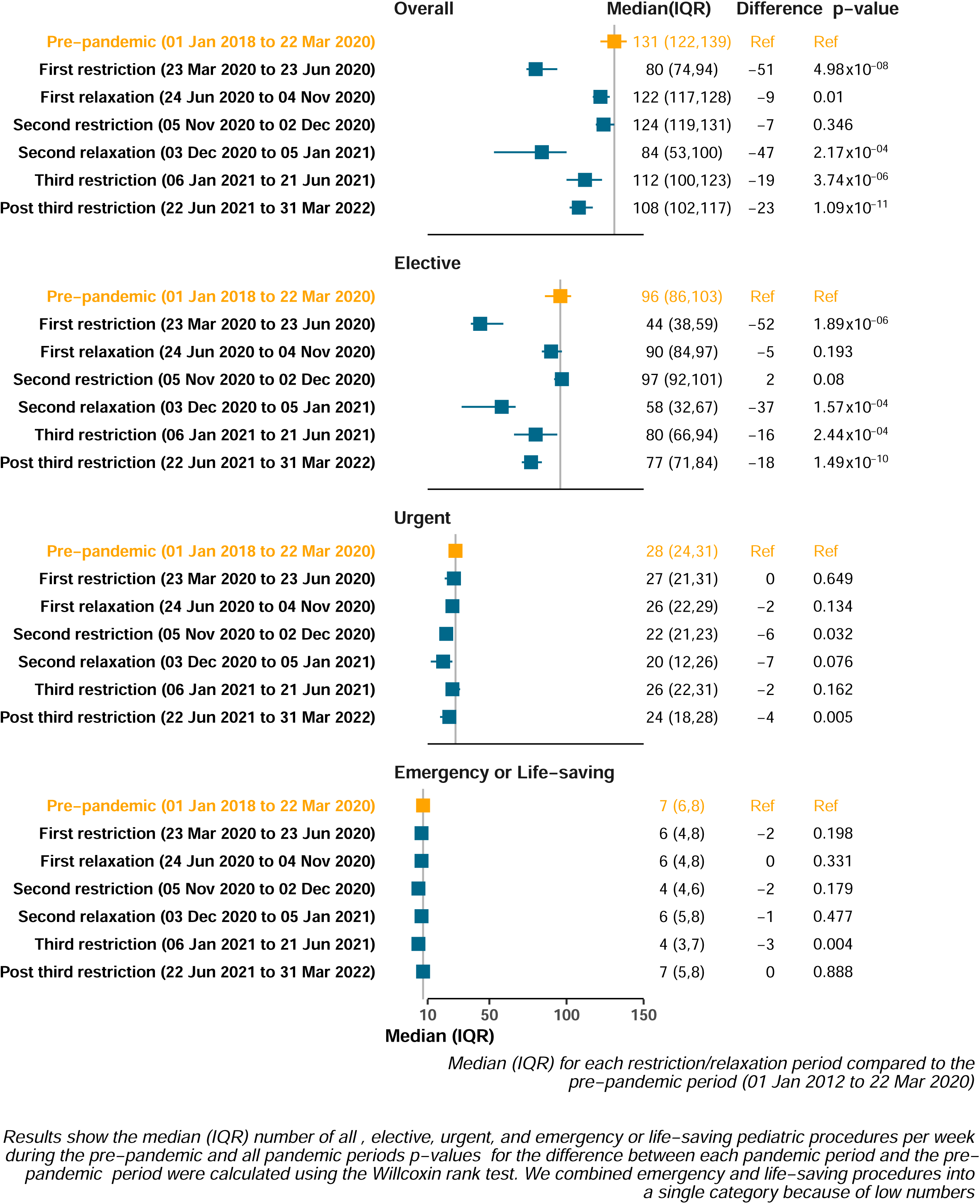
Difference in weekly median numbers of all, elective, urgent and emergency/life−saving pediatric congenital heart disease procedures comparing pandemic periods to the pre−pandemic period.

**Figure 3** illustrates difference in mean percentage of procedure types performed during pandemic periods compared to the pre-pandemic period. During the first restriction, there was a 6.1% [95%CI: 3.1, 9.1] increase in cardiac surgeries, accompanied by reduction in catheter (-2.8 % [-5.4, -0.2]) and other (-3.3% [-5.7, 0.9]) procedures. This was followed by a gradual return towards pre-pandemic levels until the final pandemic periods, with reduction in cardiac surgeries and increase in catheter procedures compared to pre-pandemic levels. **Supplementary table S7** details the differences in mean percentages for each specific procedure. Among the 86 specific procedures, 36 were less likely, 46 were more likely and 6 showed no difference between the first restriction period and pre-pandemic levels. Procedures that were less likely included electrophysiological ablation, atrial septal defect, atrial septal defect transluminal, total cavo-pulmonary connection (known as Fontan’s procedure), and patent ductus arteriosus transluminal, while those that were more likely included Fallot’s, balloon atrial septostomy, coarctation hypoplasia, superior vena cava to pulmonary artery anastomosis (known as Glenn’s anastomosis). There was no strong evidence of differences in specific procedures during other pandemic periods, though we had limited power at this granular level.

**Figure 3:**
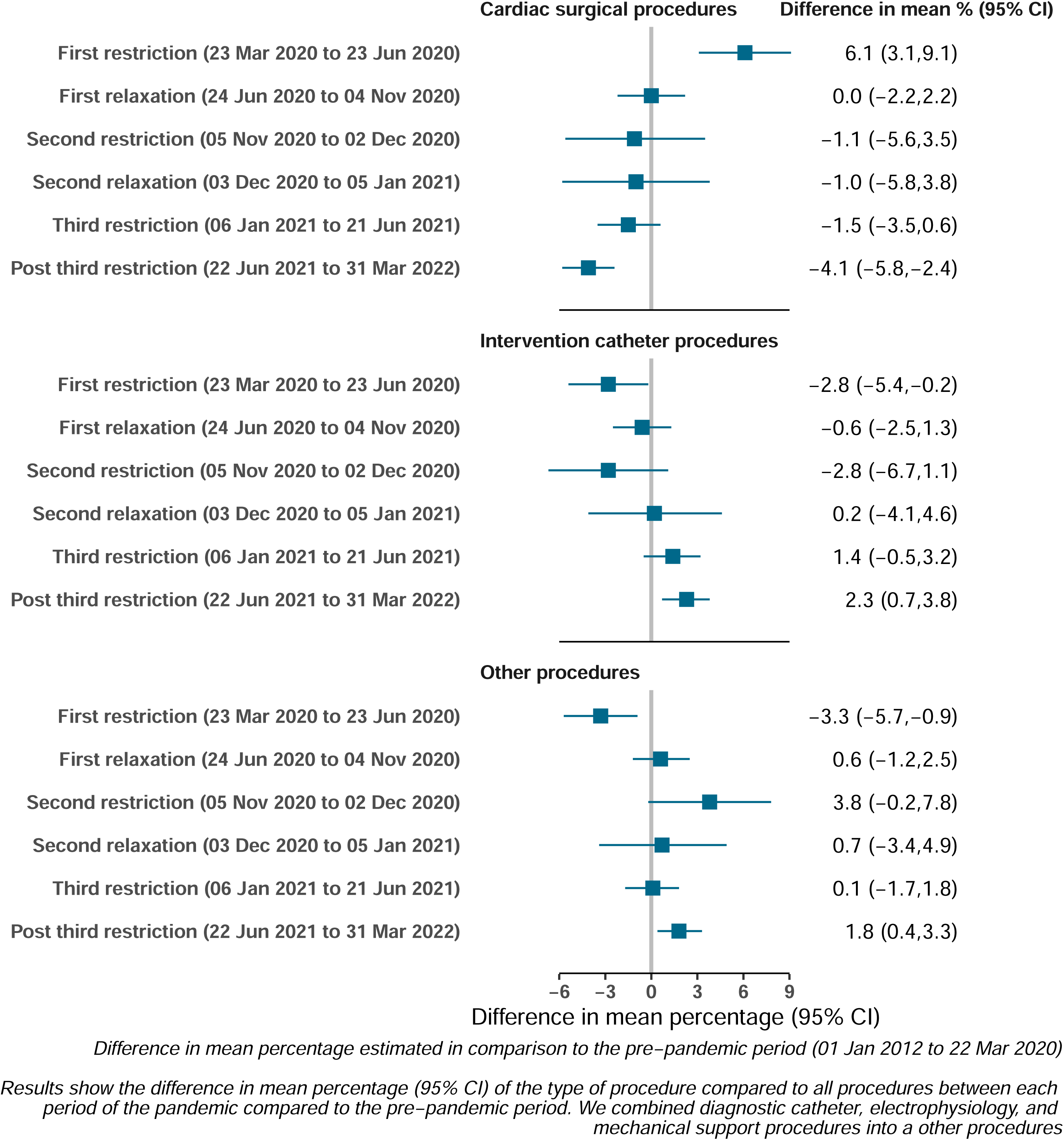
Difference in the mean percentage of each procedure during pandemic periods compared to the pre−pandemic.

Across all pandemic periods, except the third restriction and post-pandemic period, procedures among children under 1-year were higher than pre-pandemic levels (**Figure 4**). In the third restriction period, procedures in this age group were lower than in the pre-pandemic period. For other age groups, patterns varied across the pandemic periods. By the post-pandemic period, procedures for children aged 1 to below 5 were lower than the pre-pandemic period, while the other three age groups remained similar to pre-pandemic period.

**Figure 4:**
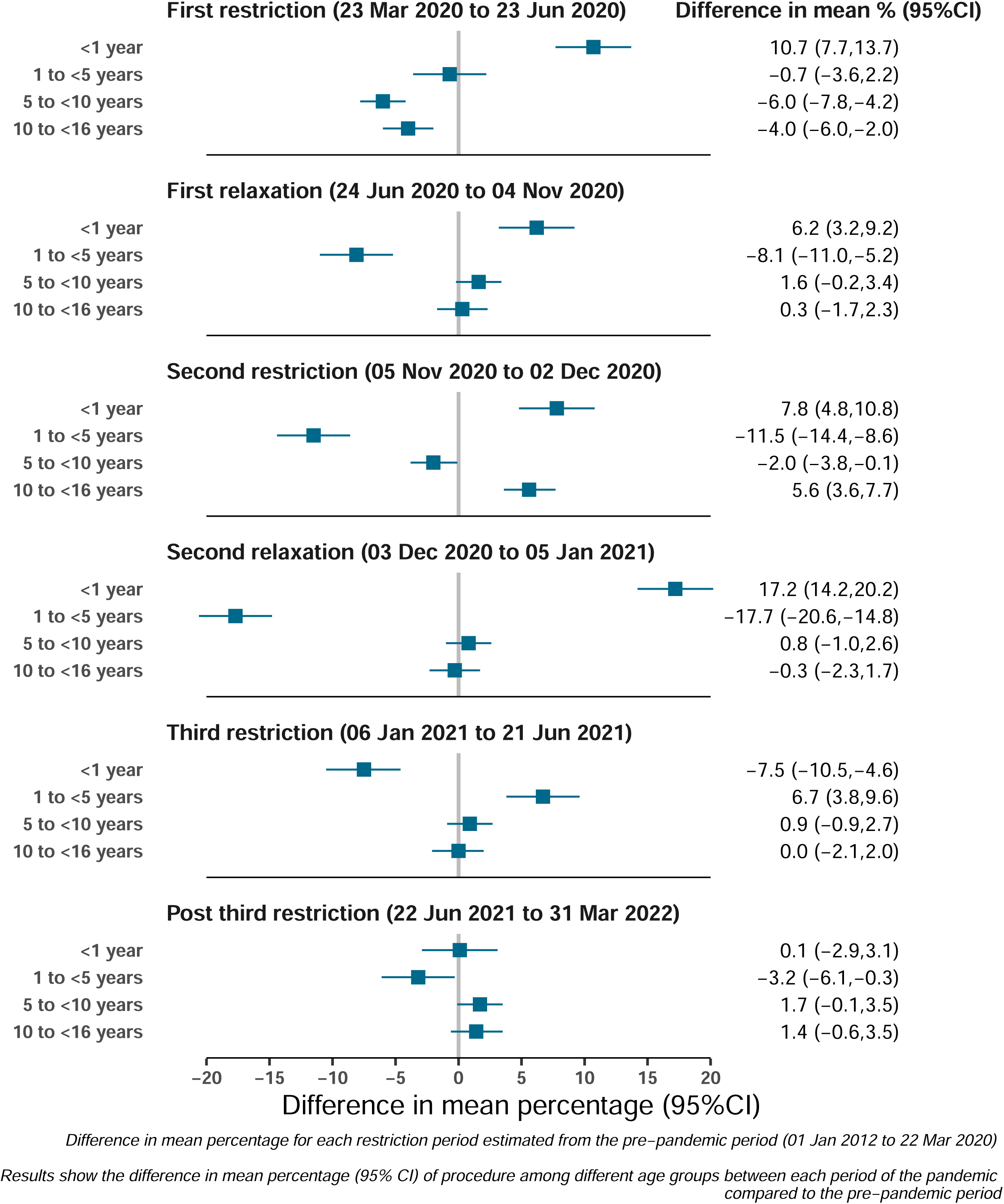
Difference in the mean percentage of the age group of procedure during pandemic periods compared to the pre−pandemic.

There was a marked increase in odds of urgent, emergency, or life-saving procedures in the first period of restrictions (age adjusted odds ratio 1.6 [95%CI: 1.4, 1.8]), followed by a reduction in the subsequent relaxation period (age adjusted odds ratio 0.8 [95%CI: 0.7, 1.9]). We did not find evidence of differences in post procedure complications or post procedure mortality within 30-days during any pandemic period, compared with pre-pandemic (**Figure 5**).

**Figure 5:**
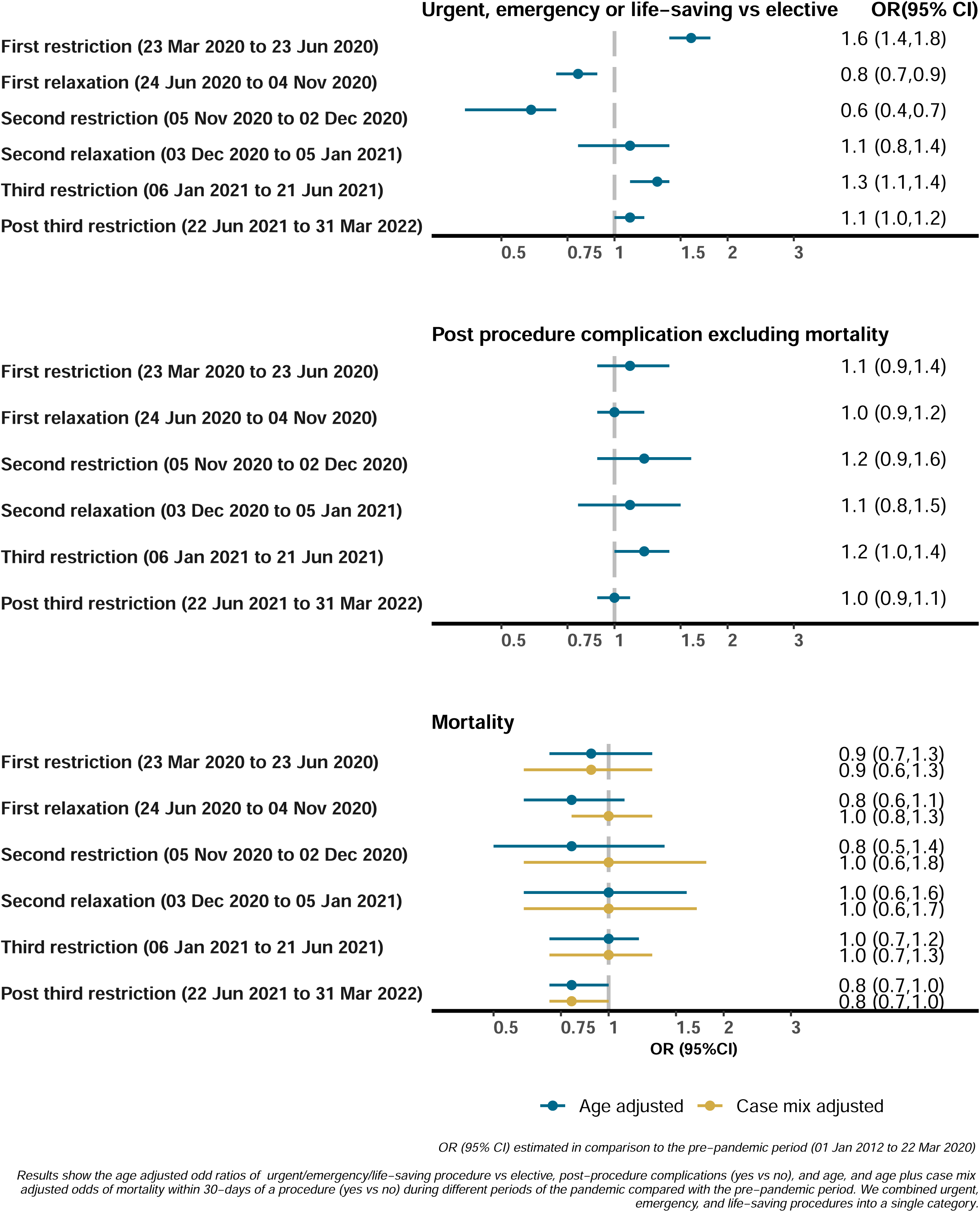
Odds ratios of urgency, post−procedure complications, and mortality within 30−days of a procedure comparing pandemic periods to the pre−pandemic period.

In sensitivity analysis, we found no difference in the odds of mortality within 30 days, compared to pre-pandemic, in age adjusted, age plus individual case mix risk factor adjusted, and age plus PRAIS2 risk score adjusted models **(Supplementary-Figure S2)**.

Exploratory subgroup analyses revealed statistical differences in the association of pandemic periods with the procedure urgency across age groups (interaction p-value = 2.96×10^-09^; **Supplementary Figure S3**). The odds of urgency increased during the first restriction period for all age groups, especially for older children, but this effect diminished in the subsequent periods. There was no evidence of differences by age group for the odds of post procedure complications (interaction p-value 0.09; **Supplementary Figure S4**) and imprecise estimates with wide confidence intervals precluded strong conclusions about mortality (interaction p-value 0.007; **Supplementary Figure S5**).

Ethnic group analysis showed increased odds for urgency except for South Asian children during the first restriction period, with patterns returning to pre-pandemic rate by the third relaxation period (interaction p-value 0.003, **Supplementary Figure S6**). There were no differences in the association of pandemic periods with complications by ethnicity (interaction p-value 0.580; **Supplementary Figure S7**). Although there was statistical evidence of differences in post-procedure mortality between ethnic groups (interaction p-value = 4.7×10^-05^, **Supplementary Figure S8**), estimates were too imprecise for meaningful conclusions.

Analysis by residential area deprivation showed no evidence that associations of pandemic periods with urgent, emergency, or life-saving procedures (interaction p-value 0.744; **Supplementary Figure S9**) or complications (interaction p-value 0.6367; **Supplementary Figure S10**). Estimates for mortality were too imprecise for robust conclusions (interaction p-value = 1.03×10^-05^; **Supplementary Figure S11**).

## Discussion

This study is, to our knowledge, the largest study using whole population data to examine the impact of the COVID-19 pandemic response on CHD procedures in children. We found that the median number of CHD procedures per week was lower during all pandemic periods compared to pre-pandemic levels. The largest reductions occurred during the first, most severe restrictions, and the relaxation period following the second restrictions, coinciding with winter pressures. These reductions were primarily driven by reductions in elective procedures, while urgent and emergency/life-saving procedures remained stable compared to pre-pandemic rates. There was evidence of prioritizing cardiac surgery over catheterization and prioritizing infants during the pandemic. Reassuringly, we found limited evidence of increased post procedure complications or mortality during the pandemic compared to the pre-pandemic levels.

Children with complex CHD require repeat procedures and/or percutaneous/hybrid interventions throughout their lives.^5^ ^6^ Some conditions, such as transposition of the great arteries, hypoplastic left heart syndrome, are time sensitive and require immediate perinatal attention. The prioritization of urgent, emergency and life-saving CHD procedures over elective ones, as seen in our and other studies.^2^ ^3^ ^25^, may explain why we observed no differences in post procedure complications or mortality within 30-days. However, the impact of delays in elective surgery and the broader effects of major disruptions to specialized surgery care during the pandemic - such as resource reallocation, staff fatigue, illness, and family anxiety - remain unknown. Continuing this study over a longer period will allow us to explore the pandemic’s impact on children’s cardiovascular and overall health. New linkages to educational administrative datasets and family members health care records will facilitate investigations into effects on children’s educational outcomes and the mental health of children, parents and other family members.

We explored whether the associations we observed differed by the child’s age, ethnicity, and residential area deprivation and found statistical evidence for some. The increased odds of urgent, emergency, or life-saving procedures in older children, during the first restriction period and other pandemic periods likely reflects the prioritization of procedures in younger children. This indicates that infants were more likely to have elective, urgent, emergency or life-saving procedures, compared with older children. However, we acknowledge that our subgroup analyses were under-powered and like all subgroup analyses, require replication.

### Strengths and limitations

A key strength of this study is the use of country-wide data for all the CHD procedures performed in England. This is made possible by the mandatory requirement for all institutions conducting paediatric cardiac procedures to submit complete data to NCHDA. We linked this data to primary and secondary care records to conduct our analyses. The NCHDA ensures high accuracy through rigorous validation processes, including complication and mortality verification. With a Data Quality Index (DQI) score >90% considered good, all paediatric centres met this standard in the recent audit report.^16^ ^26^ Furthermore, for our main analyses, there were no missing data. To our knowledge, this is the largest study to-date, allowing us to examine how healthcare provision for paediatric CHD procedures changed over an extended period of varying restrictions. While the large numbers enabled exploratory subgroup analyses, we recognize that even with substantial data, estimates remain imprecise, and larger studies would be necessary for more robust conclusions. There were small amounts of missing data for ethnicity and residential area deprivation (5.3% each), which could bias results if concentrated in specific subgroups. This is not possible to explore. However, since these data come from electronic health records and the missing proportion is small, we suspect any bias would be minimal.

Between-hospital variation in the timing of mandatory data uploads can lead to incomplete data or artificial trends towards the most recent months of analysis. To mitigate this, we initially extracted data until 30-Jun-2022 but excluded the last three months, including data up to 31-Mar-2022.

Our analysis operates at a population level limiting our ability to map individual patient experiences or quantify differences in delays, particularly regarding the impact of elective surgery delays. We categorized the pandemic months into six periods of restrictions and relaxation; however, these restrictions were not uniformly applied (see **Table 1**). For instance, the first period was the most stringent and consistent nationwide, while the second involved some regional variations in restrictions, and the third included six gradual steps of easing measures until the pandemic was declared over. We a priori decided to analyse each period of any restrictions in the same way to increase power to detect differences, including for the rarer outcomes of post procedure complications and post procedure mortality. Thus, our results cannot be interpreted as potential effects of specific restrictions, rather they illustrate the broader impact of health services pressures that necessitate delaying elective procedures and prioritizing more urgent cases.

### Implications and conclusions

Our results suggest that when pressures on health services result in prioritization of urgent, emergency and life-saving procedures in children with CHD and delaying elective procedures, this does not result in increased post procedure complications or mortality, over a period of two years. These findings have implications for future health service provision, particularly during infectious disease epidemics or global pandemics as well as during extreme weather events common across Europe.^27–29^ Notably, during the relaxation period following the second restriction, the median rates of overall and elective procedures dropped to levels comparable to those in the first restriction period, exceeding the reductions seen during the second restriction. This second relaxation occurred during winter (03-Dec-2020 to 05-Jan-2021) and may reflect winter pressures. As climate change intensifies the frequency of weather extremes, such pressures are likely to rise, highlighting the need for strategies to mitigate climate change and effective plans to manage health services pressures on from various sources.

In conclusion, our findings suggest that delaying elective procedures in children with CHD to prioritize urgent, emergency and life-saving procedures does not increase procedure-related complications or 30-day mortality, making this approach appropriate in times of healthcare pressures. However, further research is essential to assess the long-term effects of such delays on cardiovascular health of children and the mental health and wellbeing of affected children, their parents and family members.

## Data Availability

The data used in this study are available in NHS England Secure Data Environment service for England, but as restrictions apply they are not publicly available. Those wishing to gain access to the data should contact British Heart Foundation Data Science Centre in the first instance.

https://digital.nhs.uk/services/secure-data-environment-service

## Acknowledgements

This work is carried out with the support of the BHF Data Science Centre led by HDR UK (BHF Grant no. SP/19/3/34678). This study makes use of de-identified data held in NHS England’s Secure Data Environment service for England and made available via the BHF Data Science Centre’s CVD-COVID-UK/COVID-IMPACT consortium. This work uses data provided by patients and collected by the NHS as part of their care and support. We would also like to acknowledge all data providers who make health relevant data available for research.

## Competing interests

All authors declare they have no conflict of interest.

## Sources of Funding

The British Heart Foundation Data Science Centre (grant No SP/19/3/34678, awarded to Health Data Research (HDR) UK) funded co-development (with NHS England) of the Secure Data Environment service for England, provision of linked datasets, data access, user software licences, computational usage, and data management and wrangling support, with additional contributions from the HDR UK Data and Connectivity component of the UK Government Chief Scientific Adviser’s National Core Studies program to coordinate national COVID-19 priority research. Consortium partner organizations funded the time of contributing data analysts, biostatisticians, epidemiologists, and clinicians. This work was also supported by the British Heart Foundation Accelerator Award to the University of Bristol (AA/18/1/34219) and Bristol’s National Institute for Health Research Biomedical Research Centre. GA’s (R104281-102), MC’s (CH/17/1/32804) and DAL’s (CH/F/20/90003) contributions are supported by their British Heart Foundation Chairs. AKS’s and DAL’s contribution is further supported by DAL’s National Institute for Health Research Senior Investigator Award ((NF-0616-10102). AKS, VW, and DAL work in, or are affiliated with, the UK Medical Research Council Integrative Epidemiology Unit (MC_UU_00032/03 and MC_UU_00032/05). RD and VW received support from the Longitudinal Health and Wellbeing COVID-19 National Core Study, which was established by the UK Chief Scientific Officer in October 2020 and funded by UK Research and Innovation (grant references MC_PC_20030 and MC_PC_20059) and from the CONVALESCENCE study of long COVID-19 funded by National Institute for Health and Care Research (NIHR)/UK Research and Innovation.

## Author contributions

MC and DAL came up with the idea of the study, AKS and DAL wrote the initial protocol/analysis plan with RD, TD, GA, KB, MC contributing to subsequent development. AKS, RD, CS, VW, S B-N, KB contributed to obtaining and linking data and quality control. AKS completed analyses with support from RD and DAL. AKS and DAL wrote the first draft of the paper, with all other co-authors making critical comments and revisions. CS is the Director of the BHF Data Science Centre and coordinated approvals for and access to data within NHS England’s Secure Data Environment service for England, for the CVD-COVID-UK/COVID-IMPACT research programme.

## Data availability

The data used in this study are available in NHS England’s Secure Data Environment (SDE) service for England, but as restrictions apply they are not publicly available (https://digital.nhs.uk/services/secure-data-environment-service). The CVD-COVID-UK/COVID-IMPACT programme, led by the BHF Data Science Centre (https://bhfdatasciencecentre.org/), received approval to access data in NHS England’s SDE service for England from the Independent Group Advising on the Release of Data (IGARD) (https://digital.nhs.uk/about-nhs-digital/corporate-information-and-documents/independent-group-advising-on-the-release-of-data) via an application made in the Data Access Request Service (DARS) Online system (ref. DARS-NIC-381078-Y9C5K) (https://digital.nhs.uk/services/data-access-request-service-dars/dars-products-and-services). The CVD-COVID-UK/COVID-IMPACT Approvals & Oversight Board (https://bhfdatasciencecentre.org/areas/cvd-covid-uk-covid-impact/) subsequently granted approval to this project to access the data within NHS England’s SDE service for England. The de-identified data used in this study were made available to accredited researchers only. Those wishing to gain access to the data should contact in the first instance.

The North East - Newcastle and North Tyneside 2 research ethics committee provided ethical approval for the CVD-COVID-UK/COVID-IMPACT research programme (REC No 20/NE/0161) to access, within secure trusted research environments, unconsented, whole-population, de-identified data from electronic health records collected as part of patients’ routine healthcare.

Patient and Public Involvement: Patients and the public were not involved in the design, conduct, reporting, or dissemination plans of this research

## Notes

### Competing Interest Statement

The authors have declared no competing interest.

### Clinical Protocols

https://github.com/BHFDSC/CCU007_01/Protocol

### Author Declarations

The North East - Newcastle and North Tyneside 2 research ethics committee provided ethical approval for the CVD-COVID-UK/COVID-IMPACT research programme (REC No 20/NE/0161)

### Summary of Updates

Reduced word count for the main text. Moved additional methods to Supplementary text. Supplementary files updated

