## Supplementary materials for "Impact of COVID-19 pandemic on rates of congenital heart disease procedures among children: Prospective cohort analyses of 26,270 procedures in 17,860 children using CVD-COVID-UK consortium record linkage data"

We used the National Congenital Heart Disease Audit database (NCHDA) as the central dataset to which other datasets were linked in order to create the cohort used in this study. Established in 2000 as the Central Cardiac Audit Database (congenital), the NCHDA evaluates outcomes of pediatric and congenital cardiovascular procedures, including surgery, transcatheter and electrophysiological interventions, in the UK. Data submission is mandatory for all centers performing such procedures. The dataset contains information on the diagnoses, procedure, procedure urgency, discharge status, and outcomes (complications and mortality) up to 30-days after the procedure.^15^ The NCHDA data undergoes a series of validation tests, including annual site visits by an independent clinical data auditor and volunteer clinician to ensure full case ascertainment and to validate the accuracy of the data submitted.^16-18^ In addition, a random selection of patients records at each site undergo detailed analysis of all submitted data fields, comparing the dataset and the hospital records, for missing or incorrect data.

We linked the patients in the NCHDA dataset to their routine electronic health records available in General Practice Extraction Service (GPES) Data for Pandemic Planning and research (GDPPR), Hospital Episode Statistics (HES), or Office of National Statistics (ONS) death registry. These datasets provided information on date of birth, date of death, and socio-demographic data, such as neighborhood deprivation and ethnicity. We limited our analysis to procedures performed between 01^st^ January 2018 to 31^st^ March 2022 among children below 16 years of age residing in England. **Figure 1** summarizes the linkage and numbers of children/procedures included in this study.

The de-identified data were accessed within National Health Service (NHS) England’s privacy-protecting Secure Data Environment service for England^19^, made available via the British Heart Foundation (BHF) Data Science Centre’s [CVD-COVID-UK/COVID-IMPACT Consortium](https://bhfdatasciencecentre.org/areas/cvd-covid-uk-covid-impact/). The CVD-COVID-UK/COVID-IMPACT Approvals and Oversight Board provided ethical approval and oversight of ethics and governance for all analyses in the consortia. NHS England’s disclosure control rules were in place to prevent disclosure of personal, sensitive, and confidential data. This includes suppressing counts based on fewer than 10 participants and rounding counts to the nearest multiple of five where there are more than 10 participants.^20^

*Covariates:*

We calculated the age of the child on the procedure date and categorized them into four age categories: <1 year, 1 to < 5 years, 5 to < 10 years, and 10 to 16 years for adjustment and exploratory subgroup analyses. These groups reflect clinical practice. For instance, most children with CHD diagnosed early in life need at least one procedure before 1-year, and numbers of procedures differ by age.

CHD case mix

Each record in the NCHDA dataset corresponds to a unique procedure (i.e., there will be multiple records for anyone who has had more than one procedure). The NCHDA database uses the Association for European Pediatric and Congenital Cardiology (EPCC) derived version of the International Pediatric and Congenital Cardiac Code (IPCCC).^22^ A combination of up to eight individual procedure codes describe each operation.^22,23^ We used the activity analysis algorithm (version 6.14), which employs a hierarchical method to aggregate individual EPCC codes from patient procedure records into distinct activity groups: cardiac surgery, interventional catheter, diagnostic catheter, electrophysiological, mechanical support, and chest closure and exploration. ^16^ Procedures belonging to the chest closure and exploration group were excluded from this analysis as these were deemed as part of the main operation. (**Figure 1**)

We used the specific procedure algorithm, developed by NCHDA steering committee, to consolidate individual EPCC codes from patient procedure records and to establish standardized procedure categories (‘specific procedures’). It also incorporates a hierarchical structure that prioritizes recognizable procedures, with the most complex procedures at the top and the least complex at the bottom. For our analysis, we employed specific procedure algorithm version 6.05, which defines 86 specific procedure categories.^22-24^ Similarly, an existing hierarchical scheme was used to assign a primary diagnosis for each record based on the first six (NCHDA approved) diagnostic codes submitted. If the diagnosis or procedure code suggested single-ventricle physiology, the record was coded as a functionally univentricular heart.^16,24,25^ **Supplementary Tables S2 and S3** provide the full list of diagnoses and specific procedures that could be allocated.

Among cardiac surgical procedures, the variables like specific procedure group, diagnosis group, activity type of procedure, functionally univentricular heart, indicators of acquired comorbidities, additional cardiac risk factor, congenital comorbidity, indicator of severity of illness, along with age and weight of the child at the time of the procedures were used to calculate Partial Risk Adjustment in Surgery 2 (PRAIS2) score. ^26^ (refer to **Supplementary material section 1** for details) PRAIS2 is used in England to account for case mix when mortality following pediatric ‘cardiac surgery’ is being compared between hospitals.^26^

The mortality models were adjusted for case mix (see *statistical methods section* below). For this, we used the individual risk factors for mortality that contribute to the PRAIS2 score - indicators of acquired comorbidity, additional cardiac risk factor, congenital comorbidity, and severity of illness. We used the individual risk factors, rather than the derived score, as PRAIS2 has been developed to assess case mix for ‘cardiac surgery’ alone, and our main analyses included all procedures (not solely surgery). **Supplementary Table S4** lists all the risk factors from PRAIS2 that we adjusted for.

*Subgroup variables*

We hypothesized that associations between the different pandemic management periods and outcomes might vary by child age, ethnicity, and family socioeconomic position, and explore this in subgroup analyses (see *statistical methods section* below).

Information about ethnicity was obtained primarily from NCHDA, with the latest documented ethnicity in primary care data used if there was no entry or ‘unknown ethnicity’ documented in the former source. We generated four ethnicity groups for the exploratory subgroup analyses: White European, South Asian, African/Caribbean, and other ethnicities, with the latter including mixed White and Black Caribbean, mixed White and Black African, mixed White and Asian, any other mixed background, Chinese and any other ethnic groups, each of which had too few participants for robust analyses. As an indicator of family socioeconomic position, we used a small area-level measure of deprivation, the 2011 English Index of multiple deprivation (IMD) score. IMD is derived by the English government to assess relative deprivation across low-level geographical areas of an average of 1700 people. Each area is assessed across seven domains (income, employment, health deprivation and disability, education skills and training, and living environment) using information on 39 variables. ^27^ We undertook subgroup analyses within quintiles of the IMD score, with the lowest quintile reflecting those from the most deprived residential areas.

### Estimation of the PRAiS 2 risk score

The estimation is based on the (Rogers et al., 2017) paper. The variables used in the PRAiS 2 model are mentioned below. This is the documentation of the decisions needed in creating the variables to include into the equation.

Rogers, L., Brown, K.L., Franklin, R.C., Ambler, G., Anderson, D., Barron, D.J., Crowe, S., English, K., Stickley, J., Tibby, S., 2017. Improving risk adjustment for mortality after pediatric cardiac surgery: the UK PRAiS2 model. The Annals of thoracic surgery 104, 211–219.

Diagnosis grouping: This is based on the supplementary materials in the above journal. The first six stated diagnosis in record was converted into diagnosis ranking (1 to 29) and diagnosis ranking for PRAiS model (1 to 11 groups) [‘Diagnoses’ sheet from the supplementary material]

Specific procedure grouping: Converting the specific procedure algorithm groups into 15 specific grouping and one group with ‘no specific procedure’ group. As per Rogers et al 2017., only procedures which were classified as ‘bypass’ and ‘non-bypass’ procedures were used to estimate the PRAiS2 score. The mapping of the specific algorithm groups into the 15 specific group was performed using the ‘SpecificProcedures’ sheet of the supplementary material and ‘Specific Procedure Coding v5.05 25_05_16.xlsx’ sheet available from the NCHDA website ([CCAD - Congenital Analysis - Technical Information (nicor.org.uk)](https://nicor4.nicor.org.uk/CHD/an_paeds.nsf/vwContent/Technical%20Information?Opendocument)

The final PRAiS 2 model calibrated on all data from 2009-2015 is as follows:

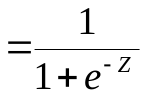

Probability of death within 30 days following paediatric cardiac surgery	where

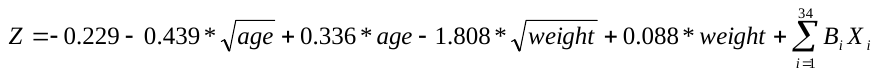

Parameters i =1 to 34 are tabulated below along with their corresponding regression coefficients,$B_{i}$ , and the condition that must be satisfied for $X_{i}=1$ ($X_{i}=0$ otherwise).

| 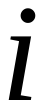 | 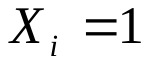 **if condition satisfied (**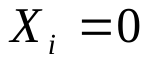 **otherwise)** | 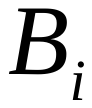 | **p-value** |
| --- | --- | --- | --- |
| 1 | Diagnosis grouping 1 | 0.000 | - |
| 2 | Diagnosis grouping 2 | -0.168 | - |
| 3 | Diagnosis grouping 3 | -0.330 | - |
| 4 | Diagnosis grouping 4 | -1.521 | - |
| 5 | Diagnosis grouping 5 | -0.512 | - |
| 6 | Diagnosis grouping 6 | -0.117 | - |
| 7 | Diagnosis grouping 7 | -0.054 | - |
| 8 | Diagnosis grouping 8 | -0.631 | - |
| 9 | Diagnosis grouping 9 | -0.468 | - |
| 10 | Diagnosis grouping 10 | -1.698 | - |
| 11 | Diagnosis grouping 11 | -1.241 | - |
| 12 | Specific procedure grouping 1 | 0.000 | - |
| 13 | Specific procedure grouping 2 | 0.216 | - |
| 14 | Specific procedure grouping 3 | 0.625 | - |
| 15 | Specific procedure grouping 4 | -0.090 | - |
| 16 | Specific procedure grouping 5 | 0.056 | - |
| 17 | Specific procedure grouping 6 | -0.747 | - |
| 18 | Specific procedure grouping 7 | 1.066 | - |
| 19 | Specific procedure grouping 8 | 0.788 | - |
| 20 | Specific procedure grouping 9 | 1.100 | - |
| 21 | Specific procedure grouping 10 | -0.787 | - |
| 22 | Specific procedure grouping 11 | -0.964 | - |
| 23 | Specific procedure grouping 12 | -0.202 | - |
| 24 | Specific procedure grouping 13 | -0.067 | - |
| 25 | Specific procedure grouping 14 | -0.937 | - |
| 26 | Specific procedure grouping 15 | -1.637 | - |
| 27 | Specific procedure grouping 20 (no specific procedure) | 0.428 | - |
| 28 | Bypass procedure | 0.398 | 0.01 |
| 29 | Definite indication of univentricular heart | 0.692 | <0.01 |
| 30 | Additional Cardiac Risk factor | 0.731 | <0.01 |
| 31 | Acquired Comorbidity | 0.538 | <0.01 |
| 32 | Congenital Comorbidity | 0.325 | 0.01 |
| 33 | Severity of Illness Indicator | 0.689 | <0.01 |
| 34 | Procedures from 2013 onwards | -0.280 | <0.01 |

*Note that for prospective use, the “Post 2013” coefficient can be absorbed into the constant term.*

### Supplementary Table S1: Code list for post operative complications

| **Complication** | **Numerical European Paediatric Cardiac Code (EPCC)* and clinical description of complication** |
| --- | --- |
| Unplanned reoperation | 124307. Unplanned reoperation/ reintervention during current admission (excludes bleeding) |
| Extracorporeal membrane oxygenation (ECMO) | 150009. Requirement for mechanical circulatory support (including ECMO) |
| Surgical site infection | 156741. Surgical site infection requiring surgical intervention |
| Pleural effusion | 158064. Prolonged pleural drainage > 7 days |
|  | 158065. Postprocedural prolonged pleural drainage (over 10 days) |
| Necrotizing enterocolitis | 158375. Postprocedural necrotizing enterocolitis - established requiring treatment |
| Other complications | 158086. Postprocedural requirement for tracheostomy |
|  | 158190. Phrenic nerve injury requiring plication of diaphragm |
|  | 158213. Acute kidney injury requiring dialysis |
|  | 158257. New neurological impairment (global or focal) present at discharge |
|  | 110633. Postprocedural complete atrioventricular block requiring permanent pacemaker system |
|  | 159014. Procedure related complication |

*NCHDA uses the Association for European Paediatric and Congenital Cardiology derived version of the International Paed*atric and Congenital Cardiac Code (*[*www.ipccc.net*](http://www.ipccc.net)*), whose derived Short list is known as the ‘European Peadiatric Cardiac Code’*

### Supplementary Table S2: PRAIS 2 diagnosis groups

| **RECOGNISED NCHDA OFFICIAL DIAGNOSES*** | **Diagnosis grouping used in PRAiS2** | **Univentricular heart function** |
| --- | --- | --- |
| 010109. Hypoplastic left heart syndrome | 1 | Yes |
| 060201. Mitral atresia | 1 | Yes |
| 091503. Aortic atresia | 1 | Yes |
| 090101. Common arterial trunk (truncus arteriosus) | 1 | No |
| 090200. Truncal valvar abnormality | 1 | No |
| 090203. Truncal valvar regurgitation | 1 | No |
| 010107. Pulmonary atresia + intact ventricular septum | 1 | Yes |
| 010114. Double inlet AV connection (double inlet ventricle) | 2 | Yes |
| 010122. Functionally univentricular heart | 2 | Yes |
| 010124. Double outlet right ventricle: with intact ventricular septum | 2 | Yes |
| 010403. Double inlet RV | 2 | Yes |
| 010404. Double inlet LV | 2 | Yes |
| 020305. Solitary ventricle of indeterminate morphology | 2 | Yes |
| 060101. Tricuspid atresia | 2 | Yes |
| 060726. AVSD with ventricular imbalance | 2 | Yes |
| 070841. Functionally univentricular heart | 2 | Yes |
| 070842. Functionally univentricular heart | 2 | Yes |
| 010106. Pulmonary atresia + VSD (including Fallot type) | 2 | No |
| 010125. Pulmonary atresia + VSD + systemic-to-pulmonary collateral artery(ies) (MAPCA(s)) | 2 | No |
| 090511. Pulmonary atresia | 2 | No |
| 090512. Pulmonary atresia: imperforate valve | 2 | No |
| 090726. Solitary arterial trunk (absent intrapericardial pulmonary arteries) | 2 | No |
| 090801. Major systemic-to-pulmonary collateral artery(ies) (MAPCA(s)) | 2 | No |
| 092025. Systemic-to-pulmonary collateral arter(ies) (MAPCA(s)) stenosis(es) | 2 | No |
| 010118. Double outlet right ventricle: transposition type (subpulmonary VSD) | 3 | No |
| 010501. Discordant VA connections (TGA) | 3 | No |
| 092931. Interrupted aortic arch | 3 | No |
| 092700. Arterial duct (ductus arteriosus) abnormality | 4 | No |
| 092721. Patent arterial duct (PDA) | 4 | No |
| 010103. Congenitally corrected transposition of great arteries (discordant AV & VA connections) | 5 | No |
| 010104. Double outlet right ventricle | 5 | No |
| 010116. Partially anomalous pulmonary venous connections: Scimitar syndrome | 5 | No |
| 010119. Double outlet right ventricle: with non-committed VSD | 5 | Yes |
| 010120. AV septal defect and Tetralogy of Fallot | 5 | No |
| 010133. Shone's syndrome: left heart obstruction at multiple sites, | 5 | No |
| 010140. Double outlet right ventricle: subaortic or doubly committed VSD without pulmonary stenosis ('VSD type') | 5 | No |
| 010503. Double outlet left ventricle | 5 | No |
| 040891. Pulmonary vein stenosis | 5 | No |
| 050201. Cor triatriatum (divided left atrium) | 5 | No |
| 090401. Aortopulmonary window | 5 | No |
| 090525. Absent pulmonary valve syndrome: Fallot-type | 5 | No |
| 091600. Supravalvar aortic stenosis | 5 | No |
| 093100. Vascular ring | 5 | No |
| 094101. Anomalous origin of coronary artery from pulmonary artery | 5 | No |
| 060100. Tricuspid valvar abnormality | 5 | No |
| 060103. Tricuspid valvar dysplasia | 5 | No |
| 060125. Tricuspid regurgitation: congenital | 5 | No |
| 060134. Ebstein's malformation of tricuspid valve | 5 | No |
| 060191. Tricuspid regurgitation | 5 | No |
| 060192. Tricuspid stenosis | 5 | No |
| 040600. Totally anomalous pulmonary venous connection: supracardiac | 5 | No |
| 040805. Totally anomalous pulmonary venous connection | 5 | No |
| 040806. Obstructed pulmonary venous connection(s) | 5 | No |
| 040810. Totally anomalous pulmonary venous connection: intracardiac | 5 | No |
| 040820. Totally anomalous pulmonary venous connection: infracardiac | 5 | No |
| 040830. Totally anomalous pulmonary venous connection: mixed | 5 | No |
| 120000. Totally anomalous pulmonary venous connection repair | 5 | No |
| 120002. Partially anomalous pulmonary venous connection repair | 5 | No |
| 120003. Pulmonary vein stenosis repair | 5 | No |
| 120017. Scimitar syndrome (PAPVC) repair | 5 | No |
| 120020. Pulmonary vein procedure | 5 | No |
| 120021. Balloon dilation of pulmonary vein | 5 | No |
| 120022. Stent placement in pulmonary vein | 5 | No |
| 120023. Balloon dilation of pulmonary vein using cutting balloon | 5 | No |
| 120024. Balloon dilation of pulmonary vein or pathway | 5 | No |
| 120025. Stent placement in pulmonary vein or pathway | 5 | No |
| 120029. Systemic venous pathway procedure (post Senning-Mustard) | 5 | No |
| 120030. Systemic vein procedure | 5 | No |
| 120036. Stent placement in superior caval vein (SVC) | 5 | No |
| 120039. Superior caval vein (SVC) procedure | 5 | No |
| 120042. Inferior caval vein (IVC) procedure | 5 | No |
| 120043. Balloon dilation of systemic vein or pathway | 5 | No |
| 120044. Stent placement in systemic vein or pathway | 5 | No |
| 120050. Coronary sinus procedure | 5 | No |
| 120055. Coronary sinus interatrial communication (ASD) repair | 5 | No |
| 120078. Partially anomalous pulmonary venous connection repair: baffle redirection to left atrium & systemic vein translocated to right atrial appendage (Warden), | 5 | No |
| 120081. Anomalous systemic venous connection repair | 5 | No |
| 120083. Systemic venous stenosis repair | 5 | No |
| 120090. Cardiopulmonary bypass used during procedure | 5 | No |
| 120091. Cardiopulmonary bypass not used during procedure | 5 | No |
| 120092. Cardiopulmonary bypass for non-cardiac lesion | 5 | No |
| 120100. Interatrial communication closure | 5 | No |
| 120101. Atrial septal defect (ASD) secundum closure | 5 | No |
| 120102. Atrial septal defect (ASD) secundum closure with direct suture | 5 | No |
| 120103. Atrial septal defect (ASD) secundum closure with patch | 5 | No |
| 120106. Atrial septal defect (ASD) secundum closure with transluminal device | 5 | No |
| 120107. PFO closure with transluminal device | 5 | No |
| 120108. Interatrial communication closure: partial | 5 | No |
| 120110. Sinus venosus defect (ASD) closure | 5 | No |
| 120122. Atrial septation procedure | 5 | No |
| 120130. Left atrial procedure | 5 | No |
| 120131. Cor triatriatum (divided left atrium) repair | 5 | No |
| 120132. Supra-mitral valvar LA-ring excision | 5 | No |
| 120141. Balloon atrial septostomy by pull back (Rashkind) | 5 | No |
| 120142. Atrial septectomy: closed (Blalock Hanlon) | 5 | No |
| 120143. Atrial septectomy | 5 | No |
| 120144. Blade atrial septostomy | 5 | No |
| 120147. Transluminal fenestration of atrial septum-tunnel | 5 | No |
| 120153. Patent foramen ovale (PFO) direct closure | 5 | No |
| 120157. Atrial baffle procedure | 5 | No |
| 120160. Right atrial procedure | 5 | No |
| 120180. Interatrial communication left open | 5 | No |
| 120190. Interatrial communication creation-enlargement | 5 | No |
| 120198. Interatrial communication closure with transluminal device | 5 | No |
| 120200. Tricuspid valvar procedure | 5 | No |
| 120202. Tricuspid leaflet (valvoplasty) procedure | 5 | No |
| 120204. Tricuspid valvar annuloplasty | 5 | No |
| 120209. Ebstein's malformation of tricuspid valve repair | 5 | No |
| 120211. Tricuspid valvar replacement | 5 | No |
| 120222. Tricuspid valvectomy | 5 | No |
| 120270. Tricuspid valvar closure | 5 | No |
| 120283. Tricuspid valve repair converted to tricuspid valvar replacement, | 5 | No |
| 120300. Mitral valvar procedure | 5 | No |
| 120301. Mitral valvotomy | 5 | No |
| 120303. Mitral leaflet (valvoplasty) procedure | 5 | No |
| 120304. Mitral valvar annuloplasty | 5 | No |
| 120310. Balloon mitral valvotomy | 5 | No |
| 120311. Mitral valvar replacement | 5 | No |
| 120319. Mitral subvalvar apparatus procedure | 5 | No |
| 120384. Mitral valve repair converted to mitral valvar replacement, | 5 | No |
| 120397. Transluminal mitral valve repair | 5 | No |
| 120400. AV septal defect procedure | 5 | No |
| 120401. AVSD: partial (primum ASD) repair | 5 | No |
| 120409. AVSD: partial with isolated ventricular component (VSD) repair | 5 | No |
| 120418. Common atrioventricular valve replacement, | 5 | No |
| 120420. AVSD: right AV valvar procedure | 5 | No |
| 120433. Common atrioventricular valve repair converted to atrioventricular valvar replacement, | 5 | No |
| 120440. AVSD: left AV valvar procedure | 5 | No |
| 120445. AVSD: left AV valvar replacement | 5 | No |
| 120501. AVSD: complete (common valve orifice) repair | 5 | No |
| 120510. AVSD: 'intermediate' repair | 5 | No |
| 120511. AVSD & Tetralogy of Fallot repair | 5 | No |
| 120600. Right ventricular outflow tract procedure | 5 | No |
| 120605. Balloon dilation of right ventricular outflow tract | 5 | No |
| 120618. Stent placement in right ventricular outflow tract | 5 | No |
| 120619. 1.5 ventricle repair: Glenn anastomosis + right ventricular outflow tract reconstruction | 5 | No |
| 120625. Transluminal right ventricular biopsy | 5 | No |
| 120626. Right ventricular procedure | 5 | No |
| 120635. Double chambered right ventricle repair | 5 | No |
| 120638. Right ventricular aneurysm repair | 5 | No |
| 120641. Right ventricular outflow tract obstruction relief | 5 | No |
| 120643. Right ventricle to pulmonary artery valveless conduit construction (including 'Sano') | 5 | No |
| 120700. Left ventricular outflow tract procedure | 5 | No |
| 120701. Subaortic fibromuscular shelf resection | 5 | No |
| 120707. Balloon dilation of left ventricular outflow tract | 5 | No |
| 120711. Left ventricular outflow tract myectomy-myotomy | 5 | No |
| 120712. Left ventricular outflow tract obstruction relief: complex (Konno etc) | 5 | No |
| 120713. Left ventricular outflow tract obstruction relief | 5 | No |
| 120719. Left ventricular outflow tract obstruction relief by transcatheter coronary chemical ablation | 5 | No |
| 120726. Left ventricular procedure | 5 | No |
| 120737. Left ventricular aneurysm repair | 5 | No |
| 120738. Partial left ventriculectomy-volume reduction (Batista) | 5 | No |
| 120801. VSD closure | 5 | No |
| 120802. VSD closure by direct suture | 5 | No |
| 120803. VSD closure using patch | 5 | No |
| 120806. VSD enlargement | 5 | No |
| 120807. VSD closure with transluminal device | 5 | No |
| 120816. Closure of multiple VSDs | 5 | No |
| 120819. Open fenestration of VSD patch | 5 | No |
| 120820. Transluminal fenestration of VSD patch | 5 | No |
| 120821. Subpulmonary obstruction relief | 5 | No |
| 120822. Subaortic obstruction relief | 5 | No |
| 120828. Intraoperative VSD closure with transluminal device (hybrid approach) | 5 | No |
| 120835. VSD enlargement/ creation | 5 | No |
| 120865. Transluminal interventricular communication creation | 5 | No |
| 120901. Ventricular septation procedure | 5 | No |
| 120903. Damus-Kaye-Stansel type procedure: pulmonary trunk to aorta end/side anastomosis | 5 | No |
| 121000. Norwood type procedure | 5 | No |
| 121004. Application of bilateral pulmonary arterial bands & transcatheter placement of stent in arterial duct | 5 | No |
| 121005. Hypoplastic left heart biventricular repair | 5 | No |
| 121014. Stent placement in arterial duct (PDA) | 5 | No |
| 121100. Common arterial trunk (truncus) repair | 5 | No |
| 121134. Truncal valve cusp(s) repair (valvoplasty) | 5 | No |
| 121141. Truncal valve replacement | 5 | No |
| 121143. Truncal valve repair converted to truncal valvar replacement, | 5 | No |
| 121201. Aortopulmonary window closure | 5 | No |
| 121208. Aortopulmonary window closure with transcatheter device | 5 | No |
| 121300. Pulmonary valvar procedure | 5 | No |
| 121302. Pulmonary valvotomy: open | 5 | No |
| 121305. Balloon pulmonary valvotomy | 5 | No |
| 121309. Pulmonary valvar transluminal perforation & dilation | 5 | No |
| 121312. Pulmonary valvectomy | 5 | No |
| 121315. Pulmonary valve closure-oversewing | 5 | No |
| 121321. Pulmonary valvar replacement (not conduit) | 5 | No |
| 121322. Pulmonary valvar replacement using homograft | 5 | No |
| 121351. Transluminal pulmonary valvar insertion with stent mounted valve | 5 | No |
| 121355. Pulmonary valve repair converted to pulmonary valvar replacement, | 5 | No |
| 121381. Transluminal aortic valvar insertion with stent mounted valve | 5 | No |
| 121384. Transapical aortic valve implantation (hybrid approach) | 5 | No |
| 121401. Pulmonary trunk arterioplasty | 5 | No |
| 121402. Pulmonary trunk band (PA band) | 5 | No |
| 121403. Pulmonary trunk band removal (de-band) | 5 | No |
| 121405. Balloon dilation of pulmonary trunk | 5 | No |
| 121419. Application of right & left pulmonary arterial bands, | 5 | No |
| 121420. Pulmonary arterioplasty/ reconstruction | 5 | No |
| 121421. Pulmonary arterioplasty/ reconstruction: central (proximal to hilar bifurcation) | 5 | No |
| 121422. Pulmonary arterioplasty/ reconstruction: peripheral (at-beyond hilar bifurcation) | 5 | No |
| 121430. Pulmonary artery origin from ascending aorta (hemitruncus) repair | 5 | No |
| 121431. Pulmonary artery ligation | 5 | No |
| 121503. Balloon dilation of right pulmonary artery | 5 | No |
| 121504. Balloon dilation of left pulmonary artery | 5 | No |
| 121511. Procedure involving pulmonary artery | 5 | No |
| 121513. Stent placement in right pulmonary artery | 5 | No |
| 121514. Stent placement in left pulmonary artery | 5 | No |
| 121524. Pulmonary aneurysm repair | 5 | No |
| 121525. Pulmonary trunk flow restriction using transcatheter implanted device | 5 | No |
| 121548. Pulmonary thromboembolectomy | 5 | No |
| 121549. Transluminal embolectomy from pulmonary tree | 5 | No |
| 121550. Stent placement in pulmonary tree | 5 | No |
| 121553. Balloon dilation of pulmonary tree with cutting balloon | 5 | No |
| 121580. Pulmonary thromboembolectomy for acute embolus | 5 | No |
| 121581. Pulmonary thromboembolectomy for chronic (longstanding) embolus | 5 | No |
| 121600. Aortic valvar procedure | 5 | No |
| 121602. Aortic valvotomy: open | 5 | No |
| 121604. Aortic valvotomy: closed | 5 | No |
| 121605. Balloon aortic valvotomy | 5 | No |
| 121611. Aortic cusp(s) repair (valvoplasty) | 5 | No |
| 121614. Annuloplasty' of aortic valve | 5 | No |
| 121621. Aortic valvar replacement | 5 | No |
| 121622. Aortic valvar replacement using homograft | 5 | No |
| 121625. Aortic valvar transluminal perforation & dilation | 5 | No |
| 121628. Aortic valvar replacement using heterograft bioprosthesis | 5 | No |
| 121629. Aortic valvar replacement using mechanical prosthesis | 5 | No |
| 121630. Ross procedure: aortic valve or root replacement with pulmonary autograft & pulmonary valvar replacement | 5 | No |
| 121633. Aortic root replacement + coronary artery reimplantation (Bentall) | 5 | No |
| 121635. Ascending aorta replacement & aortic valvar resuspension | 5 | No |
| 121640. Supravalvar aortic stenosis repair | 5 | No |
| 121642. Aorta aneurysm repair | 5 | No |
| 121650. Aortic root replacement (not Ross) | 5 | No |
| 121659. Aortic dissection repair | 5 | No |
| 121661. Aortic valve closure-oversewing | 5 | No |
| 121662. Ross-Konno procedure | 5 | No |
| 121663. Aortic root replacement using homograft | 5 | No |
| 121664. Aortic root replacement using mechanical prosthesis | 5 | No |
| 121665. Ascending aorta replacement | 5 | No |
| 121666. Aortic arch aneurysm repair | 5 | No |
| 121667. Descending aorta aneurysm repair | 5 | No |
| 121668. Abdominal aorta aneurysm repair | 5 | No |
| 121680. Aortic sinus of Valsalva procedure | 5 | No |
| 121681. Aortic sinus of Valsalva distal fistula closure | 5 | No |
| 121685. Aortic sinus of Valsalva aneurysm repair | 5 | No |
| 121690. Aorto-left ventricular tunnel closure | 5 | No |
| 121697. Aortic valve repair converted to aortic valvar replacement, | 5 | No |
| 121711. Vascular ring procedure | 5 | No |
| 121731. Aortopexy | 5 | No |
| 121732. Pulmonary arterial sling repair | 5 | No |
| 121790. Aortic root replacement using bioprosthesis | 5 | No |
| 121791. Aortic root replacement: valve sparing technique | 5 | No |
| 121800. Coarctation-hypoplasia of aorta repair | 5 | No |
| 121801. Aortic coarctation-hypoplasia repair by resection & end to end anastomosis | 5 | No |
| 121802. Aortic coarctation-hypoplasia repair by patch aortoplasty | 5 | No |
| 121803. Aortic coarctation-hypoplasia repair by subclavian flap aortoplasty | 5 | No |
| 121804. Balloon dilation of native aortic coarctation-hypoplasia | 5 | No |
| 121808. Balloon dilation of aortic recoarctation | 5 | No |
| 121810. Aortic coarctation-hypoplasia repair by resection & extended end to end anastomosis | 5 | No |
| 121815. Aortic coarctation-hypoplasia repair by resection & insertion of tube graft | 5 | No |
| 121817. Stent placement at site of aortic coarctation | 5 | No |
| 121822. Stent placement at site of aortic recoarctation | 5 | No |
| 121827. Aortic coarctation transluminal obstruction relief | 5 | No |
| 121830. Aortic arch repair | 5 | No |
| 121848. Stent placement at site of native aortic coarctation-hypoplasia | 5 | No |
| 121870. Thoracic aorta aneurysm transcatheter stent implantation | 5 | No |
| 122020. Hypoplastic left heart syndrome hybrid approach (transcatheter & surgery): stage 1, | 5 | No |
| 122021. Hypoplastic left heart syndrome hybrid approach (transcatheter & surgery) | 5 | No |
| 122022. Hypoplastic left heart syndrome hybrid approach (transcatheter & surgery) 'stage 2': aortopulmonary amalgamation + superior cavopulmonary anastomosis(es) + debanding of pulmonary arteries, | 5 | No |
| 122023. Hypoplastic left heart syndrome hybrid approach (transcatheter & surgery) 'stage 2': aortopulmonary amalgamation + superior cavopulmonary anastomosis(es) + debanding of pulmonary arteries + arch repair, | 5 | No |
| 122100. Interrupted aortic arch repair | 5 | No |
| 122200. Systemic arterial procedure | 5 | No |
| 122300. Anomalous coronary artery (eg ALCAPA) repair | 5 | No |
| 122307. Coronary fistula procedure | 5 | No |
| 122308. Coronary arterial bypass graft (CABG) procedure | 5 | No |
| 122309. Coronary arterial procedure | 5 | No |
| 122311. Ligation of coronary fistula | 5 | No |
| 122313. Coronary arterial fistula transluminal occlusion | 5 | No |
| 122330. Transluminal intracoronary injection of thrombolytic agent | 5 | No |
| 122331. Transluminal balloon coronary angioplasty (PTCA) | 5 | No |
| 122338. Transluminal coronary stent implantation | 5 | No |
| 122342. Transluminal chemical occlusion of coronary artery | 5 | No |
| 122380. Anomalous aortic origin of coronary artery repair, | 5 | No |
| 122400. Arterial duct-ligament procedure | 5 | No |
| 122404. Arterial duct (PDA) closure with transluminal device | 5 | No |
| 122410. Arterial duct (PDA) closure | 5 | No |
| 122420. Patent arterial duct (PDA) closure: surgical | 5 | No |
| 122421. Arterial duct (PDA) closure with transluminal coil | 5 | No |
| 122422. Arterial duct (PDA) closure with transluminal Amplatzer plug | 5 | No |
| 122500. Systemic-to-pulmonary collateral artery(ies) (MAPCA(s)) unifocalisation procedure | 5 | No |
| 122502. Arteriovenous fistula occlusion | 5 | No |
| 122507. Pulmonary arteriovenous fistula closure | 5 | No |
| 122518. Systemic-to-pulmonary collateral artery(ies) (MAPCA(s)) occlusion | 5 | No |
| 122519. Transluminal procedure to systemic-to-pulmonary collateral artery (MAPCA(s)) | 5 | No |
| 122562. Stent placement in systemic-to-pulmonary collateral artery (MAPCA(s)) | 5 | No |
| 122565. Transluminal occlusion of systemic-to-pulmonary collateral artery(ies) (MAPCA(s)) with coil-device | 5 | No |
| 122572. Balloon dilation of systemic-to-pulmonary collateral artery(ies) (MAPCA(s)) | 5 | No |
| 122601. Tetralogy of Fallot repair | 5 | No |
| 122613. Tetralogy of Fallot repair with transannular patch | 5 | No |
| 122620. Tetralogy of Fallot repair without transannular patch | 5 | No |
| 122621. Absent pulmonary valve syndrome (Fallot type) repair | 5 | No |
| 122701. Double outlet right ventricle with subaortic or doubly committed VSD & pulmonary stenosis (Fallot-type) repair | 5 | No |
| 122702. Double outlet right ventricle repair with intraventricular tunnel | 5 | No |
| 122745. REV procedure: intraventricular left ventricle to aorta tunnel with infundibular septum resection & direct right ventricle to pulmonary trunk anastomosis | 5 | No |
| 122746. Congenitally corrected transposition of great arteries repair | 5 | No |
| 122750. Double outlet left ventricle repair | 5 | No |
| 122778. Aortic root translocation to over left ventricle (including Nikaidoh), | 5 | No |
| 122801. Pulmonary atresia & VSD (including Fallot-type) repair | 5 | No |
| 122811. Pulmonary atresia, VSD & systemic-to-pulmonary collateral artery(ies) (MAPCA(s)) repair | 5 | No |
| 122901. Senning procedure (atrial inversion) | 5 | No |
| 122902. Mustard procedure (atrial inversion) | 5 | No |
| 122911. Rastelli procedure: intraventricular left ventricle to aorta tunnel & right ventricle to pulmonary artery conduit | 5 | No |
| 122920. Double outlet right ventricle repair | 5 | No |
| 122921. Arterial switch procedure | 5 | No |
| 122925. Arterial switch & atrial inversion procedures ('double switch') | 5 | No |
| 122926. Atrial inversion and Rastelli procedures | 5 | No |
| 122940. Complex transposition of great arteries repair | 5 | No |
| 122952. Pulmonary venous pathway procedure (post Senning-Mustard) | 5 | No |
| 122979. Atrial inversion procedure (Mustard or Senning) revision | 5 | No |
| 123001. Fontan type procedure | 5 | No |
| 123005. Total cavopulmonary connection (TCPC) using extracardiac inferior caval vein (IVC)-pulmonary artery conduit with fenestration, | 5 | No |
| 123006. Total cavopulmonary connection (TCPC) with fenestrated lateral atrial tunnel, | 5 | No |
| 123013. Fontan procedure with atrioventricular connection | 5 | No |
| 123020. Fenestration of atrial septum | 5 | No |
| 123021. Right atrial septum-tunnel fenestration closure with transluminal device | 5 | No |
| 123027. Fenestration of Fontan type connection | 5 | No |
| 123028. Fontan-type connection without fenestration | 5 | No |
| 123031. Takedown of Fontan type procedure | 5 | No |
| 123032. Fontan procedure with direct atriopulmonary anastomosis | 5 | No |
| 123034. Conversion of Fontan repair to total cavopulmonary connection | 5 | No |
| 123037. Fontan type procedure revision or conversion | 5 | No |
| 123041. Atrial fenestration closure | 5 | No |
| 123050. Total cavopulmonary connection (TCPC) | 5 | No |
| 123051. Total cavopulmonary conn (TCPC) with lateral atrial tunnel | 5 | No |
| 123054. Total cavopulmonary connection (TCPC) using extracardiac inferior caval vein (IVC)-pulmonary artery conduit | 5 | No |
| 123056. Takedown of total cavopulmonary connection (TCPC) | 5 | No |
| 123060. Completion of total cavopulmonary connection (TCPC) using transcatheter inferior to superior caval vein covered stent | 5 | No |
| 123074. Transluminal interatrial communication creation | 5 | No |
| 123103. Modified right Blalock interposition shunt | 5 | No |
| 123104. Modified left Blalock interposition shunt | 5 | No |
| 123105. Waterston (ascending aorta-right pulmonary artery) anastomosis | 5 | No |
| 123106. Central systemic-to-pulmonary arterial interposition shunt | 5 | No |
| 123111. Bidirectional superior cavopulmonary (Glenn) anastomosis | 5 | No |
| 123115. Hemi-Fontan procedure | 5 | No |
| 123119. Balloon dilation of systemic-PA shunt | 5 | No |
| 123130. Systemic-to-pulmonary arterial shunt procedure | 5 | No |
| 123131. Closure of systemic-to-pulmonary arterial shunt | 5 | No |
| 123134. Occlusion of systemic-PA shunt by transluminal embolus/device | 5 | No |
| 123142. Takedown of Glenn | 5 | No |
| 123144. Bilateral bidirectional superior cavopulmonary (Glenn) anastomoses | 5 | No |
| 123145. Unidirectional superior cavopulmonary (Glenn) anastomosis | 5 | No |
| 123146. Modified Blalock interposition shunt | 5 | No |
| 123172. Superior caval vein to pulmonary artery anastomosis, | 5 | No |
| 123200. Post-operative procedure | 5 | No |
| 123206. Lung biopsy procedure | 5 | No |
| 123209. Pericardiectomy | 5 | No |
| 123210. Heart tumour resection | 5 | No |
| 123213. Transplantation of heart and lungs | 5 | No |
| 123214. DC cardioversion | 5 | No |
| 123217. Parietal pleurectomy | 5 | No |
| 123218. Post-operative procedure to control bleeding | 5 | No |
| 123220. Removal of cardiac vegetations | 5 | No |
| 123221. Cardiac procedure | 5 | No |
| 123222. Removal of cardiac thrombus | 5 | No |
| 123228. Thoracic duct occlusion | 5 | No |
| 123229. Diaphragm procedure, | 5 | No |
| 123240. Pericardiocentesis | 5 | No |
| 123241. Pericardial drainage: open (pericardiotomy) | 5 | No |
| 123243. Pericardiocentesis: percutaneous transcatheter | 5 | No |
| 123246. Pericardial window creation | 5 | No |
| 123253. Pericardial biopsy | 5 | No |
| 123259. Procedure involving pericardium | 5 | No |
| 123270. Plication of hemidiaphragm | 5 | No |
| 123280. Insertion of pleural tube drain | 5 | No |
| 123283. Insertion of mediastinal tube drain | 5 | No |
| 123290. Instigation of renal dialysis | 5 | No |
| 123310. Traumatic injury of heart repair | 5 | No |
| 123351. Peripheral vascular procedure | 5 | No |
| 123352. Non-cardiothoracic-vascular procedure | 5 | No |
| 123353. Non-cardiothoracic-vascular procedure on cardiac patient under cardiac anaesthesia, | 5 | No |
| 123360. Operation related to transcatheter procedure | 5 | No |
| 123370. Medical management of pericardial disease | 5 | No |
| 123450. Pacemaker system placement: single chamber | 5 | No |
| 123451. Pacemaker system placement: dual chamber | 5 | No |
| 123452. Pacemaker system placement: biventricular | 5 | No |
| 123460. Pacemaker system placement: temporary | 5 | No |
| 123463. Pacemaker system placement: permanent epicardial | 5 | No |
| 123464. Pacemaker system placement: permanent endocardial | 5 | No |
| 123467. Pacemaker system placement: permanent | 5 | No |
| 123468. Pacemaker procedure | 5 | No |
| 123470. Pacemaker wire procedure | 5 | No |
| 123473. Cardiac resynchronisation therapy (biventricular pacing) | 5 | No |
| 123484. Pacemaker wire revision procedure | 5 | No |
| 123485. Pulse generator box placement | 5 | No |
| 123513. Pulse generator box replacement | 5 | No |
| 123514. Removal of complete implanted cardiac pacemaker system, | 5 | No |
| 123540. Arrhythmia surgical procedure | 5 | No |
| 123546. Transluminal cryoablation procedure for arrhythmia | 5 | No |
| 123548. Transluminal radiofrequency ablation procedure for arrhythmia | 5 | No |
| 123553. Maze operation | 5 | No |
| 123557. Transluminal procedure for arrhythmia | 5 | No |
| 123560. Pacing to abolish arrhythmia | 5 | No |
| 123572. Cox-Maze IV procedure, | 5 | No |
| 123580. Surgical ablation procedure for atrial arrhythmia | 5 | No |
| 123581. Surgical ablation procedure for ventricular arrhythmia | 5 | No |
| 123582. Transluminal procedure for atrial arrhythmia | 5 | No |
| 123583. Transluminal procedure for ventricular arrhythmia | 5 | No |
| 123584. Transluminal ablation procedure with pulmonary vein exclusion | 5 | No |
| 123600. Conduit construction procedure | 5 | No |
| 123601. Right ventricle to pulmonary arterial tree conduit construction | 5 | No |
| 123602. Left ventricle to pulmonary artery conduit construction | 5 | No |
| 123610. Replacement of cardiac conduit | 5 | No |
| 123614. Balloon dilation of cardiac conduit | 5 | No |
| 123623. Stent placement in cardiac conduit | 5 | No |
| 123635. Ventricle to aorta conduit construction | 5 | No |
| 123640. Procedure involving constructed cardiac conduit-shunt | 5 | No |
| 123701. Heart transplant | 5 | No |
| 123702. Transplantation of heart: orthotopic allotransplant | 5 | No |
| 123703. Transplantation of heart: heterotopic (piggy back) allotransplant | 5 | No |
| 123704. Prosthetic heart implantation | 5 | No |
| 123706. Transplantation of heart: ABO incompatible donor | 5 | No |
| 123713. Single lung transplant | 5 | No |
| 123720. Double lung transplant | 5 | No |
| 123760. Lung(s) transplant, | 5 | No |
| 123770. Organ procurement for transplantation | 5 | No |
| 123825. Transluminal left atrial appendage occlusion with device, | 5 | No |
| 123840. Transluminal ablation procedure for arrhythmia, | 5 | No |
| 123850. Surgical localisation and mapping procedure for arrhythmia | 5 | No |
| 123869. Percutaneous radiofrequency epicardial ablation procedure for arrhythmia, | 5 | No |
| 124000. Thoracotomy, | 5 | No |
| 124006. Thoracoscopic approach (VATS) | 5 | No |
| 124013. Minimally invasive procedure | 5 | No |
| 124015. Robotic surgical approach | 5 | No |
| 124016. Thoracotomy: redo | 5 | No |
| 124029. Median sternotomy: redo x 1-3 | 5 | No |
| 124030. Median sternotomy: redo x 4 or more | 5 | No |
| 124099. Cardiac incision, | 5 | No |
| 124130. Hybrid approach (combined surgical & transluminal) | 5 | No |
| 124231. Implantable cardioverter & defibrillator (ICD) implantation | 5 | No |
| 124233. Automatic cardioverter & defibrillator (AICD) transluminal implantation | 5 | No |
| 124234. Implantable cardioverter & defibrillator (ICD) system removal | 5 | No |
| 124235. Automatic cardioverter & defibrillator (AICD) transluminal removal | 5 | No |
| 124239. Implantable cardioverter & defibrillator (AICD) procedure | 5 | No |
| 124261. Implantable cardioverter & defibrillator (ICD) implantation: single chamber | 5 | No |
| 124264. Implantable cardioverter & defibrillator (ICD) implantation: dual chamber | 5 | No |
| 124265. Implantable cardioverter & defibrillator (ICD) implantation: biventricular | 5 | No |
| 124300. Reoperation | 5 | No |
| 124325. Palliative procedure, | 5 | No |
| 124475. Removal of implanted pacemaker lead | 5 | No |
| 124500. Cardiovascular catheter procedure | 5 | No |
| 124501. Catheterisation transeptal approach with transeptal puncture | 5 | No |
| 124504. Transluminal retrieval of device or foreign body | 5 | No |
| 124506. Repeat cardiovascular catheter procedure for residual or recurrent lesion | 5 | No |
| 124507. Transluminal diagnostic test occlusion, | 5 | No |
| 124510. Stent redilation | 5 | No |
| 124511. Stent placement | 5 | No |
| 124512. Balloon dilation | 5 | No |
| 124513. Transluminal device implantation | 5 | No |
| 124514. Cardiovascular catherisation occlusion procedure with coil | 5 | No |
| 124515. Transluminal implantation of valve, | 5 | No |
| 124519. Transluminal prosthetic valve leak closure using device | 5 | No |
| 124521. Balloon dilation of valve, | 5 | No |
| 124528. Therapeutic cardiovascular catheter procedure, | 5 | No |
| 124530. Transluminal procedure for catheterisation complication | 5 | No |
| 124558. Transluminal therapeutic perforation to establish interchamber and/or intervessel communication, | 5 | No |
| 124559. Transluminal procedure using adjunctive therapy, | 5 | No |
| 124600. AV valvar procedure in double inlet ventricle | 5 | No |
| 124801. Common atrioventricular valvar leaflet (valvoplasty) procedure, | 5 | No |
| 124802. AVSD: suturing together superior + inferior bridging leaflets to left ventricular side of septum ('cleft') | 5 | No |
| 126400. Bronchoscopy | 5 | No |
| 126408. Bronchoscopic removal of foreign body | 5 | No |
| 126420. Tracheal procedure | 5 | No |
| 126421. Tracheostomy creation | 5 | No |
| 126440. Tracheobronchial reconstruction procedure | 5 | No |
| 126505. Mediastinal exploration | 5 | No |
| 126506. Mediastinal procedure | 5 | No |
| 126513. Pectus carinatum repair | 5 | No |
| 126514. Pectus excavatum repair | 5 | No |
| 126523. Anterior chest wall (pectus) repair | 5 | No |
| 126545. Debridement of chest wall incision | 5 | No |
| 126548. Sternal wire removal from previous sternotomy | 5 | No |
| 126556. Sternotomy wound drainage | 5 | No |
| 126560. Delayed closure of sternum | 5 | No |
| 126572. Open excision of pleural lesion | 5 | No |
| 126582. Pleurodesis | 5 | No |
| 126589. Pleural procedure | 5 | No |
| 126600. Lung procedure | 5 | No |
| 126601. Lung decortication | 5 | No |
| 126602. Lung mass excision | 5 | No |
| 126605. Lung lobectomy | 5 | No |
| 126606. Pneumonectomy | 5 | No |
| 126607. Lung sequestration repair | 5 | No |
| 127008. Venovenous collateral occlusion with device | 5 | No |
| 128000. Thoracic-mediastinal procedure | 5 | No |
| 128001. Oesophageal procedure | 5 | No |
| 128010. Intestinal procedure | 5 | No |
| 128037. Percutaneous feeding gastrostomy tube placement (PEG) | 5 | No |
| 128038. Laparotomy | 5 | No |
| 128701. Cardiac support procedure | 5 | No |
| 128702. Intra-aortic balloon pump (IABP) removal: transluminal | 5 | No |
| 128704. Intra-aortic balloon pump (IABP) insertion: open | 5 | No |
| 128710. Intra-aortic balloon pump (IABP) insertion | 5 | No |
| 128721. Ventricular assist device implantation | 5 | No |
| 128722. Right ventricular assist device implantation | 5 | No |
| 128723. Left ventricular assist device implantation | 5 | No |
| 128724. Biventricular assist device implantation | 5 | No |
| 128725. Cardiac support using Extracorporeal Membrane Oxygenation (ECMO) circuitry | 5 | No |
| 128728. Procedure involving Extracorporeal Membrane Oxygenation (ECMO) circuitry, | 5 | No |
| 128731. Cardiomyoplasty procedure | 5 | No |
| 128741. Ventricular assist device removal | 5 | No |
| 128745. Take down of Extracorporeal Membrane Oxygenation (ECMO) circuitry | 5 | No |
| 129001. Atrioventricular valvar repair, | 5 | No |
| 130123. Transcatheter intervention with intracardiac echo guidance (ICE) | 5 | No |
| 130501. Diagnostic cardiovascular catheterisation procedure | 5 | No |
| 130505. Diagnostic cardiovascular catheterisation procedure: angiographic data obtained, | 5 | No |
| 130506. Diagnostic cardiovascular catheterisation procedure: haemodynamic data obtained, | 5 | No |
| 130507. Diagnostic cardiovascular catheterisation procedure with haemodynamic alteration (challenge), | 5 | No |
| 130508. Diagnostic cardiovascular catheterisation procedure with electrophysiological alteration (challenge), | 5 | No |
| 130512. Electrophysiological study (EPS) | 5 | No |
| 130513. Catheterisation study for pulmonary hypertension evaluation | 5 | No |
| 130514. Transcatheter procedure undertaken with x-ray guidance | 5 | No |
| 130515. Transcatheter procedure undertaken with magnetic resonance imaging guidance | 5 | No |
| 130516. Transcatheter procedure undertaken with x-ray & magnetic resonance imaging guidance | 5 | No |
| 130517. Electrophysiological study (EPS) with three dimensional mapping | 5 | No |
| Q50811. - transatrial approach | 5 | No |
| Q50812. - trans-right ventricular approach | 5 | No |
| Q50813. - trans-right ventricular approach, | 5 | No |
| Q58530. - with sedation, | 5 | No |
| 030102. Visceral heterotaxy (abnormal arrangement thoraco-abdominal organs) | 5 | No |
| 030109. Position or morphology of thoraco-abdominal organs abnormal | 5 | No |
| 030209. Lung anomaly | 5 | No |
| 030214. Functionally congenital single lung | 5 | No |
| 030305. Tracheobronchial anomaly | 5 | No |
| 030603. Intestines malrotated | 5 | No |
| 030703. Spleen absent (asplenia) | 5 | No |
| 030704. Multiple spleens (polysplenia) | 5 | No |
| 100665. Preprocedural endocarditis | 5 | No |
| 101363. Elevated lung resistance for biventricular repair (> 6 Wood units) | 5 | No |
| 101364. Elevated lung resistance for heart transplant (> 4 Wood units) | 5 | No |
| 101365. Elevated lung resistance for univentricular repair (> 2 Wood units) | 5 | No |
| 101400. Secondary systemic hypertension | 5 | No |
| 101402. Primary (essential) systemic hypertension | 5 | No |
| 101444. Abdominal aorta aneurysm | 5 | No |
| 101445. Rupture of thoracic aortic aneurysm | 5 | No |
| 101446. Rupture of abdominal aortic aneurysm | 5 | No |
| 101454. Descending aorta dissection & distal propagation (DeBakey type III/ Stanford type B) | 5 | No |
| 101460. Systemic arteritis | 5 | No |
| 101501. Persistent pulmonary hypertension of the newborn (PFC) | 5 | No |
| 101505. Necrotising enterocolitis | 5 | No |
| 101512. Meconium aspiration | 5 | No |
| 102002. Preprocedural shock | 5 | No |
| 102003. Preprocedural arrhythmia | 5 | No |
| 102005. Preprocedural acidosis | 5 | No |
| 102006. Preprocedural coagulation disorder | 5 | No |
| 102007. Preprocedural renal failure (creatinine >176) | 5 | No |
| 102008. Preprocedural renal failure requiring dialysis | 5 | No |
| 102009. Preprocedural septicaemia | 5 | No |
| 102012. Preprocedural neurological impairment | 5 | No |
| 102013. Preprocedural cerebral abnormality on imaging | 5 | No |
| 102014. Preprocedural mechanical ventilatory support | 5 | No |
| 102015. Preprocedural mechanical circulatory support | 5 | No |
| 102016. Preprocedural pulmonary hypertension | 5 | No |
| 102017. Preprocedural tracheostomy | 5 | No |
| 102018. Preprocedural seizures | 5 | No |
| 102031. Preprocedural shock at time of surgery (persistent) | 5 | No |
| 102032. Preprocedural shock resolved by time of surgery | 5 | No |
| 102033. Preprocedural cardiopulmonary resuscitation (< 48 hours) | 5 | No |
| 102037. Preprocedural respiratory syncytial virus (RSV) infection | 5 | No |
| 102038. Preprocedural necrotising enterocolitis: treated medically | 5 | No |
| 102039. Preprocedural necrotising enterocolitis: treated surgically | 5 | No |
| 102040. Preprocedural pulmonary hypertension (pulmonary pressure > or = systemic pressure): echo data | 5 | No |
| 102041. Preprocedural pulmonary hypertension (pulmonary pressure > or = systemic pressure): catheter data | 5 | No |
| 102045. Preprocedural pulmonary hypertension (pulmonary pressure more than or equal to systemic pressure): echo data, | 5 | No |
| 102046. Preprocedural pulmonary hypertension (pulmonary pressure more than or equal to systemic pressure): catheter data | 5 | No |
| 102202. Premature birth | 5 | No |
| 102203. Infant of diabetic mother | 5 | No |
| 102205. Premature birth 32-35 weeks | 5 | No |
| 102206. Premature birth < 32 weeks | 5 | No |
| 102304. Hereditary disorder associated with heart disease | 5 | No |
| 109024. Traumatic injury of tracheobronchial tree or lungs | 5 | No |
| 140101. Chromosomal anomaly | 5 | No |
| 140102. Trisomy 21: Down’s syndrome | 5 | No |
| 140103. Trisomy 18: Edwards' syndrome | 5 | No |
| 140104. Trisomy 13: Patau's syndrome | 5 | No |
| 140105. 45XO: Turner’s syndrome | 5 | No |
| 140121. 22q11 microdeletion | 5 | No |
| 140200. Syndrome-association with cardiac involvement | 5 | No |
| 140206. DiGeorge sequence | 5 | No |
| 140210. Friedreich’s ataxia | 5 | No |
| 140217. Marfan syndrome | 5 | No |
| 140219. Noonan syndrome | 5 | No |
| 140221. Pompe’s disease: glycogen storage disease type IIa | 5 | No |
| 140228. Tuberous sclerosis | 5 | No |
| 140230. Williams syndrome (infantile hypercalcaemia) | 5 | No |
| 140232. Fetal rubella syndrome | 5 | No |
| 140234. Duchenne’s muscular dystrophy | 5 | No |
| 140258. Muscular dystrophy | 5 | No |
| 140266. Alagille syndrome: arteriohepatic dysplasia | 5 | No |
| 140300. Non-cardiac abnormality associated with heart disease | 5 | No |
| 140304. Non-cardiothoracic-vascular abnormality | 5 | No |
| 140305. Psychomotor developmental delay | 5 | No |
| 140306. Cystic fibrosis | 5 | No |
| 140307. Congenital diaphragmatic hernia | 5 | No |
| 140308. Tracheo-oesophageal fistula | 5 | No |
| 140309. Gastro-oesophageal reflux disease (GORD) | 5 | No |
| 140310. Omphalocoele | 5 | No |
| 140311. Duodenal stenosis/atresia | 5 | No |
| 140323. Renal abnormality | 5 | No |
| 140328. Congenital coagulation disorder | 5 | No |
| 140329. Thoracic-mediastinal abnormality | 5 | No |
| 140333. Microcephaly | 5 | No |
| 140340. Brain abscess | 5 | No |
| 140342. Cerebrovascular accident (stroke), | 5 | No |
| 140344. Transient ischaemic attack: less than 24 hours (TIA), | 5 | No |
| 140347. Choanal atresia | 5 | No |
| 140349. Tracheobronchial malacia | 5 | No |
| 140352. Hypothyroidism | 5 | No |
| 140359. Obesity (Body Mass Index over 30) | 5 | No |
| 140372. Anoxic-ischaemic encephalopathy | 5 | No |
| 140375. Hyperthyroidism | 5 | No |
| 140390. Diabetes mellitus | 5 | No |
| 140391. Cerebral anomaly | 5 | No |
| 140392. Connective tissue disease | 5 | No |
| 140397. Reversible ischaemic neurologic deficit: 24-72 hours (RIND), | 5 | No |
| 140404. Pectus carinatum | 5 | No |
| 140405. Pectus excavatum | 5 | No |
| 140409. Kyphoscoliosis | 5 | No |
| 140412. Cleft lip or palate | 5 | No |
| 140414. Anterior chest wall (pectus) deformity | 5 | No |
| 140501. Maternal teratogen associated with congenital heart disease | 5 | No |
| 140601. Multiple congenital malformations | 5 | No |
| 152231. Residual pulmonary hypertension after relief of L to R shunt | 5 | No |
| 158210. Renal failure | 5 | No |
| 159503. Lymphoproliferative disease following transplantation | 5 | No |
| 160111. Empyema | 5 | No |
| 160200. Bronchial fistula | 5 | No |
| 160305. Lung disease | 5 | No |
| 160310. Asthma | 5 | No |
| 160513. Mediastinal disease | 5 | No |
| 160800. Acquired bronchial disease | 5 | No |
| 160900. Airway disease | 5 | No |
| 161001. Tracheal stenosis | 5 | No |
| 161009. Tracheal disease | 5 | No |
| 161300. Diaphragm disorder: acquired | 5 | No |
| 161320. Diaphragm paralysis | 5 | No |
| 162010. Oesophageal disorder | 5 | No |
| 163001. Respiratory failure | 5 | No |
| Q19182. - status: post lung(s) transplant | 5 | No |
| 010100. Normal heart | 5 | No |
| 010300. Usual atrial arrangement (atrial situs solitus) | 5 | No |
| 010310. Normal atrial arrangement (situs), AV & VA connections | 5 | No |
| 010500. Concordant VA connections | 5 | No |
| 020103. Laevocardia: heart predominantly in left hemithorax, | 5 | No |
| 050310. Intact atrial septum (no interatrial communication) | 5 | No |
| 071601. Spontaneous closure of VSD | 5 | No |
| 072001. Aneurysm of membranous septum | 5 | No |
| 072100. Intact ventricular septum | 5 | No |
| 101201. Innocent murmur | 5 | No |
| 102000. No pre-procedural risk factors | 5 | No |
| 141072. Liveborn | 5 | No |
| 100811. Postpericardiotomy syndrome | 5 | No |
| 101242. Chest pain | 5 | No |
| 101251. Angina pectoris | 5 | No |
| 101500. Neonatal disorder | 5 | No |
| 101700. Symptom-sign of heart disease | 5 | No |
| 101703. Cyanosis | 5 | No |
| 101705. Heart failure | 5 | No |
| 101709. Cardiac symptom without pathology | 5 | No |
| 101712. Cyanotic spells | 5 | No |
| 101713. Palpitations | 5 | No |
| 101750. Platypnoea-orthodeoxia syndrome | 5 | No |
| 101824. Postmyocardial infarction complication | 5 | No |
| 101852. Musculoskeletal chest pain | 5 | No |
| 101901. Dyslipidaemia | 5 | No |
| 102019. Preprocedural risk factor | 5 | No |
| 102207. Weight less than 2.5 kg | 5 | No |
| 102300. Hereditary-non-cardiac abnormality not apparent | 5 | No |
| 102301. Family history of congenital heart lesion | 5 | No |
| 102302. Maternal SLE | 5 | No |
| 102303. Family history of disorder with cardiac involvement | 5 | No |
| 102311. Family history of sudden death, | 5 | No |
| 103030. Pulmonary oedema | 5 | No |
| 104001. Syncope | 5 | No |
| 104030. Hypotension | 5 | No |
| 110021. Cardiac arrest | 5 | No |
| 110204. Sinus bradycardia | 5 | No |
| 110207. Sinus tachycardia | 5 | No |
| 110215. Vagal sinus bradycardia: bradycardia(s) of prematurity | 5 | No |
| 110321. Premature atrial beats (complexes-contractions) | 5 | No |
| 110412. Junctional ectopic tachycardia (His bundle): post-op | 5 | No |
| 110617. Postprocedural complete AV block | 5 | No |
| 110632. Procedure related complete AV block requiring temporary pacing | 5 | No |
| 110633. Procedure related complete AV block requiring permanent pacemaker system | 5 | No |
| 112000. ECG abnormality | 5 | No |
| 122341. Transluminal intracoronary echocardiography (IVUS) | 5 | No |
| 123203. Exploratory procedure to assess heart | 5 | No |
| 123331. Intraoperative death | 5 | No |
| 123333. Death within 30 days of procedure | 5 | No |
| 123334. Death unrelated to cardiac procedure | 5 | No |
| 124570. Fetal transluminal catheter procedure | 5 | No |
| 124571. Fetal balloon pulmonary valvotomy | 5 | No |
| 124573. Fetal balloon aortic valvotomy | 5 | No |
| 124575. Fetal pericardiocentesis: transcatheter | 5 | No |
| 124576. Fetal balloon atrial septostomy | 5 | No |
| 124590. Fetal procedure | 5 | No |
| 124591. Division of placental communicating vessels in twin-to-twin transfusion syndrome by laser coagulation | 5 | No |
| 130001. Clinical evaluation of patient | 5 | No |
| 130002. Diagnostic investigation | 5 | No |
| 130010. Electrocardiogram (ECG) | 5 | No |
| 130011. 24 hour electrocardiogram (ECG) recording | 5 | No |
| 130013. Exercise stress test | 5 | No |
| 130014. Insertable electrocardiogram (ECG) loop recorder (e.g. Reveal) implantation | 5 | No |
| 130015. Insertable electrocardiogram (ECG) loop recorder (e.g. Reveal) removal | 5 | No |
| 130016. Cardiopulmonary exercise stress test | 5 | No |
| 130017. Ambulatory electrocardiography (ECG) recording | 5 | No |
| 130021. Chest x-ray | 5 | No |
| 130023. Computerised tomographic scan of chest | 5 | No |
| 130024. Cardiovascular Magnetic Resonance Imaging (CMRI) | 5 | No |
| 130025. Pulmonary Function Tests | 5 | No |
| 130027. Tilt table testing | 5 | No |
| 130031. Myocardial perfusion scan | 5 | No |
| 130032. Non-cardiac computed tomographic angiography on cardiac patient, | 5 | No |
| 130033. Non-cardiovascular Magnetic Resonance Imaging on cardiac patient, | 5 | No |
| 130034. Diagnostic radiographic procedure on cardiac patient, | 5 | No |
| 130035. Therapeutic radiological procedure on cardiac patient, | 5 | No |
| 130036. Ambulatory blood pressure monitoring, | 5 | No |
| 130050. Arrhythmia provocation test | 5 | No |
| 130071. Post mortem examination of heart-lungs | 5 | No |
| 130100. Echocardiographic examination | 5 | No |
| 130101. Fetal echocardiographic examination | 5 | No |
| 130102. Transthoracic echocardiographic examination | 5 | No |
| 130103. Transoesophageal echocardiographic examination | 5 | No |
| 130104. Epicardial echocardiographic examination | 5 | No |
| 130124. Transluminal intracardiac echocardiographic examination | 5 | No |
| 130125. Transabdominal fetal echocardiographic examination | 5 | No |
| 130126. Transvaginal fetal echocardiographic examination | 5 | No |
| 130127. Intravascular ultrasound (IVUS) examination | 5 | No |
| 130189. Satisfactory fetal cardiac images | 5 | No |
| 130190. Inadequate fetal cardiac images | 5 | No |
| 140446. Migraine, | 5 | No |
| 140447. Diver's decompression sickness, | 5 | No |
| 140470. Smoking: tobacco use, | 5 | No |
| 140500. Maternal teratogen or disease potentially associated with congenital heart disease | 5 | No |
| 140540. Maternally derived fetal disease or syndrome associated with heart disease | 5 | No |
| 140541. Fetal infection | 5 | No |
| 141000. Fetal cardiac abnormality not detected | 5 | No |
| 141001. Fetal abnormality | 5 | No |
| 141002. Fetal hydrops | 5 | No |
| 141006. Fetal arrhythmia | 5 | No |
| 141007. Fetal unexplained right heart dominance | 5 | No |
| 141008. Fetal echogenic focus ('golf ball') | 5 | No |
| 141011. Twin-to-twin transfusion syndrome | 5 | No |
| 141012. Increased nuchal thickness | 5 | No |
| 141018. In utero death | 5 | No |
| 141025. Fetal echocardiographic abnormality | 5 | No |
| 141026. Fetal venous Doppler flow(s) abnormal for gestational age | 5 | No |
| 141027. Fetal progression of right ventricular outflow tract obstruction | 5 | No |
| 141028. Fetal progression of left ventricular outflow tract obstruction | 5 | No |
| 141034. Intrauterine growth restriction (retardation) | 5 | No |
| 141035. Spontaneous abortion-miscarriage (< 24 wks) | 5 | No |
| 141036. Stillbirth (> 24 wks) | 5 | No |
| 141037. Termination of pregnancy procedure | 5 | No |
| 141038. Terminated pregnancy (with respect to fetus) | 5 | No |
| 141045. Fetal unexplained left heart dominance | 5 | No |
| 141052. Fetal abnormal 4 chamber view | 5 | No |
| 141053. Fetal abnormal great arterial view | 5 | No |
| 141060. Fetal structural heart abnormality | 5 | No |
| 141073. Selective feticide in multiple pregnancy | 5 | No |
| 141090. Fetal failure of right ventricular growth | 5 | No |
| 141091. Fetal failure of left ventricular growth | 5 | No |
| 141095. Fetal cardiomegaly | 5 | No |
| 141098. Fetal heart failure | 5 | No |
| 141099. Fetal heart failure due to extracardiac disease | 5 | No |
| 141140. Fetal arterial Doppler flow(s) abnormal for gestational age | 5 | No |
| 141141. Fetal aortic arch flow abnormal for gestational age | 5 | No |
| 141142. Fetal reversal of aortic arch blood flow for gestational age | 5 | No |
| 141143. Fetal ductal arch flow abnormal for gestational age | 5 | No |
| 141144. Fetal reversal of ductal arch blood flow for gestational age | 5 | No |
| 141180. Fetal heart muscle abnormality | 5 | No |
| 141181. Fetal cardiomyopathy | 5 | No |
| 141187. Fetal ventricular dysfunction | 5 | No |
| 141220. Fetal fluid retention | 5 | No |
| 141352. Abnormal fetal Cardiovascular Profile Score (Huhta) | 5 | No |
| 150001. Cardiac arrest during procedure | 5 | No |
| 150002. Cardiac arrest following procedure | 5 | No |
| 150003. Postprocedural low cardiac output | 5 | No |
| 150005. Myocardial infarction following procedure | 5 | No |
| 150009. Postprocedural requirement for mechanical circulatory support | 5 | No |
| 150030. Postprocedural hypovolaemia | 5 | No |
| 150200. Postprocedural haemorrhage | 5 | No |
| 150203. Postprocedural coagulopathy | 5 | No |
| 150207. Postprocedural haemolysis | 5 | No |
| 150265. Postprocedural haemorrhage requiring reoperation | 5 | No |
| 150300. Median sternotomy complication | 5 | No |
| 150303. Infection of median sternotomy wound | 5 | No |
| 150308. Dehiscence of median sternotomy wound | 5 | No |
| 150315. Keloid-hypertrophic scar of median sternotomy wound | 5 | No |
| 150330. Lateral thoracotomy complication | 5 | No |
| 150332. Infection of lateral thoracotomy wound | 5 | No |
| 150350. Wound infection | 5 | No |
| 150351. Wound dehiscence | 5 | No |
| 150352. Mediastinitis | 5 | No |
| 150415. Postprocedural femoral vein complication | 5 | No |
| 150434. Postprocedural major vein complication | 5 | No |
| 152420. Postprocedural femoral arterial complication | 5 | No |
| 154139. Postprocedural coronary artery bypass graft (CABG) complication | 5 | No |
| 154306. Unplanned reoperation during current admission | 5 | No |
| 155000. Cardiac catheterisation complication | 5 | No |
| 155001. Intramyocardial injection of contrast medium | 5 | No |
| 155003. Perforation of cardiac chamber-vessel during cardiac catheterisation | 5 | No |
| 155011. Lost pulse after cardiac catheterisation | 5 | No |
| 155030. Equipment problem during cardiac catheterisation | 5 | No |
| 155037. Embolisation of catheter introduced device | 5 | No |
| 155040. Failed attempt to implant coil-device during transcatheter intervention | 5 | No |
| 155060. Complication involving device implantation | 5 | No |
| 155070. Complication involving stent | 5 | No |
| 155080. Fetal intervention complication | 5 | No |
| 155081. Preterm delivery immediately following fetal intervention | 5 | No |
| 155083. Failed fetal procedure | 5 | No |
| 155086. Fetal death following fetal cardiac procedure | 5 | No |
| 155100. Complication following arrhythmia related procedure | 5 | No |
| 155702. Extracorporeal Membrane Oxygenation (ECMO) circuit complication | 5 | No |
| 155703. Ventricular assist device complication | 5 | No |
| 155721. Intraaortic balloon pump (IABP) complication | 5 | No |
| 155801. Complication related to echocardiographic procedure | 5 | No |
| 155900. Medication related complication or error | 5 | No |
| 156002. Arrhythmia following procedure | 5 | No |
| 156738. Wound related complication | 5 | No |
| 157700. Cardiopulmonary bypass complication | 5 | No |
| 158000. General systemic complication of cardiac procedure | 5 | No |
| 158001. Postprocedural metabolic derangement | 5 | No |
| 158005. Postprocedural septicaemia | 5 | No |
| 158006. Capillary leak syndrome | 5 | No |
| 158015. Postprocedural acidosis | 5 | No |
| 158016. Multiple organ dysfunction syndrome (MODS) | 5 | No |
| 158019. Systemic inflammatory response syndrome (SIRS) | 5 | No |
| 158020. Respiratory complication after cardiac procedure | 5 | No |
| 158021. Postprocedural pulmonary infection | 5 | No |
| 158022. Postprocedural pulmonary hypertensive crises | 5 | No |
| 158029. Postprocedural Acute Respiratory Distress Syndrome (ARDS) | 5 | No |
| 158031. Postprocedural lung collapse (atelectasis) | 5 | No |
| 158032. Postprocedural requirement for mechanical respiratory support > 7 days | 5 | No |
| 158033. Postprocedural requirement for reintubation | 5 | No |
| 158050. Postprocedural pleural effusion | 5 | No |
| 158051. Postprocedural right pleural effusion | 5 | No |
| 158052. Postprocedural left pleural effusion | 5 | No |
| 158055. Postprocedural chylothorax | 5 | No |
| 158056. Postprocedural haemothorax | 5 | No |
| 158061. Pleural effusion requiring drainage | 5 | No |
| 158062. Postprocedural pneumothorax | 5 | No |
| 158070. Postprocedural complication involving tracheo-bronchial tree | 5 | No |
| 158086. Postprocedural requirement for tracheostomy | 5 | No |
| 158087. Postprocedural bronchial compression | 5 | No |
| 158090. Intraprocedural phrenic nerve injury (paralysed diaphragm) | 5 | No |
| 158093. Intraprocedural recurrent laryngeal nerve injury (palsy) | 5 | No |
| 158094. Postprocedural Horner’s syndrome | 5 | No |
| 158200. Postprocedural renal failure | 5 | No |
| 158206. Renal failure requiring temporary dialysis | 5 | No |
| 158207. Renal failure requiring permanent dialysis | 5 | No |
| 158221. Postprocedural gastrointestinal bleeding | 5 | No |
| 158223. Postprocedural inability to sustain gastric feeding | 5 | No |
| 158228. Postprocedural intestinal obstruction | 5 | No |
| 158229. Postprocedural peritonitis | 5 | No |
| 158230. Postprocedural necrotising enterocolitis | 5 | No |
| 158232. Pseudomembranous colitis | 5 | No |
| 158233. Postprocedural protein losing enteropathy | 5 | No |
| 158238. Postprocedural feeding difficulties | 5 | No |
| 158243. Postprocedural hepatic impairment | 5 | No |
| 158247. Postprocedural acute pancreatitis | 5 | No |
| 158250. Neurological complication after cardiac procedure | 5 | No |
| 158251. Postprocedural generalised seizures | 5 | No |
| 158253. Postprocedural temporary neurological impairment | 5 | No |
| 158257. Postprocedural permanent neurological impairment | 5 | No |
| 158264. Postprocedural brain death | 5 | No |
| 158266. Postprocedural cerebral abscess | 5 | No |
| 158267. Postprocedural new onset seizures | 5 | No |
| 158268. Postprocedural neurological impairment persisting at discharge | 5 | No |
| 158281. Postprocedural cerebral abnormality on imaging | 5 | No |
| 158800. Vascular line (access) related complication | 5 | No |
| 159001. Postprocedural complication | 5 | No |
| 159003. No postprocedural complications | 5 | No |
| 159014. Procedure related complication | 5 | No |
| 159020. Complication during period of anaesthetic care | 5 | No |
| 159500. Complication after heart or lung transplant | 5 | No |
| 159564. Post-lung transplant obliterative bronchiolitis | 5 | No |
| 159566. Lung disease in lung transplant rejection | 5 | No |
| 160101. Pneumothorax | 5 | No |
| 160104. Pleural effusion | 5 | No |
| 160107. Chylothorax | 5 | No |
| 160121. Pleural disease: benign | 5 | No |
| 160122. Pleural disease: malignant | 5 | No |
| 160301. Lung disease: benign | 5 | No |
| 160302. Lower respiratory tract infection | 5 | No |
| 160321. Lung disease: malignant | 5 | No |
| 160511. Mediastinal disease: benign | 5 | No |
| 160512. Mediastinal disease: malignant | 5 | No |
| 161509. Diaphragm disease | 5 | No |
| 162001. Oesophageal disease: benign | 5 | No |
| 162002. Oesophageal disease: malignant | 5 | No |
| 165020. Complication following respiratory tract stent implantation | 5 | No |
| 170001. Medical therapy for cardiovascular disease | 5 | No |
| 171002. Medical therapy for endocarditis | 5 | No |
| Q19051. - status: post-procedure | 5 | No |
| Q19067. - diagnosis uncertain | 5 | No |
| 010161. Aneurysm | 6 | No |
| 040006. Systemic venovenous collateral(s), | 6 | No |
| 070001. Ventricular dyssynchrony | 6 | No |
| 070110. Arrhythmogenic right ventricular cardiomyopathy | 6 | No |
| 070111. Right ventricular dysfunction | 6 | No |
| 070610. Left ventricular dysfunction | 6 | No |
| 070850. Ventricular myocardial noncompaction cardiomyopathy | 6 | No |
| 090515. Pulmonary valvar atresia: acquired | 6 | No |
| 090591. Pulmonary regurgitation | 6 | No |
| 090700. Pulmonary trunk (MPA) abnormality | 6 | No |
| 091000. Pulmonary arterial abnormality | 6 | No |
| 091044. Pulmonary arterial aneurysm | 6 | No |
| 091605. Ascending aorta dilation associated with Marfan syndrome | 6 | No |
| 091609. Ascending aorta dilation | 6 | No |
| 091613. Aortic root dilation | 6 | No |
| 092816. Descending aorta dilation | 6 | No |
| 094601. Coronary arterial aneurysm(s) | 6 | No |
| 100301. Heart tumour | 6 | No |
| 100501. Acute rheumatic fever | 6 | No |
| 100521. Rheumatic fever with cardiac involvement | 6 | No |
| 100530. Rheumatic valvar disease | 6 | No |
| 100531. Rheumatic mitral valvar disease | 6 | No |
| 100533. Rheumatic aortic valvar disease | 6 | No |
| 100600. Endocarditis | 6 | No |
| 100601. Infective endocarditis | 6 | No |
| 100620. Heart abscess | 6 | No |
| 100641. Bacterial endocarditis | 6 | No |
| 100664. Postprocedural endocarditis | 6 | No |
| 100701. Infectious myocarditis | 6 | No |
| 100703. Viral myocarditis | 6 | No |
| 100705. Drug induced heart muscle disease | 6 | No |
| 100708. Trypanosomal myocarditis (Chagas' disease) | 6 | No |
| 100740. Myocardial failure in end stage congenital heart disease | 6 | No |
| 100742. Heart muscle disease in cardiac rejection | 6 | No |
| 100761. Nutritional heart muscle disease | 6 | No |
| 100771. Heart muscle disease in infant of diabetic mother | 6 | No |
| 100781. Heart muscle disease in collagen vascular/ connective tissue disorder | 6 | No |
| 100800. Pericarditis | 6 | No |
| 100801. Infectious pericarditis | 6 | No |
| 100803. Viral pericarditis | 6 | No |
| 100804. Bacterial pericarditis | 6 | No |
| 100809. Constrictive pericarditis | 6 | No |
| 100813. Cardiac tamponade | 6 | No |
| 100815. Chylopericardium | 6 | No |
| 100829. Pericardial abnormality: acquired | 6 | No |
| 100831. Pericardial effusion | 6 | No |
| 100901. Kawasaki disease | 6 | No |
| 100902. Kawasaki disease with aneurysm(s) or dilated coronary vessels | 6 | No |
| 100908. Kawasaki disease without cardiac involvement | 6 | No |
| 100910. Acquired coronary arterial disease | 6 | No |
| 100930. Ischaemic heart disease | 6 | No |
| 101001. Cardiomyopathy | 6 | No |
| 101011. Idiopathic restrictive cardiomyopathy | 6 | No |
| 101012. Endocardial fibroelastosis | 6 | No |
| 101013. Infiltrative cardiomyopathy | 6 | No |
| 101020. Hypertrophic cardiomyopathy | 6 | No |
| 101025. Dilated cardiomyopathy | 6 | No |
| 101239. Failure to thrive | 6 | No |
| 101301. Pulmonary arterial hypertension | 6 | No |
| 101302. Primary pulmonary hypertension | 6 | No |
| 101306. Pulmonary vascular disease | 6 | No |
| 101308. Irreversible pulmonary vascular disease due to congenital heart disease (Eisenmenger Syndrome) | 6 | No |
| 101320. Secondary pulmonary hypertension | 6 | No |
| 101321. Pulmonary hypertension due to left to right shunt | 6 | No |
| 101350. Pulmonary arterial disease: acquired | 6 | No |
| 101351. Pulmonary embolism | 6 | No |
| 101368. Right pulmonary arterial stenosis: acquired | 6 | No |
| 101369. Left pulmonary arterial stenosis: acquired | 6 | No |
| 101401. Systemic hypertension | 6 | No |
| 101404. Systemic hypertension due to aortic arch obstruction | 6 | No |
| 101440. Ascending aorta dilation: acquired | 6 | No |
| 101442. Ascending aorta aneurysm | 6 | No |
| 101443. Descending aorta aneurysm | 6 | No |
| 101450. Aortic aneurysm | 6 | No |
| 101451. Aortic dissection | 6 | No |
| 101452. Ascending aorta dissection & propagation beyond arch (DeBakey type I) | 6 | No |
| 101453. Ascending aorta dissection not beyond arch (DeBakey type II/ Stanford type A) | 6 | No |
| 101470. Abnormality of aorta: acquired | 6 | No |
| 101472. Recoarctation of aorta | 6 | No |
| 101477. Supravalvar aortic stenosis: acquired | 6 | No |
| 101480. Arterial duct (ductus arteriosus) abnormality: acquired | 6 | No |
| 101495. Ascending aortopathy associated with conotruncal malformations, | 6 | No |
| 101510. Transient myocardial ischaemia | 6 | No |
| 101600. Right ventricular abnormality: acquired | 6 | No |
| 101608. Right ventricular-congestive heart failure | 6 | No |
| 101616. Right ventricular outflow tract obstruction: acquired | 6 | No |
| 101640. Left ventricular abnormality: acquired | 6 | No |
| 101646. Recurrent left ventricular outflow tract obstruction | 6 | No |
| 101647. Left ventricular failure | 6 | No |
| 101660. Abnormality associated with ventricular septum: acquired | 6 | No |
| 101662. Post-myocardial infarct VSD | 6 | No |
| 101681. Narrowing of constructed intraventricular tunnel: acquired | 6 | No |
| 101682. Subaortic stenosis in complex heart disease: acquired | 6 | No |
| 101683. Subpulmonary stenosis in complex heart disease: acquired | 6 | No |
| 101686. Subaortic stenosis: acquired | 6 | No |
| 101688. Subpulmonary stenosis: acquired | 6 | No |
| 101723. Shock | 6 | No |
| 101740. Atrial septum abnormality: acquired | 6 | No |
| 101800. Myocardial infarction | 6 | No |
| 101801. Acute myocardial infarction | 6 | No |
| 102034. Preprocedural myocardial dysfunction | 6 | No |
| 102400. Pulmonary venous abnormality: acquired | 6 | No |
| 103000. Systemic vein abnormality: acquired | 6 | No |
| 103009. Systemic vein obstruction | 6 | No |
| 103101. Superior caval vein (SVC) abnormality: acquired | 6 | No |
| 103121. Inferior caval vein (IVC) abnormality - acquired | 6 | No |
| 103200. Heart valvar abnormality: acquired | 6 | No |
| 103201. Tricuspid valvar abnormality: acquired | 6 | No |
| 103300. Prosthetic valve failure | 6 | No |
| 103301. Mitral valvar abnormality: acquired | 6 | No |
| 103302. Mitral stenosis: acquired | 6 | No |
| 103303. Mitral valvar stenosis: recurrent | 6 | No |
| 103304. Mitral regurgitation: acquired | 6 | No |
| 103306. Mitral regurgitation: recurrent | 6 | No |
| 103444. Left AV valvar regurgitation: acquired | 6 | No |
| 103460. AV valvar abnormality in AVSD: acquired | 6 | No |
| 103501. Pulmonary valvar abnormality: acquired | 6 | No |
| 103502. Pulmonary valvar stenosis: acquired | 6 | No |
| 103503. Pulmonary valvar stenosis: recurrent | 6 | No |
| 103504. Pulmonary regurgitation: acquired | 6 | No |
| 103601. Aortic valvar abnormality: acquired | 6 | No |
| 103602. Aortic valvar stenosis: acquired | 6 | No |
| 103603. Aortic valvar stenosis: recurrent | 6 | No |
| 103604. Aortic regurgitation: acquired | 6 | No |
| 103606. Aortic regurgitation: recurrent | 6 | No |
| 103701. Truncal valvar abnormality: acquired | 6 | No |
| 105101. Common arterial trunk (truncus) abnormality: acquired | 6 | No |
| 109001. Traumatic injury of heart | 6 | No |
| 111100. Pacemaker dysfunction-complication necessitating replacement | 6 | No |
| 111101. Pacemaker dysfunction-complication, | 6 | No |
| 111103. Pacemaker battery exhaustion: end of life (EOL) | 6 | No |
| 111117. Pacemaker-ICD loss of capture | 6 | No |
| 111121. Pacemaker (AV dyssychrony) syndrome | 6 | No |
| 111140. Pacemaker lead dysfunction-complication | 6 | No |
| 111159. Pacemaker generator site local complication | 6 | No |
| 111160. Implantable cardioverter & defibrillator (ICD) dysfunction-complication | 6 | No |
| 111170. Insertable ECG loop recorder complication | 6 | No |
| 111180. Insertable electrocardiographic (ECG) loop recorder complication, | 6 | No |
| 150401. Postprocedural superior caval vein (SVC) complication | 6 | No |
| 150405. Postprocedural inferior caval vein (IVC) complication | 6 | No |
| 150501. Postprocedural pulmonary vein complication | 6 | No |
| 150503. Pulmonary vein obstruction | 6 | No |
| 151010. Right atrial abnormality: acquired | 6 | No |
| 151011. Postprocedural right atrial complication | 6 | No |
| 151013. Obstruction of right atrial conduit (TCPC) | 6 | No |
| 151020. Left atrial abnormality: acquired | 6 | No |
| 151021. Postprocedural left atrial complication | 6 | No |
| 151061. Postprocedural atrial septum complication | 6 | No |
| 151063. Residual interatrial communication ('ASD') | 6 | No |
| 151066. Ineffective balloon atrial septostomy | 6 | No |
| 151100. Postprocedural tricuspid valvar complication | 6 | No |
| 151103. Residual tricuspid regurgitation | 6 | No |
| 151108. Tricuspid valvar prosthesis complication | 6 | No |
| 151200. Postprocedural mitral valvar complication | 6 | No |
| 151201. Residual mitral valvar stenosis | 6 | No |
| 151203. Residual mitral regurgitation | 6 | No |
| 151209. Mitral valvar prosthesis complication | 6 | No |
| 151302. Residual common AV valvar regurgitation | 6 | No |
| 151400. Postprocedural right AV valvar complication | 6 | No |
| 151500. Postprocedural left AV valvar complication | 6 | No |
| 151600. Postprocedural AV septal defect complication | 6 | No |
| 151602. Residual ventricular component of AV septal defect | 6 | No |
| 152001. Postprocedural right ventricular complication | 6 | No |
| 152021. Postprocedural right ventricular outflow tract complication | 6 | No |
| 152023. Residual right ventricular outflow tract obstruction | 6 | No |
| 152025. Aneurysm of right ventricular outflow tract patch | 6 | No |
| 152075. Residual subaortic stenosis in complex heart disease | 6 | No |
| 152076. Residual subpulmonary stenosis in complex heart disease | 6 | No |
| 152101. Postprocedural left ventricular complication | 6 | No |
| 152121. Postprocedural left ventricular outflow tract complication | 6 | No |
| 152122. Residual left ventricular outflow tract obstruction | 6 | No |
| 152202. Residual VSD | 6 | No |
| 152400. Postprocedural systemic arterial complication | 6 | No |
| 152503. Residual truncal regurgitation | 6 | No |
| 152531. Postprocedural common arterial trunk complication | 6 | No |
| 153000. Postprocedural pulmonary valvar complication | 6 | No |
| 153001. Residual pulmonary valvar stenosis | 6 | No |
| 153003. Residual pulmonary regurgitation | 6 | No |
| 153008. Pulmonary valvar prosthesis complication | 6 | No |
| 153201. Postprocedural pulmonary trunk complication | 6 | No |
| 153221. Postprocedural right pulmonary artery complication | 6 | No |
| 153223. Residual right pulmonary artery stenosis | 6 | No |
| 153241. Postprocedural left pulmonary artery complication | 6 | No |
| 153243. Residual left pulmonary artery stenosis | 6 | No |
| 153500. Postprocedural aortic valvar complication | 6 | No |
| 153501. Residual aortic valvar stenosis | 6 | No |
| 153503. Residual aortic regurgitation | 6 | No |
| 153508. Aortic valvar prosthesis complication | 6 | No |
| 153601. Postprocedural ascending aorta complication | 6 | No |
| 153701. Postprocedural descending aorta complication | 6 | No |
| 153705. Residual aortic coarctation | 6 | No |
| 153707. Postprocedural aneurysm of aorta at coarctation site | 6 | No |
| 153773. Postprocedural aortic complication | 6 | No |
| 153901. Postprocedural arterial duct complication | 6 | No |
| 153902. Residual arterial duct (PDA) patency | 6 | No |
| 153950. Postprocedural systemic-to-pulmonary collateral artery complication | 6 | No |
| 154100. Postprocedural coronary arterial complication | 6 | No |
| 154113. Cardiac transplant associated coronary allograft vasculopathy | 6 | No |
| 155500. Cardiac conduit complication | 6 | No |
| 155516. Cardiac conduit failure | 6 | No |
| 155524. Pulmonary autograft failure, | 6 | No |
| 155600. Systemic-to-pulmonary arterial shunt complication | 6 | No |
| 155601. Systemic-to-pulmonary arterial shunt partial obstruction | 6 | No |
| 155602. Systemic-to-pulmonary arterial shunt complete obstruction | 6 | No |
| 155621. Systemic-to-pulmonary arterial shunt failure, | 6 | No |
| 158300. Pericardial effusion requiring drainage | 6 | No |
| 159060. Failed' Fontan type circulation | 6 | Yes |
| Q19180. - status: post heart transplant | 6 | No |
| Q19181. - status: post heart & lung(s) transplant | 6 | No |
| 050601. Common atrium (virtual absence of atrial septum) | 7 | No |
| 060501. AVSD AV valvar abnormality | 7 | No |
| 060506. AVSD AV valvar regurgitation | 7 | No |
| 060600. Atrioventricular septal defect | 7 | No |
| 060601. AVSD: isolated atrial component (primum ASD)(partial) | 7 | No |
| 060608. AVSD: isolated ventricular component | 7 | No |
| 060609. AVSD: atrial & ventricular components with common AV orifice (complete) | 7 | No |
| 060610. AVSD: atrial & (restrictive) ventricular components + separate AV valves ('intermediate') | 7 | No |
| 010101. Tetralogy of Fallot | 7 | No |
| 010117. Double outlet right ventricle: Fallot type (subaortic or doubly committed VSD & pulmonary stenosis) | 7 | No |
| 091501. Aortic valvar stenosis: congenital | 8 | No |
| 091512. Eccentric opening of tricuspid aortic valve | 8 | No |
| 091513. Aortic valvar stenosis | 8 | No |
| 091592. Aortic stenosis | 8 | No |
| 050202. Supravalvar mitral ring | 8 | No |
| 060200. Mitral valvar abnormality | 8 | No |
| 060207. Mitral valvar stenosis: congenital | 8 | No |
| 060212. Mitral subvalvar apparatus abnormality | 8 | No |
| 060213. Mitral subvalvar stenosis | 8 | No |
| 060225. Mitral regurgitation: congenital | 8 | No |
| 060235. Mitral valvar prolapse | 8 | No |
| 060236. True cleft of mitral leaflet (without AVSD) | 8 | No |
| 060256. Parachute malformation of mitral valve | 8 | No |
| 060291. Mitral regurgitation | 8 | No |
| 060292. Mitral stenosis | 8 | No |
| 060293. Mitral valve stenosis | 8 | No |
| 010139. Cardiac abnormality | 8 | No |
| 010160. Vascular abnormality | 8 | No |
| 010306. Abnormal atrial arrangement | 8 | No |
| 010309. AV and-or VA connections abnormal | 8 | Yes |
| 010510. Concordant VA connections with parallel great arteries (anatomically corrected malposition) | 8 | No |
| 020102. Dextrocardia: heart predominantly in right hemithorax | 8 | No |
| 020104. Midline heart (mesocardia), | 8 | No |
| 020109. Position-orientation of heart abnormal | 8 | No |
| 020704. Abnormal position or relationship of great vessels | 8 | No |
| 030103. Total mirror imagery (situs inversus) | 8 | No |
| 030104. Right isomerism ('asplenia') | 8 | No |
| 030105. Left isomerism ('polysplenia') | 8 | No |
| 040100. Superior caval vein (SVC) abnormality | 8 | No |
| 040101. Left superior caval vein (SVC) persisting to coronary sinus | 8 | No |
| 040200. Hepatic vein abnormality | 8 | No |
| 040300. Inferior caval vein (IVC) abnormality | 8 | No |
| 040310. Inferior caval vein (IVC) interruption (absent suprarenal segment) with azygos continuation | 8 | No |
| 040400. Coronary sinus abnormality | 8 | No |
| 040500. Systemic vein abnormality: congenital | 8 | No |
| 040800. Pulmonary vein abnormality | 8 | No |
| 050100. Right atrial abnormality | 8 | No |
| 050200. Left atrial abnormality | 8 | No |
| 050300. Atrial septum abnormality | 8 | No |
| 050301. Patent foramen ovale (PFO) | 8 | No |
| 060109. Straddling tricuspid valve | 8 | No |
| 060209. Straddling mitral valve | 8 | No |
| 060598. Deficient mural-lateral leaflet of left ventricular component of common atrioventricular valve (left atrioventricular vale) | 8 | No |
| 070100. Right ventricular abnormality | 8 | No |
| 070114. Right ventricular aneurysm | 8 | No |
| 070200. Right ventricular hypoplasia | 8 | No |
| 070301. Double chambered right ventricle | 8 | No |
| 070501. Right ventricular outflow tract obstruction | 8 | No |
| 070600. Left ventricular abnormality | 8 | No |
| 070613. Left ventricular aneurysm | 8 | No |
| 070700. Left ventricular hypoplasia | 8 | No |
| 070901. Left ventricular outflow tract obstruction | 8 | No |
| 070931. Aortic abnormality | 8 | No |
| 072000. Ventricular septal abnormality | 8 | No |
| 090522. Pulmonary regurgitation: congenital | 8 | No |
| 090711. Pulmonary trunk hypoplasia | 8 | No |
| 090713. Supravalvar pulmonary trunk stenosis | 8 | No |
| 090906. Pulmonary arterial sling | 8 | No |
| 090908. Pulmonary artery from ascending aorta (hemitruncus) | 8 | No |
| 091001. Pulmonary arterial stenosis | 8 | No |
| 091006. Peripheral pulmonary arterial stenoses: at-beyond hilar bifurcation | 8 | No |
| 091007. Central pulmonary arterial stenosis: proximal to hilar bifurcation | 8 | No |
| 091010. Discontinuous (non-confluent) pulmonary arteries | 8 | No |
| 091011. Pulmonary arterial hypoplasia | 8 | No |
| 091025. Right pulmonary arterial stenosis | 8 | No |
| 091026. Left pulmonary arterial stenosis | 8 | No |
| 091500. Aortic valvar abnormality | 8 | No |
| 091522. Bicuspid aortic valve | 8 | No |
| 091602. Ascending aorta hypoplasia | 8 | No |
| 091610. Ascending aorta abnormality | 8 | No |
| 091701. Aorto-ventricular tunnel | 8 | No |
| 091702. Aorto: left ventricular tunnel | 8 | No |
| 091801. Aortic sinus of Valsalva aneurysm | 8 | No |
| 091901. Arteriovenous fistula (malformation) | 8 | No |
| 091905. Pulmonary arteriovenous fistula (malformation) | 8 | No |
| 092020. Distal systemic arterial abnormality | 8 | No |
| 092800. Aortic arch abnormality | 8 | No |
| 092809. Double aortic arch | 8 | No |
| 092815. Right aortic arch | 8 | No |
| 092916. Descending-abdominal aorta hypoplasia (middle aortic syndrome) | 8 | No |
| 093000. Aortic arch branch abnormality | 8 | No |
| 093002. Aberrant origin right subclavian artery | 8 | No |
| 093004. Aberrant origin left subclavian artery | 8 | No |
| 094200. Coronary artery: anomalous aortic origin or course | 8 | No |
| 094220. Anomalous aortic origin of cornoary artery (AAOCA) | 8 | No |
| 094305. Intramural proximal coronary arterial course | 8 | No |
| 094318. Aberrant course of coronary artery: across right ventricular outflow tract | 8 | No |
| 094501. Coronary fistula | 8 | No |
| 094511. Coronary fistulas from RV ('sinusoidal') | 8 | No |
| 094600. Coronary arterial abnormality | 8 | No |
| 094606. Right ventricle dependent coronary circulation | 8 | No |
| 100100. Pericardial abnormality | 8 | No |
| 010102. Transposition of great arteries (concordant AV & discordant VA connections) & IVS | 9 | No |
| 092901. Aortic coarctation | 10 | No |
| 092911. Aortic arch hypoplasia (tubular) | 10 | No |
| 070530. Subpulmonary stenosis | 10 | No |
| 090500. Pulmonary valvar abnormality | 10 | No |
| 090501. Pulmonary valvar stenosis | 10 | No |
| 090504. Pulmonary valvar stenosis: congenital | 10 | No |
| 090592. Pulmonary stenosis | 10 | No |
| 070900. Subaortic stenosis | 11 | No |
| 070903. Subaortic stenosis due to fibromuscular shelf | 11 | No |
| 091507. Aortic regurgitation: congenital | 11 | No |
| 091530. Aortic valvar prolapse | 11 | No |
| 091591. Aortic regurgitation | 11 | No |
| 071000. VSD | 11 | No |
| 071001. Perimembranous VSD | 11 | No |
| 071012. VSD + malaligned outlet septum | 11 | No |
| 071101. Muscular VSD | 11 | No |
| 071200. Subarterial VSD | 11 | No |
| 071201. Doubly committed subarterial VSD | 11 | No |
| 071402. Communication between left ventricle + right atrium (Gerbode defect) | 11 | No |
| 071405. Inlet VSD | 11 | No |
| 071501. Tiny VSD (Maladie de Roger) | 11 | No |
| 071504. Multiple VSDs | 11 | No |
| 071505. Single VSD | 11 | No |
| 040701. Partially anomalous pulmonary venous connection(s) | 11 | No |
| 050401. Interatrial communication ('ASD') | 11 | No |
| 050402. Atrial septal defect (ASD) within oval fossa (secundum) | 11 | No |
| 050403. Spontaneous closure of atrial septal defect (ASD) within oval fossa (secundum) | 11 | No |
| 050500. Sinus venosus defect (ASD) | 11 | No |
| 050503. Interatrial communication (ASD) through coronary sinus orifice | 11 | No |
| 110000. Arrhythmia | 11 | No |
| 110011. Sudden Arrhythmic Death Syndrome (SADS) | 11 | No |
| 110100. Supraventricular tachycardia | 11 | No |
| 110101. Supraventricular rhythm disturbance | 11 | No |
| 110203. Sinus node dysfunction (including sick sinus) | 11 | No |
| 110305. Paroxysmal atrial tachycardia | 11 | No |
| 110307. Atrial flutter | 11 | No |
| 110308. Atrial fibrillation | 11 | No |
| 110312. Focal atrial tachycardia: ectopic (automatic) | 11 | No |
| 110313. Macro-reentrant atrial tachycardia (including atrial flutter) | 11 | No |
| 110315. Multifocal atrial tachycardia (chaotic) | 11 | No |
| 110366. Cavotricuspid isthmus dependent reentry atrial tachycardia: atrial flutter | 11 | No |
| 110367. Non-cavotricuspid isthmus dependent atrial tachycardia | 11 | No |
| 110400. Rhythm disturbance at level of AV junction | 11 | No |
| 110407. AV junctional (nodal) tachycardia | 11 | No |
| 110411. AV nodal reentry tachycardia (AVNRT) | 11 | No |
| 110442. Junctional ectopic tachycardia (His bundle) | 11 | No |
| 110500. Ventricular rhythm disturbance | 11 | No |
| 110506. Ventricular tachycardia | 11 | No |
| 110509. Ventricular flutter | 11 | No |
| 110510. Ventricular fibrillation | 11 | No |
| 110517. Catecholaminergic polymorphic ventricular tachycardia, | 11 | No |
| 110521. Premature ventricular beats (complexes-contractions) | 11 | No |
| 110544. Brugada syndrome (ventricular tachycardia with anterior raised ST), | 11 | No |
| 110550. Non-sustained ventricular tachycardia | 11 | No |
| 110556. Focal ventricular tachycardia | 11 | No |
| 110557. Macro-reentrant ventricular tachycardia | 11 | No |
| 110584. Idiopathic fascicular ventricular tachycardia | 11 | No |
| 110600. Conduction disturbance | 11 | No |
| 110601. Sinoatrial block | 11 | No |
| 110602. 1st degree AV block | 11 | No |
| 110603. 2nd degree AV block | 11 | No |
| 110607. Complete AV block (3rd degree) | 11 | No |
| 110610. Acquired complete AV block | 11 | No |
| 110616. Congenital complete heart block | 11 | No |
| 110623. Complete R bundle branch block | 11 | No |
| 110624. Complete L bundle branch block | 11 | No |
| 110635. Preprocedural complete AV block | 11 | No |
| 110701. AV reciprocating (reentry) tachycardia: manifest preexcitation in sinus rhythm (Wolff Parkinson White) | 11 | No |
| 110706. Accessory pathway: retrograde conduction only (concealed: no preexcitation sinus rhythm) | 11 | No |
| 110711. Manifest accessory pathway | 11 | No |
| 110714. Permanent junctional reciprocating tachycardia (PJRT) | 11 | No |
| 110722. AV reciprocating (reentry) tachycardia: orthodromic | 11 | No |
| 110723. AV reentry (reciprocating) tachycardia: antidromic (typically wide QRS) | 11 | No |
| 110726. Multiple accessory pathways | 11 | No |
| 110728. AV reentry (reciprocating) tachycardia: orthodromic & antidromic | 11 | No |
| 110729. AV reciprocating (reentry) tachycardia (accessory pathway mediated) | 11 | No |
| 111200. Ion channelopathy | 11 | No |
| 111201. Prolonged QT interval | 11 | No |
| 111229. Long QT syndrome | 11 | No |
| 112300. Morphological abnormality of conduction system | 11 | No |

**codes taken form the European Paediatric Cardiac Code Short List OPCSV4.7 -1 April 2015*

### Supplementary Table S3: PRAIS 2 model specific procedure groups used for adjusting the case mix

| **Allowed NCHDA Specific Procedure*** | **Specific Procedure Grouping used in Model** |
| --- | --- |
| HLHS Hybrid Approach | Procedure grouping 1 |
| Norwood procedure (Stage 1) | Procedure grouping 1 |
| Arterial switch + aortic arch obstruction repair  (with-without VSD closure) | Procedure grouping 2 |
| Interrupted aortic arch repair | Procedure grouping 2 |
| TAPVC Repair + Arterial Shunt | Procedure grouping 2 |
| TAPVc Repair + Arterial Shunt | Procedure grouping 2 |
| Truncus and interruption repair | Procedure grouping 2 |
| Truncus arteriosus repair | Procedure grouping 2 |
| Arterial shunt | Procedure grouping 3 |
| Arterial switch + VSD closure | Procedure grouping 4 |
| Isolated Pulmonary artery band | Procedure grouping 4 |
| Repair of total anomalous pulmonary venous connection | Procedure grouping 4 |
| PDA ligation (surgical) | Procedure grouping 5 |
| Anomalous coronary artery repair | Procedure grouping 6 |
| Aortopulmonary window repair | Procedure grouping 6 |
| Arterial switch (for isolated transposition) | Procedure grouping 6 |
| Isolated coarctation/ hypoplastic aortic arch repair | Procedure grouping 6 |
| Mitral valve replacement | Procedure grouping 7 |
| Pulmonary atresia VSD repair | Procedure grouping 7 |
| Pulmonary vein stenosis procedure | Procedure grouping 7 |
| Ross-Konno procedure | Procedure grouping 7 |
| Senning or Mustard procedure | Procedure grouping 7 |
| Tetralogy with absent pulmonary valve repair | Procedure grouping 7 |
| Unifocalisation procedure (with/without shunt) | Procedure grouping 7 |
| Aortic root replacement (not Ross) | Procedure grouping 8 |
| Aortic valve repair | Procedure grouping 8 |
| Cardiac conduit replacement | Procedure grouping 8 |
| Heart Transplant | Procedure grouping 8 |
| Isolated RV to PA conduit construction | Procedure grouping 8 |
| Pulmonary valve replacement | Procedure grouping 8 |
| Tricupid valve repair | Procedure grouping 8 |
| Tricuspid valve repair | Procedure grouping 8 |
| Tricuspid valve replacement | Procedure grouping 8 |
| Atrioventricular septal defect and tetralogy repair | Procedure grouping 9 |
| Atrioventricular septal defect and tetralogy repair | Procedure grouping 9 |
| Cor triatriatum repair | Procedure grouping 9 |
| Multiple VSD Closure | Procedure grouping 9 |
| Rastelli - REV procedure | Procedure grouping 9 |
| Supravalvar aortic stenosis repair | Procedure grouping 9 |
| Bidirectional cavopulmonary shunt | Procedure grouping 10 |
| Atrioventricular septal defect (complete) repair | Procedure grouping 11 |
| Fontan procedure | Procedure grouping 12 |
| Aortic valve replacement – Ross | Procedure grouping 13 |
| Mitral valve repair | Procedure grouping 13 |
| Sinus Venosus ASD and-or PAPVC repair | Procedure grouping 13 |
| Subvalvar aortic stenosis repair | Procedure grouping 13 |
| Atrioventricular septal defect (partial) repair | Procedure grouping 14 |
| Tetralogy and Fallot-type DORV repair | Procedure grouping 14 |
| Vascular ring procedure | Procedure grouping 14 |
| Aortic Valve Replacement - non Ross | Procedure grouping 15 |
| ASD repair | Procedure grouping 15 |
| VSD Repair | Procedure grouping 15 |
| No Specific Procedure | Procedure grouping 20 |
| Minor and Excluded Procedures | Excluded |

**NCHDA National Congenital Heart Disease Audit*

### Supplementary Table S4: PRAIS 2 risk factor groups used for adjusting the case mix

| **RECOGNISED OFFICIAL NCHDA CO-MORBIDITIES*** | **Additional Risk Factor Group** |
| --- | --- |
| 101351. Pulmonary embolism | Acquired Comorbidity |
| 101400. Secondary systemic hypertension | Acquired Comorbidity |
| 101401. Systemic hypertension | Acquired Comorbidity |
| 101402. Primary (essential) systemic hypertension | Acquired Comorbidity |
| 101404. Systemic hypertension due to aortic arch obstruction | Acquired Comorbidity |
| 101501. Persistent pulmonary hypertension of the newborn (PFC), | Acquired Comorbidity |
| 101505. Necrotising enterocolitis | Acquired Comorbidity |
| 101512. Meconium aspiration | Acquired Comorbidity |
| 102006. Pre-procedural coagulation disorder | Acquired Comorbidity |
| 102007. Pre-procedural renal failure | Acquired Comorbidity |
| 102008. Pre-procedural renal failure requiring dialysis | Acquired Comorbidity |
| 102009. Pre-procedural septicaemia | Acquired Comorbidity |
| 102012. Pre-procedural neurological impairment | Acquired Comorbidity |
| 102013. Preprocedural cerebral abnormality on imaging | Acquired Comorbidity |
| 102017. Pre-procedural tracheostomy | Acquired Comorbidity |
| 102018. Preprocedural seizures | Acquired Comorbidity |
| 102037. Preprocedural respiratory syncytial virus (RSV) infection | Acquired Comorbidity |
| 102038. Preprocedural necrotising enterocolitis: treated medically | Acquired Comorbidity |
| 102039. Preprocedural necrotising enterocolitis: treated surgically | Acquired Comorbidity |
| 140305. Psychomotor developmental delay | Acquired Comorbidity |
| 140340. Brain Abcess | Acquired Comorbidity |
| 140342. Cerebrovascular accident (stroke) | Acquired Comorbidity |
| 140372. Anoxic-ischaemic encephalopathy | Acquired Comorbidity |
| 140375. Hyperthyroidism | Acquired Comorbidity |
| 140390. Diabetes mellitus | Acquired Comorbidity |
| 140494. Diabetes mellitus: requiring insulin | Acquired Comorbidity |
| 140565. Meningitis | Acquired Comorbidity |
| 158210. Kidney failure | Acquired Comorbidity |
| 160111. Empyema | Acquired Comorbidity |
| 160302. Lower respiratory tract infection | Acquired Comorbidity |
| 160305. Lung disease | Acquired Comorbidity |
| 160310. Asthma | Acquired Comorbidity |
| 160800. Acquired bronchial disease | Acquired Comorbidity |
| 160900. Airway disease, | Acquired Comorbidity |
| 161300. Diaphragm disorder: acquired | Acquired Comorbidity |
| 161320. Diaphragm paralysis, | Acquired Comorbidity |
| 162010. Oesophageal disorder | Acquired Comorbidity |
| 070001. Ventricular dyssynchrony | Additional Cardiac Risk Factors |
| 070110. Arrhythmogenic right ventricular cardiomyopathy | Additional Cardiac Risk Factors |
| 070111. Right ventricular dysfunction | Additional Cardiac Risk Factors |
| 070610. Left ventricular dysfunction | Additional Cardiac Risk Factors |
| 070850. Ventricular myocardial noncompaction cardiomyopathy | Additional Cardiac Risk Factors |
| 100701. Infectious myocarditis | Additional Cardiac Risk Factors |
| 100703. Viral myocarditis | Additional Cardiac Risk Factors |
| 100705. Drug induced heart muscle disease | Additional Cardiac Risk Factors |
| 100708. Trypanosomal myocarditis (Chagas' disease) | Additional Cardiac Risk Factors |
| 100740. Myocardial failure in end stage congenital heart disease | Additional Cardiac Risk Factors |
| 100742. Heart muscle disease in cardiac rejection | Additional Cardiac Risk Factors |
| 100761. Nutritional heart muscle disease | Additional Cardiac Risk Factors |
| 100771. Heart muscle disease in infant of diabetic mother | Additional Cardiac Risk Factors |
| 100781. Heart muscle disease in collagen vascular/ connective tissue disorder | Additional Cardiac Risk Factors |
| 100790. Myocarditis | Additional Cardiac Risk Factors |
| 100930. Ischaemic heart disease | Additional Cardiac Risk Factors |
| 101001. Cardiomyopathy | Additional Cardiac Risk Factors |
| 101010. Restrictive cardiomyopathy | Additional Cardiac Risk Factors |
| 101011. Idiopathic restrictive cardiomyopathy | Additional Cardiac Risk Factors |
| 101012. Endocardial fibroelastosis | Additional Cardiac Risk Factors |
| 101013. Infiltrative cardiomyopathy | Additional Cardiac Risk Factors |
| 101020. Hypertrophic cardiomyopathy | Additional Cardiac Risk Factors |
| 101025. Dilated cardiomyopathy | Additional Cardiac Risk Factors |
| 101301. Pulmonary arterial hypertension | Additional Cardiac Risk Factors |
| 101302. Idiopathic (primary) pulmonary hypertension | Additional Cardiac Risk Factors |
| 101306. Pulmonary vascular disease | Additional Cardiac Risk Factors |
| 101308. Irreversible pulmonary vascular disease due to congenital heart disease (Eisenmenger Syndrome) | Additional Cardiac Risk Factors |
| 101320. Secondary pulmonary hypertension | Additional Cardiac Risk Factors |
| 101321. Pulmonary hypertension due to congenital systemic-to-pulmonary shunt | Additional Cardiac Risk Factors |
| 101363. Elevated lung resistance for biventricular repair (> 6 Wood units) | Additional Cardiac Risk Factors |
| 101364. Elevated lung resistance for heart transplant (> 4 Wood units) | Additional Cardiac Risk Factors |
| 101365. Elevated lung resistance for univentricular repair (> 2 Wood units) | Additional Cardiac Risk Factors |
| 101510. Transient myocardial ischaemia | Additional Cardiac Risk Factors |
| 101800. Myocardial infarction | Additional Cardiac Risk Factors |
| 101801. Acute myocardial infarction | Additional Cardiac Risk Factors |
| 102016. Pre-procedural pulmonary hypertension | Additional Cardiac Risk Factors |
| 102034. Preprocedural myocardial infarction | Additional Cardiac Risk Factors |
| 102045. Preprocedural pulmonary hypertension (pulmonary pressure more than or equal to systemic pressure): echo data, | Additional Cardiac Risk Factors |
| 102046. Preprocedural pulmonary hypertension (pulmonary pressure more than or equal to systemic pressure): catheter data, | Additional Cardiac Risk Factors |
| 152231. Residual pulmonary hypertension after relief of L to R shunt | Additional Cardiac Risk Factors |
| 030102. Visceral heterotaxy (abnormal arrangement thoraco-abdominal organs), | Congenital Comorbidity |
| 030109. Position or morphology of thoraco-abdominal organs abnormal | Congenital Comorbidity |
| 030209. Lung anomaly | Congenital Comorbidity |
| 030214. Functionally congenital single lung | Congenital Comorbidity |
| 030305. Tracheobronchial anomaly | Congenital Comorbidity |
| 030603. Intestines malrotated, | Congenital Comorbidity |
| 102304. Hereditary disorder associated with heart disease | Congenital Comorbidity |
| 140101. Chromosomal anomaly | Congenital Comorbidity |
| 140103. Trisomy 18 - Edwards syndrome | Congenital Comorbidity |
| 140104. Trisomy 13 - Pataus syndrome | Congenital Comorbidity |
| 140105. 45XO - Turners syndrome | Congenital Comorbidity |
| 140121. 22q11 microdeletion - CATCH 22 | Congenital Comorbidity |
| 140200. Syndrome/association with cardiac involvement | Congenital Comorbidity |
| 140206. DiGeorge sequence | Congenital Comorbidity |
| 140210. Friedreich’s ataxia, | Congenital Comorbidity |
| 140217. Marfan syndrome | Congenital Comorbidity |
| 140219. Noonan syndrome | Congenital Comorbidity |
| 140221. Pompe’s disease: glycogen storage disease type IIa, | Congenital Comorbidity |
| 140228. Tuberous sclerosis | Congenital Comorbidity |
| 140230. Williams syndrome (infantile hypercalcaemia) | Congenital Comorbidity |
| 140232. Fetal rubella syndrome | Congenital Comorbidity |
| 140234. Duchenne’s muscular dystrophy, | Congenital Comorbidity |
| 140258. Muscular dystrophy, | Congenital Comorbidity |
| 140262. Ehlers-Danlos syndrome | Congenital Comorbidity |
| 140266. Alagille syndrome: arteriohepatic dysplasia | Congenital Comorbidity |
| 140300. Non-cardiac abnormality associated with heart disease | Congenital Comorbidity |
| 140304. Non-cardiothoracic / vascular abnormality (DESCRIBE) | Congenital Comorbidity |
| 140306. Cystic fibrosis | Congenital Comorbidity |
| 140307. Diaphragmatic hernia | Congenital Comorbidity |
| 140308. Tracheo-oesophageal fistula | Congenital Comorbidity |
| 140310. Omphalocoele | Congenital Comorbidity |
| 140311. Duodenal stenosis/atresia | Congenital Comorbidity |
| 140321. Sickle cell disease | Congenital Comorbidity |
| 140323. Renal abnormality | Congenital Comorbidity |
| 140328. Congenital coagulation disorder, | Congenital Comorbidity |
| 140329. Thoracic / mediastinal abnormality | Congenital Comorbidity |
| 140333. Microcephaly | Congenital Comorbidity |
| 140347. Choanal atresia, | Congenital Comorbidity |
| 140349. Tracheobronchial malacia | Congenital Comorbidity |
| 140352. Hypothyroidism | Congenital Comorbidity |
| 140391. Cerebral anomaly | Congenital Comorbidity |
| 140392. Connective tissue disease, | Congenital Comorbidity |
| 140409. Kyphoscoliosis | Congenital Comorbidity |
| 140412. Cleft lip / palate | Congenital Comorbidity |
| 140485. Loeys-Dietz Syndrome (transforming growth factor beta receptor (TGFBR) gene) | Congenital Comorbidity |
| 140490. Von Willebrand disease | Congenital Comorbidity |
| 140540.  Maternally derived fetal disease or syndrome associated with heart disease, | Congenital Comorbidity |
| 140550. Major anomaly of gastrointestinal system | Congenital Comorbidity |
| 140601. Multiple congenital malformations | Congenital Comorbidity |
| 161001. Tracheal stenosis | Congenital Comorbidity |
| 161009. Tracheal disease | Congenital Comorbidity |
| 140102. Trisomy 21 - Downs syndrome | Downs Syndrome (not used in PRAiS) |
| 101600. Right ventricular abnormality: acquired | None (not used in PRAiS) |
| 101608. Right ventricular-congestive heart failure | None (not used in PRAiS) |
| 101640. Left ventricular abnormality: acquired | None (not used in PRAiS) |
| 101647. Left ventricular failure | None (not used in PRAiS) |
| 102202. Premature birth | Premature (not used in PRAiS) |
| 102205. Premature birth 32-35 weeks, | Premature (not used in PRAiS) |
| 102206. Premature birth less than 32 weeks, | Premature (not used in PRAiS) |
| 101723. Shock | Severity of illness |
| 102002. Pre-procedural shock | Severity of illness |
| 102005. Pre-procedural acidosis | Severity of illness |
| 102014. Pre-procedural mechanical ventilatory support | Severity of illness |
| 102015. Pre-procedural mechanical circulatory support | Severity of illness |
| 102031. Preprocedural shock at time of surgery (persistent) | Severity of illness |
| 102033. Preprocedural cardiopulmonary resuscitation (< 48 hours) | Severity of illness |
| 110021. Cardiac Arrest | Severity of illness |
| 163001. Respiratory failure | Severity of illness |

**codes taken form the European Paediatric Cardiac Code Short List OPCSV4.7 -1 April 2015*

### Supplementary Table S5: Description of covariates used in the analysis.

| **Variable** | **Description** | **Source** | **Codes used** |
| --- | --- | --- | --- |
| Age at procedure | Continuous variable | Primary or secondary care data |  |
| Gender | Categorical variable  Male  Female | National Congenital Heart Disease Audit (NCHDA) or primary or secondary care data |  |
| Deprivation | Categorical variable (quintiles) 1- most deprived; 5 least deprived | Primary or secondary care | Based on LSOA reference table and english_indices_of_dep_v02 (corporate asset) |
| Region | Categorical variable | Primary or secondary care | Based on LSOA reference table and curated asset from consortium curr901a_lsoa_region_lookup |
| Ethnicity | White  South Asian  African / Caribbean  Mixed  Others  Not stated/unknown | NCHDA or primary or secondary care |  |
| Weight | Continuous variables (kg) | NCHDA |  |
| Primary diagnosis | Categorical variable  (27 diagnostic groupings) | NCHDA | EPCC adapted from IPCC diagnostic codes downloaded from <http://nicor4.nicor.org.uk> |
| Urgency of procedure | Categorical variable  Elective  Urgent  Emergency  Life-saving | NCHDA |  |
| Procedure activity group | Categorical variable  Cardiac surgery  Interventional catheter  Diagnostic catheter  Electrophysiology  Mechanical support  Chest closure and exploration | NCHDA | 01.activity_analysis_algorithm_v6.14. R script and EPCC adapted from IPCC diagnostic codes downloaded from [http://nicor4.nicor.org.uk](http://www.ucl.ac.uk/nicor) |
| Specific procedure group | Categorical variable  (88 recognizable operations) | NCHDA | 01.specific_procedure_algorithm_v6.05.R script EPCC adapted from IPCC diagnostic codes downloaded from [http://nicor4.nicor.org.uk](http://www.ucl.ac.uk/nicor) |
| Complication | Categorical variable | NCHDA | EPCC adapted from IPCC diagnostic codes downloaded from [http://nicor4.nicor.org.uk](http://www.ucl.ac.uk/nicor) |
| Unplanned reoperation | Categorical variable | NCHDA |  |
| Post procedure seizure | Categorical variable | NCHDA |  |
| Catheterization complication severity rating | Categorical variable  Mild  Moderate  Major  Catastrophic | NCHDA |  |
| Discharge destination | Categorical variable | NCHDA |  |
| Discharge status | Categorical variable | NCHDA |  |
| Hospital stay | Continuous variable | NCHDA |  |
| PRAiS2 risk score | Continuous variable | Derived from NCHDA variables |  |
| Additional cardiac risk factor | Categorical variable | NCHDA | EPCC adapted from IPCC diagnostic codes downloaded from [http://nicor4.nicor.org.uk](http://www.ucl.ac.uk/nicor) |
| Acquired comorbidity | Categorical variable | NCHDA | EPCC adapted from IPCC diagnostic codes downloaded from [http://nicor4.nicor.org.uk](http://www.ucl.ac.uk/nicor) |
| Congenital comorbidity | Categorical variable | NCHDA | EPCC adapted from IPCC diagnostic codes downloaded from [http://nicor4.nicor.org.uk](http://www.ucl.ac.uk/nicor) |
| Severity of illness indicator | Categorical variable | NCHDA | EPCC adapted from IPCC diagnostic codes downloaded from [http://nicor4.nicor.org.uk](http://www.ucl.ac.uk/nicor) |

*NCHDA – National Congenital Heart Disease Audit; EPCC- European Paediatric Congenital Cardiac Codes; IPCC- International Paediatric Congenital Cardiac Codes; LSOA – Lower Layer Super Output Areas*

### Supplementary Table S6: Distribution of the primary diagnosis among children who had congenital heart disease in England between 01 Jan 2018 and 31 Mar 2022

| **Primary diagnosis** | **n (%)** |
| --- | --- |
| Patent Ductus Arteriosus | 2255 (8.6%) |
| Arrhythmia | 1845 (7.0%) |
| Ventricular Septal Defect | 1845 (7.0%) |
| MISC congenital primary diagnosis^2^ | 1680 (6.4%) |
| Hypoplastic left heart syndrome | 1640 (6.2%) |
| Fallot / DORV-Fallot type | 1620 (6.2%) |
| Interatrial communication ('Atrial septal defect') | 1555 (5.9%) |
| Functionally univentricular heart | 1500 (5.7%) |
| Aortic arch obstruction +/- VSD/ASD^1^ | 1460 (5.6%) |
| Pulmonary atresia + VSD (including Fallot type)^5^ | 1425 (5.4%) |
| Atrioventricular septal defect | 1400 (5.3%) |
| TGA+VSD/ DORV-TGA type ^6^ | 1175 (4.5%) |
| Acquired | 1115 (4.2%) |
| Pulmonary stenosis ^5^ | 995 (3.8%) |
| Aortic valve stenosis (isolated) ^1^ | 840 (3.2%) |
| Mitral valve abnormality (including supravalvar, subvalvar) ^3^ | 640 (2.4%) |
| Tricuspid valve abnormality (including Ebstein’s) ^3^ | 600 (2.3%) |
| Transposition of great arteries (concordant AV & discordant VA connections) & IVS ^6^ | 485 (1.8%) |
| Miscellaneous congenital terms | 410 (1.6%) |
| Common arterial trunk (truncus arteriosus) | 400 (1.5%) |
| Pulmonary atresia and IVS ^5^ | 290 (1.1%) |
| Total Anomalous Pulmonary Venous Connection (pulmonary related CHD)^1^ | 235 (0.9%) |
| Subaortic stenosis (isolated) ^1^ | 185 (0.7%) |
| Aortic regurgitation ^2^ | 150 (0.6%) |
| Interrupted aortic arch ^1^ | 140 (0.5%) |
| Empty/ unknown^4^ | 95 (0.4%) |
| Comorbidity^4^ | 75 (0.3%) |
| Normal^4^ | 45 (0.2%) |
| Missing^4^ | 175 (0.7%) |

*1- Left ventricular outflow obstruction; 2 – Miscellaneous congenital primary diagnosis; 3- Primary atrioventricular valvar disease; 4- Primary congenital diagnosis information missing; 5- Pulmonary atresia and stenosis; 6-Transposition of great arteries*

### Supplementary Table S7: Difference in mean percentage of all specific procedures during different phases of restriction compared to the pre pandemic period among all congenital heart disease procedures among children in England

| **Procedure** | **Pre-pandemic (01 Jan 2018 to**  **22 Mar 2020) percentage** | **Percentage difference from pre-pandemic period (95%CI)** | | | | | |
| --- | --- | --- | --- | --- | --- | --- | --- |
|  |  | **First restriction**  **(23 Mar 2020 to**  **23 Jun 2020)** | **First relaxation**  **22 Jun 2020 to**  **04-Nov-20** | **Second restriction**  **05 Nov 2020 to**  **02-Dec-20** | **Second relaxation**  **03 Dec 2020 to**  **05-Jan-21** | **Third restriction 06 Jan 2021 to**  **21-Jun-21** | **Post third restriction**  **22 Jun 2021 to**  **31-Mar-22** |
| Unallocated | 13.33 | 0.79 (-1.33,2.9) | -1.66 (-3.07,-0.24) | 2.34 (-0.95,5.62) | 1.16 (-2.25,4.56) | -0.65 (-2.02,0.73) | 0.09 (-1.06,1.24) |
| Catheter diagnostic | 11.67 | -1.3 (-3.16,0.56) | 0.95 (-0.49,2.4) | -0.12 (-3.01,2.77) | -3.36 (-6.04,-0.67) | -0.21 (-1.53,1.1) | 0.39 (-0.7,1.49) |
| PDA transluminal | 6.47 | -1.37 (-2.72,-0.02) | 0.06 (-1.02,1.14) | -1.52 (-3.49,0.45) | 0.42 (-2.03,2.87) | 0.85 (-0.21,1.91) | 1.73 (0.82,2.64) |
| EP ablation | 5.33 | -3.01 (-3.96,-2.05) | 0.38 (-0.63,1.39) | 2.09 (-0.27,4.45) | 2.51 (-0.09,5.1) | 0.15 (-0.78,1.09) | 0.15 (-0.62,0.91) |
| Ventricular Septal Defect | 3.98 | 0.31 (-0.92,1.54) | -0.12 (-0.96,0.73) | 0.97 (-0.99,2.92) | -1.37 (-2.92,0.19) | -0.44 (-1.2,0.33) | -0.05 (-0.7,0.61) |
| Fallot | 2.97 | 2.48 (1.13,3.84) | -0.05 (-0.78,0.69) | -0.49 (-1.9,0.92) | -0.35 (-1.9,1.19) | -0.28 (-0.95,0.39) | -0.41 (-0.96,0.13) |
| Coarctation hypoplasia | 2.82 | 1.2 (0.02,2.38) | -0.42 (-1.09,0.26) | 0.27 (-1.29,1.84) | 1.93 (-0.12,3.98) | -0.13 (-0.8,0.53) | -0.06 (-0.61,0.5) |
| Balloon pulmonary valve | 2.78 | 0.26 (-0.78,1.3) | 0.18 (-0.55,0.92) | -1.34 (-2.43,-0.24) | -0.88 (-2.21,0.45) | 0.91 (0.15,1.68) | -0.06 (-0.61,0.49) |
| ASD transluminal | 2.67 | -1.6 (-2.26,-0.94) | -0.87 (-1.47,-0.27) | 0.01 (-1.45,1.47) | -1.25 (-2.41,-0.09) | -0.32 (-0.95,0.31) | 0 (-0.55,0.54) |
| PA ballooning | 2.61 | 0.08 (-0.9,1.06) | 0.06 (-0.65,0.76) | 0.28 (-1.23,1.79) | 0.01 (-1.54,1.55) | 0.04 (-0.62,0.7) | 0.62 (0.03,1.2) |
| PDA ligation | 2.6 | -0.36 (-1.27,0.54) | -0.11 (-0.79,0.57) | -0.33 (-1.68,1.02) | -1.41 (-2.48,-0.34) | -1.14 (-1.66,-0.62) | -1.1 (-1.54,-0.66) |
| Atrial Septal Defect | 2.12 | -1.76 (-2.18,-1.34) | 0.54 (-0.15,1.23) | -0.88 (-1.9,0.13) | -0.22 (-1.55,1.1) | -0.44 (-0.98,0.1) | -0.86 (-1.26,-0.45) |
| Glenn | 1.92 | 1.12 (0.09,2.15) | 0.44 (-0.21,1.1) | -0.06 (-1.29,1.16) | -0.5 (-1.65,0.66) | 0.58 (-0.05,1.21) | 0.06 (-0.41,0.53) |
| Fontan | 1.81 | -1.54 (-1.92,-1.17) | 0.16 (-0.44,0.77) | -0.58 (-1.58,0.43) | -0.39 (-1.54,0.76) | -0.13 (-0.67,0.4) | 0.21 (-0.26,0.68) |
| Atrio ventricular septal defect complete | 1.79 | 0.44 (-0.45,1.33) | 0.35 (-0.27,0.98) | -0.97 (-1.8,-0.14) | -0.61 (-1.66,0.45) | 0.37 (-0.22,0.96) | 0.39 (-0.09,0.88) |
| Sub valvar aortic stenosis | 1.66 | 0.4 (-0.46,1.25) | 0.4 (-0.21,1.01) | 0.2 (-1.02,1.42) | -0.71 (-1.66,0.24) | -0.05 (-0.57,0.46) | -0.05 (-0.47,0.38) |
| Vascular ring | 1.61 | 0.27 (-0.56,1.09) | 0.15 (-0.42,0.72) | 0.45 (-0.83,1.73) | -0.19 (-1.34,0.96) | -0.01 (-0.52,0.51) | 0.46 (-0.01,0.93) |
| Balloon atrial septostomy | 1.55 | 1.31 (0.31,2.31) | -0.01 (-0.54,0.53) | -0.73 (-1.56,0.1) | 1.06 (-0.47,2.6) | -0.21 (-0.69,0.27) | -0.35 (-0.73,0.03) |
| Transposition | 1.48 | 0.66 (-0.21,1.53) | 0.23 (-0.33,0.8) | -0.25 (-1.25,0.76) | 0.42 (-0.9,1.73) | 0.34 (-0.2,0.89) | -0.31 (-0.68,0.06) |
| PA stent | 1.38 | -0.75 (-1.25,-0.25) | 0 (-0.51,0.51) | -0.76 (-1.48,-0.04) | 0.29 (-0.95,1.52) | -0.29 (-0.73,0.14) | -0.13 (-0.51,0.25) |
| PA band | 1.32 | 0.11 (-0.61,0.83) | -0.37 (-0.8,0.06) | -0.08 (-1.08,0.92) | 0.82 (-0.57,2.22) | -0.16 (-0.6,0.29) | -0.1 (-0.47,0.28) |
| EP diagnostic | 1.2 | -0.58 (-1.07,-0.08) | -0.13 (-0.58,0.33) | 0.24 (-0.83,1.32) | -0.01 (-1.06,1.04) | -0.08 (-0.52,0.35) | 0.41 (0,0.82) |
| Pacemaker endocardial | 1.16 | 0.8 (-0.03,1.64) | -0.09 (-0.54,0.36) | -0.13 (-1.05,0.78) | 0.26 (-0.88,1.41) | 0.03 (-0.41,0.48) | 0.22 (-0.17,0.61) |
| MAPCA transluminal | 1.13 | 0.03 (-0.62,0.68) | 0.33 (-0.18,0.85) | -0.1 (-1.01,0.82) | -0.18 (-1.12,0.76) | 0.51 (0,1.02) | 0.07 (-0.3,0.43) |
| Norwood | 1.08 | 0.62 (-0.16,1.39) | 0.04 (-0.42,0.49) | -0.05 (-0.96,0.86) | 1.29 (-0.17,2.76) | 0.08 (-0.36,0.51) | -0.09 (-0.43,0.25) |
| Balloon aortic valve | 1.07 | -0.72 (-1.1,-0.33) | 0 (-0.45,0.45) | 0.58 (-0.57,1.72) | 0.35 (-0.79,1.5) | -0.33 (-0.69,0.04) | -0.06 (-0.4,0.28) |
| TAPVC | 0.83 | -0.12 (-0.63,0.4) | -0.45 (-0.74,-0.15) | -0.42 (-1.01,0.17) | 0.12 (-0.82,1.06) | -0.05 (-0.41,0.32) | -0.03 (-0.33,0.28) |
| PR PA conduit | 0.83 | -0.11 (-0.63,0.4) | -0.35 (-0.67,-0.04) | -0.41 (-1,0.18) | -- | -0.45 (-0.73,-0.18) | -0.32 (-0.58,-0.06) |
| Pacemaker epicardial | 0.83 | 0.25 (-0.37,0.87) | -0.01 (-0.4,0.38) | -0.21 (-0.92,0.51) | -0.59 (-1.08,-0.1) | 0.22 (-0.19,0.63) | -0.11 (-0.4,0.18) |
| AVSD partial | 0.81 | -0.1 (-0.61,0.42) | -0.13 (-0.49,0.24) | 0.01 (-0.81,0.83) | 0.14 (-0.8,1.08) | -0.03 (-0.39,0.33) | -0.1 (-0.39,0.19) |
| Aortic valve repair | 0.79 | 0.01 (-0.53,0.55) | 0.32 (-0.13,0.77) | 0.03 (-0.78,0.85) | -0.32 (-0.99,0.35) | -0.05 (-0.4,0.31) | -0.19 (-0.46,0.08) |
| Recoarctation balloon | 0.78 | -0.06 (-0.58,0.45) | -0.26 (-0.59,0.06) | -0.37 (-0.95,0.22) | 0.17 (-0.77,1.11) | 0.08 (-0.3,0.46) | 0.03 (-0.27,0.33) |
| PDA stent | 0.76 | 0.49 (-0.17,1.16) | -0.03 (-0.4,0.34) | -0.14 (-0.85,0.57) | 0.43 (-0.62,1.47) | -0.05 (-0.4,0.3) | 0.14 (-0.17,0.45) |
| RVOT stent | 0.76 | 0.31 (-0.31,0.93) | -0.11 (-0.47,0.24) | -0.55 (-0.98,-0.13) | -0.28 (-0.96,0.39) | -0.12 (-0.46,0.21) | -0.32 (-0.56,-0.08) |
| Pulmonary valve replacement | 0.74 | -0.11 (-0.59,0.37) | -0.18 (-0.51,0.15) | 0.09 (-0.73,0.9) | -0.5 (-0.99,-0.02) | -0.22 (-0.52,0.09) | -0.14 (-0.41,0.13) |
| Conduit balloon stent | 0.71 | -0.44 (-0.78,-0.11) | 0.53 (0.06,1) | 0.73 (-0.34,1.8) | 0.24 (-0.7,1.17) | 0.26 (-0.14,0.65) | 0.39 (0.05,0.73) |
| Sinus venosus ASD PAPVC | 0.67 | -0.14 (-0.58,0.31) | 0.19 (-0.21,0.58) | 0.36 (-0.55,1.27) | -0.43 (-0.92,0.05) | 0 (-0.34,0.34) | -0.26 (-0.49,-0.03) |
| VSD RVOTO | 0.61 | 0.01 (-0.46,0.49) | -0.05 (-0.38,0.27) | 0.01 (-0.7,0.72) | -0.14 (-0.8,0.53) | 0.14 (-0.21,0.48) | -0.22 (-0.44,0) |
| Mitral valve replacement | 0.59 | -0.14 (-0.55,0.27) | -0.2 (-0.48,0.08) | 0.44 (-0.47,1.35) | 0.6 (-0.45,1.64) | -0.11 (-0.4,0.18) | -0.04 (-0.29,0.21) |
| Transposition VSD | 0.56 | 0.52 (-0.1,1.13) | 0.04 (-0.29,0.38) | -0.35 (-0.77,0.07) | 0.63 (-0.41,1.67) | -0.11 (-0.39,0.17) | -0.05 (-0.29,0.19) |
| Arterial shunt | 0.55 | 0.52 (-0.09,1.14) | -0.21 (-0.47,0.06) | -0.34 (-0.77,0.08) | -0.31 (-0.79,0.17) | 0.05 (-0.27,0.36) | -0.02 (-0.27,0.23) |
| Mitral valve repair | 0.52 | -0.17 (-0.53,0.2) | -0.22 (-0.47,0.03) | 0.51 (-0.4,1.41) | 0.19 (-0.62,1) | 0.19 (-0.15,0.52) | 0.05 (-0.2,0.3) |
| Tricuspid valve repair | 0.52 | 0.11 (-0.37,0.58) | -0.04 (-0.35,0.26) | 0.1 (-0.61,0.81) | 0.43 (-0.5,1.37) | 0.23 (-0.12,0.57) | -0.19 (-0.4,0.01) |
| Conduit replacement | 0.5 | 0.13 (-0.35,0.6) | 0.32 (-0.06,0.7) | -0.29 (-0.71,0.13) | -0.26 (-0.74,0.22) | -0.01 (-0.3,0.27) | -0.08 (-0.3,0.14) |
| ICD catheter | 0.5 | 0.04 (-0.4,0.48) | 0.23 (-0.13,0.6) | 0.74 (-0.25,1.73) | 0.45 (-0.48,1.39) | 0.17 (-0.15,0.5) | 0.22 (-0.06,0.49) |
| Pacemaker lead | 0.42 | 0.02 (-0.38,0.43) | -0.17 (-0.4,0.07) | 0.61 (-0.3,1.51) | 0.53 (-0.41,1.46) | 0.32 (-0.02,0.67) | 0.08 (-0.15,0.32) |
| VSD transluminal | 0.41 | -0.32 (-0.52,-0.12) | -0.19 (-0.41,0.02) | -- | 0.3 (-0.51,1.11) | 0.04 (-0.23,0.31) | 0 (-0.21,0.22) |
| Ross | 0.38 | 0.24 (-0.23,0.72) | 0.09 (-0.21,0.39) | 0.03 (-0.55,0.61) | 0.57 (-0.36,1.5) | 0.07 (-0.21,0.34) | 0.03 (-0.18,0.25) |
| Coarctation stent | 0.38 | 0.25 (-0.22,0.72) | 0.14 (-0.17,0.45) | -0.17 (-0.59,0.25) | 0.1 (-0.57,0.76) | 0.07 (-0.2,0.34) | -0.03 (-0.23,0.17) |
| Stent dilatation | 0.38 | -0.11 (-0.43,0.2) | 0.3 (-0.05,0.65) | 0.85 (-0.13,1.84) | -- | -0.12 (-0.34,0.1) | 0.1 (-0.13,0.33) |
| Pulmonary atresia VSD | 0.36 | -0.01 (-0.37,0.36) | -0.1 (-0.33,0.12) | 0.05 (-0.53,0.63) | 0.11 (-0.55,0.78) | 0.31 (-0.01,0.63) | -0.13 (-0.3,0.04) |
| Pulmonary vein intervention | 0.36 | 0 (-0.36,0.36) | 0.12 (-0.18,0.41) | 0.26 (-0.44,0.97) | -0.12 (-0.59,0.36) | 0.39 (0.05,0.73) | 0.36 (0.09,0.63) |
| Heart transplant | 0.33 | 0.12 (-0.28,0.52) | 0.23 (-0.09,0.55) | 0.29 (-0.41,0.99) | -0.09 (-0.57,0.38) | -0.03 (-0.26,0.2) | 0.09 (-0.13,0.3) |
| Common arterial trunk repair | 0.32 | -0.14 (-0.41,0.12) | -0.02 (-0.26,0.22) | 0.09 (-0.49,0.67) | 0.15 (-0.51,0.82) | -0.06 (-0.27,0.15) | -0.07 (-0.24,0.11) |
| Supra valvar aortic stenosis | 0.28 | -0.1 (-0.36,0.16) | 0.02 (-0.22,0.26) | -- | -0.04 (-0.52,0.43) | 0.05 (-0.18,0.29) | -0.05 (-0.22,0.11) |
| Pulmonary valve replacement transluminal | 0.28 | -0.1 (-0.36,0.16) | 0.02 (-0.22,0.26) | -0.08 (-0.49,0.34) | -- | -0.13 (-0.3,0.04) | 0.02 (-0.17,0.2) |
| EP miscellaneous | 0.27 | 0.36 (-0.11,0.83) | -0.05 (-0.26,0.15) | 0.14 (-0.43,0.72) | -0.03 (-0.5,0.44) | 0.07 (-0.17,0.3) | 0.08 (-0.12,0.27) |
| Coarctation balloon | 0.26 | -0.08 (-0.34,0.18) | -0.04 (-0.25,0.16) | -0.05 (-0.46,0.36) | -- | -0.03 (-0.23,0.17) | -0.02 (-0.19,0.14) |
| IAA | 0.24 | 0.21 (-0.19,0.6) | 0.06 (-0.18,0.29) | -- | 0 (-0.48,0.47) | -0.17 (-0.3,-0.04) | -0.1 (-0.24,0.03) |
| Pulmonary vein stenosis | 0.23 | -0.14 (-0.33,0.05) | -0.06 (-0.24,0.13) | -- | 0.01 (-0.46,0.48) | -0.04 (-0.22,0.14) | -0.09 (-0.22,0.04) |
| Ross konno a | 0.22 | 0.14 (-0.22,0.49) | 0.12 (-0.13,0.37) | -- | 0.02 (-0.46,0.49) | -0.07 (-0.24,0.09) | 0.03 (-0.14,0.2) |
| Anomalous coronary | 0.22 | 0.23 (-0.17,0.62) | 0.04 (-0.18,0.26) | -0.02 (-0.43,0.4) | 0.49 (-0.32,1.3) | 0 (-0.19,0.2) | -0.01 (-0.17,0.14) |
| Transposition arch | 0.18 | 0 (-0.26,0.25) | -0.05 (-0.21,0.11) | 0.23 (-0.34,0.81) | -- | -0.07 (-0.21,0.07) | 0 (-0.14,0.15) |
| Aortic root replacement | 0.18 | 0.09 (-0.22,0.4) | -0.14 (-0.25,-0.03) | 0.02 (-0.38,0.43) | -- | -0.03 (-0.19,0.13) | -0.16 (-0.24,-0.08) |
| MAPCA unifocalisation | 0.18 | -0.09 (-0.28,0.1) | -0.1 (-0.23,0.04) | -- | -- | -0.11 (-0.23,0.02) | -0.02 (-0.16,0.12) |
| Transposition complex | 0.17 | 0.1 (-0.21,0.41) | 0 (-0.18,0.18) | -- | -- | 0.06 (-0.13,0.25) | -0.03 (-0.16,0.1) |
| Rastelli rev | 0.15 | -0.06 (-0.24,0.13) | 0.28 (0.01,0.55) | 0.06 (-0.35,0.47) | 0.09 (-0.38,0.56) | 0.15 (-0.06,0.37) | 0.08 (-0.07,0.24) |
| Aortic valve replacement | 0.15 | 0.03 (-0.22,0.29) | -0.02 (-0.18,0.14) | 0.06 (-0.35,0.47) | 0.09 (-0.38,0.56) | 0.26 (0.01,0.51) | 0.06 (-0.09,0.21) |
| AP window | 0.15 | 0.12 (-0.19,0.43) | 0.02 (-0.16,0.2) | 0.06 (-0.35,0.47) | 0.33 (-0.33,0.99) | -0.07 (-0.19,0.05) | -0.06 (-0.16,0.05) |
| Multiple VSD | 0.15 | -0.06 (-0.24,0.13) | -0.06 (-0.2,0.07) | 0.06 (-0.35,0.47) | -- | -- | 0.06 (-0.09,0.21) |
| Cor triatriatum | 0.13 | -- | -0.08 (-0.19,0.02) | 0.08 (-0.33,0.49) | 0.11 (-0.36,0.58) | -0.02 (-0.15,0.12) | -0.06 (-0.16,0.04) |
| Pulmonary valve radiofrequency | 0.13 | 0.14 (-0.17,0.45) | 0 (-0.16,0.16) | 0.08 (-0.33,0.49) | 0.35 (-0.31,1.01) | 0.21 (-0.02,0.43) | 0.08 (-0.07,0.23) |
| CCTGArepair a | 0.11 | -- | 0.02 (-0.13,0.18) | -- | -- | -0.07 (-0.16,0.02) | 0.01 (-0.11,0.12) |
| Absent pulmonary valve syndrome | 0.08 | 0.01 (-0.17,0.19) | -- | -- | 0.39 (-0.26,1.05) | -0.04 (-0.13,0.04) | -0.06 (-0.12,0.01) |
| Lung transplant | 0.07 | 0.02 (-0.16,0.2) | -- | -- | -- | 0 (-0.11,0.11) | -0.05 (-0.11,0.01) |
| AVSD Fallot a | 0.07 | 0.38 (-0.01,0.77) | 0.15 (-0.04,0.34) | -- | -- | 0.01 (-0.1,0.12) | -0.02 (-0.1,0.06) |
| Tricuspid valve replacement | 0.07 | 0.02 (-0.16,0.2) | 0.06 (-0.09,0.21) | -- | -- | 0.08 (-0.07,0.23) | -0.04 (-0.11,0.02) |
| Pacemaker crt bv | 0.07 | 0.11 (-0.14,0.36) | -- | 0.14 (-0.27,0.54) | -- | -0.03 (-0.11,0.05) | 0.05 (-0.06,0.16) |
| Common arterial trunk aorta repair | 0.05 | 0.13 (-0.12,0.38) | -- | -- | -- | -- | -0.02 (-0.08,0.03) |
| Ross konno b | 0.05 | -- | -- | 0.16 (-0.25,0.56) | -- | 0.06 (-0.07,0.2) | 0.02 (-0.06,0.11) |
| Blade atrial septostomy | 0.05 | -- | -- | -- | -- | -- | -- |
| No qualifying codes | 0.03 | -- | -- | 0.18 (-0.23,0.58) | -- | -- | -- |
| PFO transluminal | 0.03 | 0.06 (-0.12,0.23) | -- | -- | -- | 0 (-0.08,0.08) | 0.01 (-0.06,0.08) |
| AVSD Fallot b | 0.02 | -- | -- | -- | -- | -- | -- |
| CCTGA repair b | 0.01 | -- | 0.03 (-0.06,0.12) | -- | 0.22 (-0.24,0.69) | -- | 0.06 (-0.02,0.14) |
| Atrial switch | 0.01 | -- | -- | -- | -- | 0.02 (-0.05,0.1) | 0.01 (-0.04,0.06) |
| TAPVC shunt | 0.01 | -- | -- | -- | -- | 0.03 (-0.04,0.1) | -- |
| Arrhythmia surgical | 0.01 | -- | -- | 0.19 (-0.21,0.6) | -- | 0.06 (-0.04,0.17) | -- |

*--Procedure not performed in the specific period; AP - Aortopulmonary; AV – Atrio Ventricular; AVSD – Atrio Ventricular Septal Defect; CCTGA-Congenitally Corrected Transposition of Great Arteries; CRT BV- Cardiac Resynchronisation Therapy Biventricular pacemaker; DORV- Double Outlet Right Ventricle; IAA – Interrupted Aortic Ach Repair; ICD – Implantable Cardioverter & Defibrillator; MAPCA – Major Aortopulmonary Collateral Arteries; PA – Pulmonary Artery; PAPVC – Partial Anomalous Pulmonary Venous Connection; PDA – Patent Ductus Arteriosus; PFO – Patent Foramen Ovale; RV PA – Right Ventricular to Pulmonary Artery conduit; RVOT – Right Ventricular Outflow Tract; RVOTO – Right Ventricular Outflow Tract Obstruction; TAPVC – Total Anomalous Pulmonary Venous Connection; TGA – Transposition of Great Arteries; VA – Ventricular Arterial discordance; VSD – Ventricular Septal Defect*

### Supplementary Figure S1: Difference in the mean percentage (95%CI) of elective, urgent, and emergency or life-saving procedures during each pandemic period compared to pre-pandemic

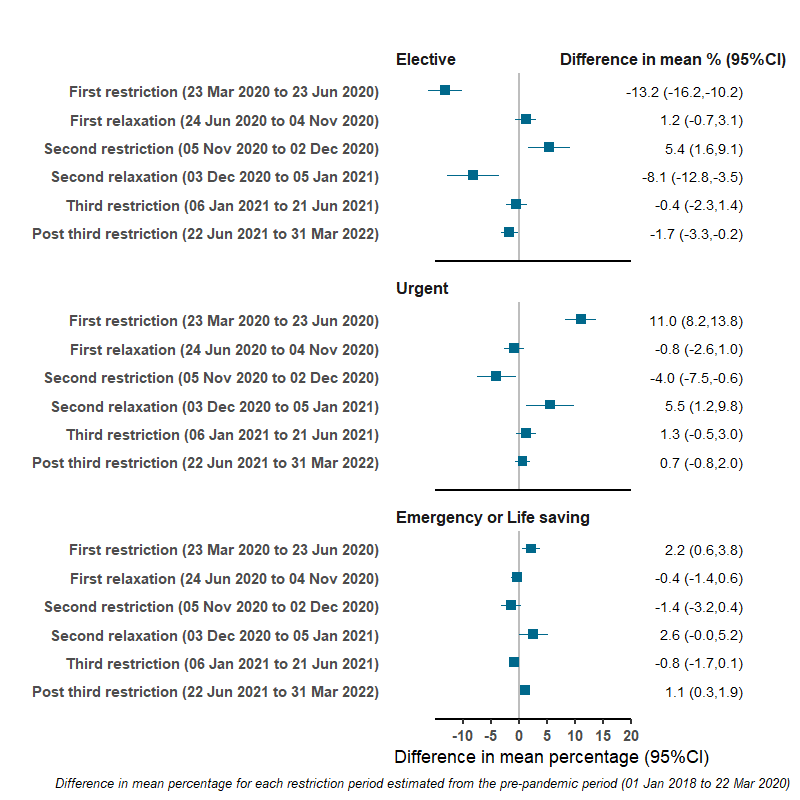

*Results show the difference in mean percentage (95% CI) of elective, urgent, and emergency/life-saving pediatric procedures during each period compared to the pre-pandemic period. We combined emergency and life-saving procedures.* *We combined emergency and life-saving procedures into a single category because of low numbers*

### Supplementary Figure S2: Odds ratios of mortality within 30-days of cardiac surgery comparing pandemic period to the pre-pandemic period showing the impact of adjustment with age and two different approaches to adjusting for case mix

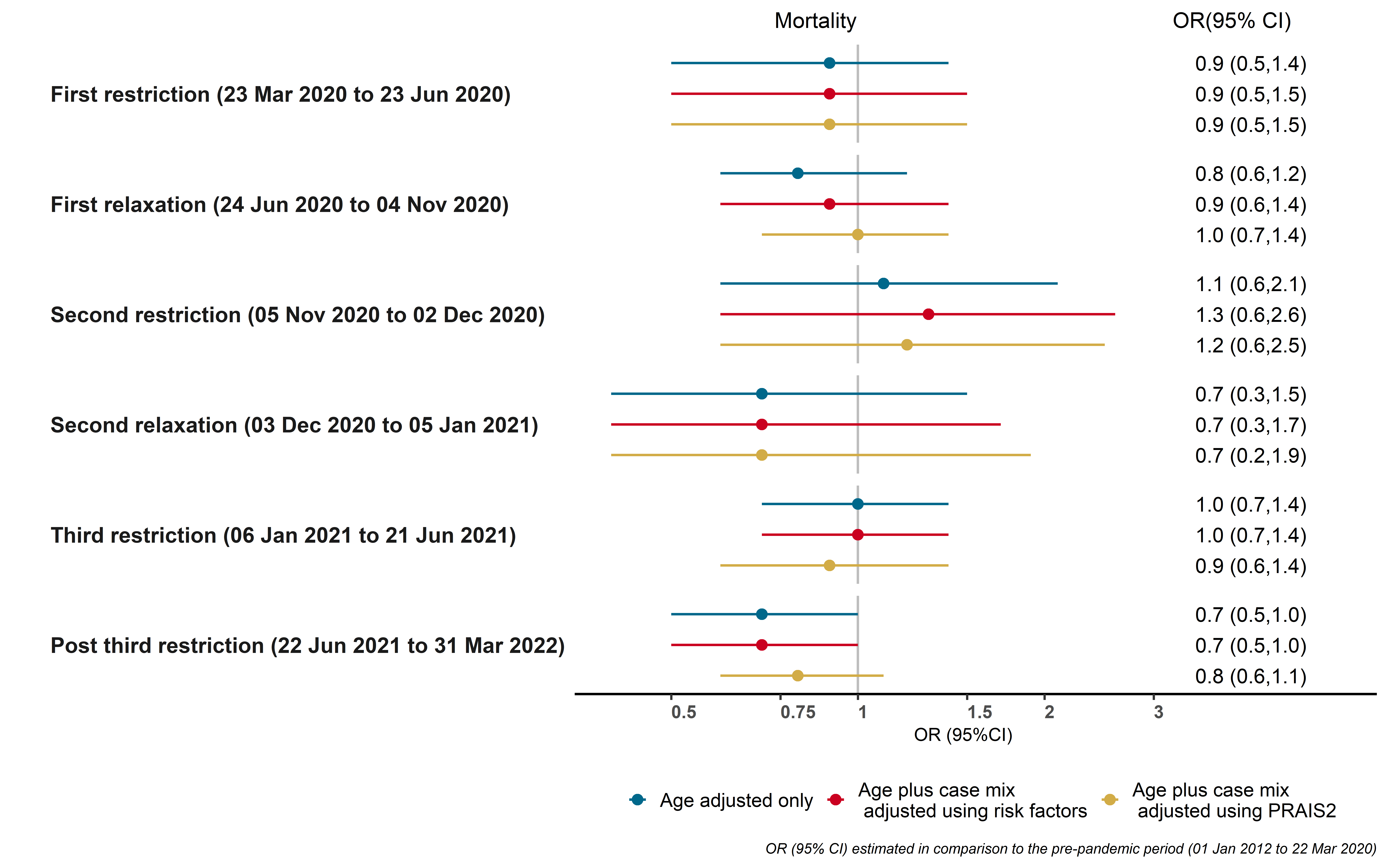

*Results show the age adjusted, age plus case mix adjusted using risk factors included in PRAIS2 and age plus case-mix adjusted using the derived PRAIS2 score odds of mortality within 30-days of cardiac surgery between different periods of the pandemic compared with the pre-pandemic periods*

### Supplementary Figure S3: Odds ratios of urgency, emergency, or life-saving pediatric congenital heart disease procedure during pandemic among different age groups

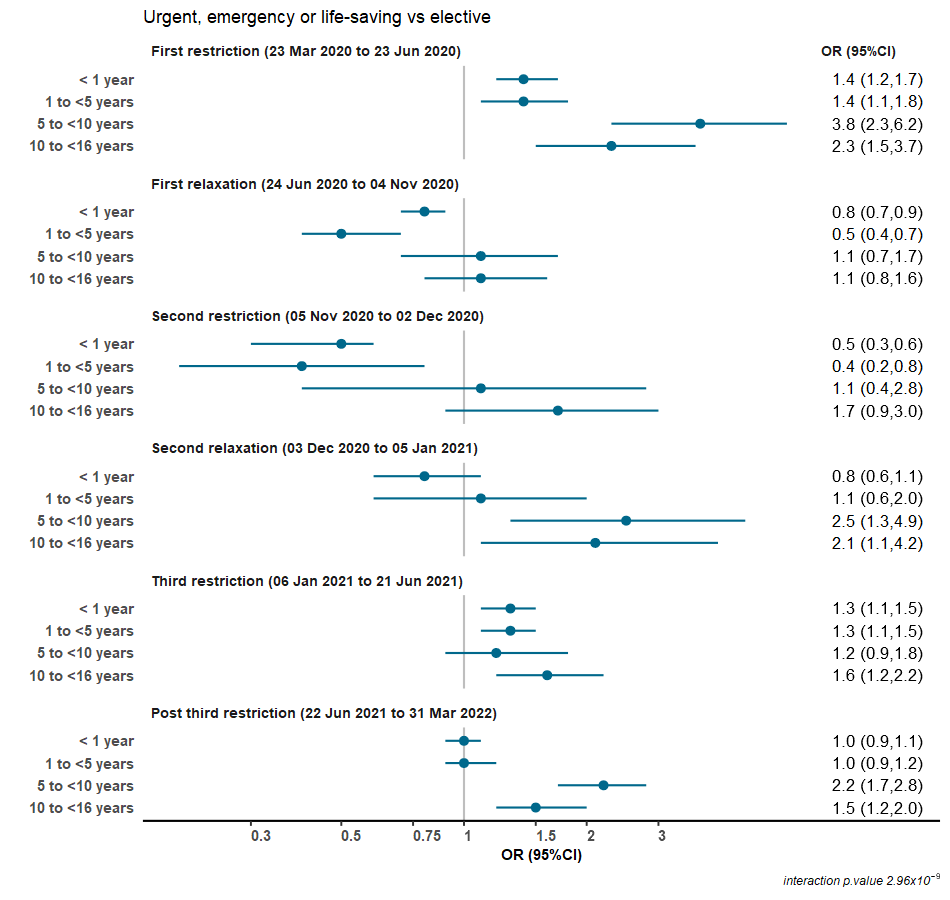

*Results show the odds of urgent/emergency/life-saving vs elective procedure among children who had congenital heart disease procedures during different periods of restriction or relaxation compared with the pre-pandemic periods among different age groups. Interaction p-value for differences = 2.96x10^-9^ We combined urgent, emergency, and life-saving procedures into a single category.*

### Supplementary Figure S4: Odds ratios of post procedure complication following pediatric congenital heart disease procedure during pandemic among different age groups.

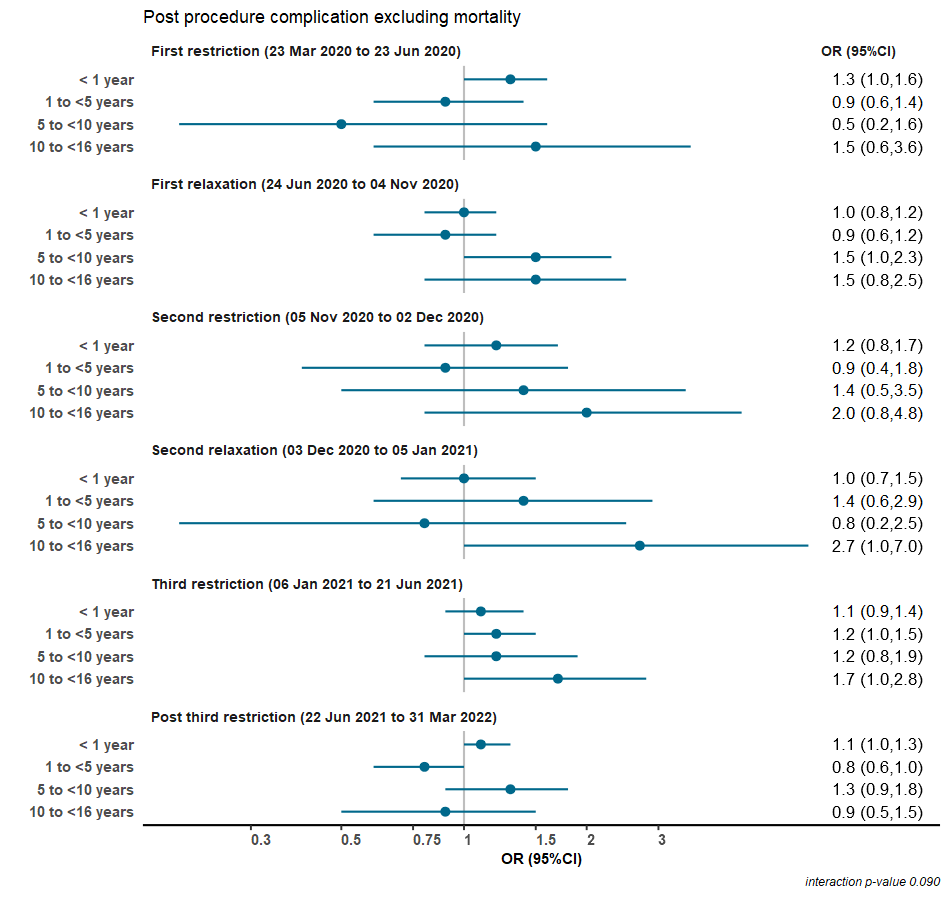

*Results show the odds of post-procedure complications (yes vs no) among children who had congenital heart disease procedures during different periods of restriction or relaxation compared with the pre-pandemic periods among different age groups. Interaction p-value for differences =0.090*

### Supplementary Figure S5: Odds ratios of mortality within 30 days of a pediatric congenital heart disease procedure during pandemic among different age groups

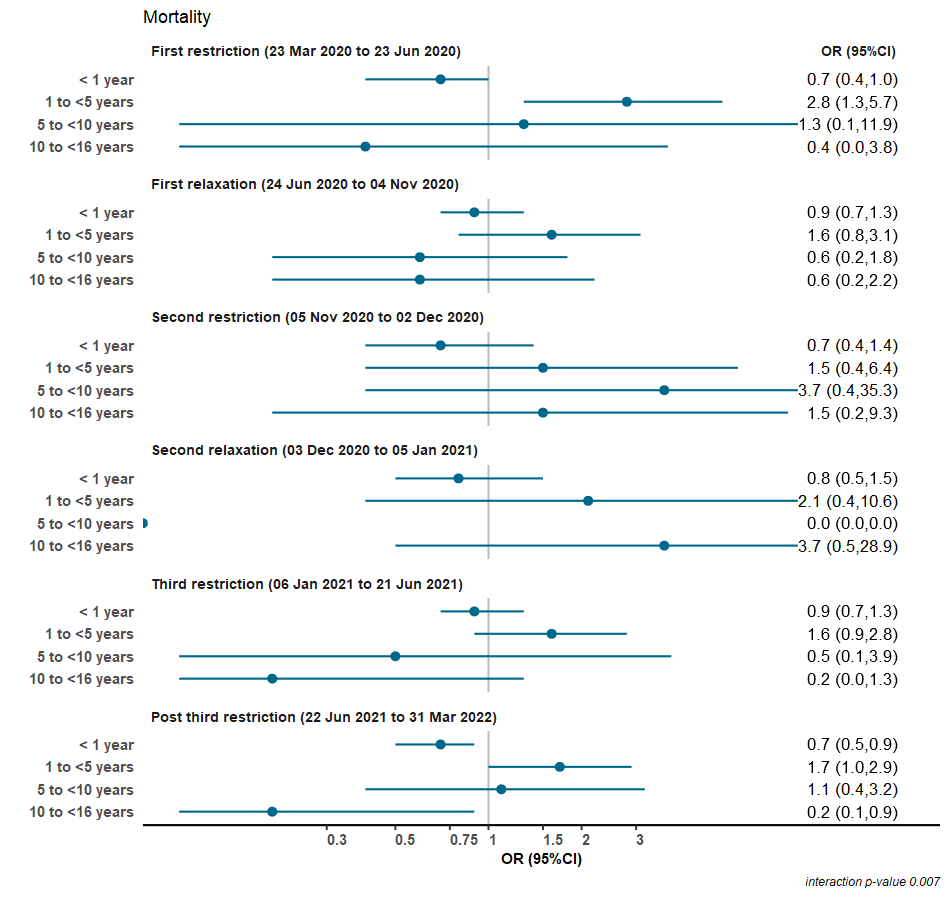
 *Results show the odds of mortality within 30-days of a procedure among children who had congenital heart disease procedures during different periods of restriction or relaxation compared with the pre-pandemic periods among different age groups. Results are adjusted for case mix. Interaction p-value for differences = 0.007*

### Supplementary Figure S6: Odds of urgency of a pediatric congenital heart disease procedure during pandemic among different ethnic groups

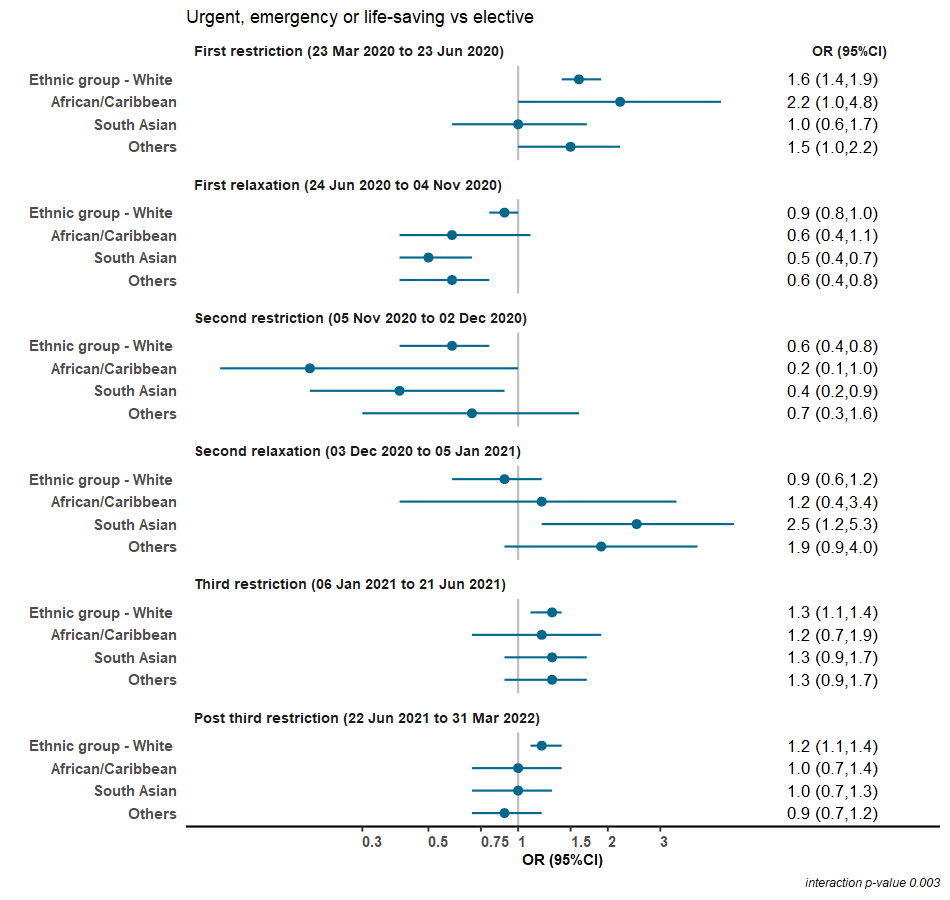

*Results show the odds of urgent/emergency/life-saving procedure among children who had congenital heart disease procedures during different periods of restriction or relaxation compared with the pre-pandemic periods among different ethnic group. Interaction p-value for differences = 0.003. We combined urgent, emergency, and life-saving procedures into a single category.*

### Supplementary Figure S7: Odds ratios of post-procedure complication of a pediatric congenital heart disease procedure during pandemic among different ethnic groups

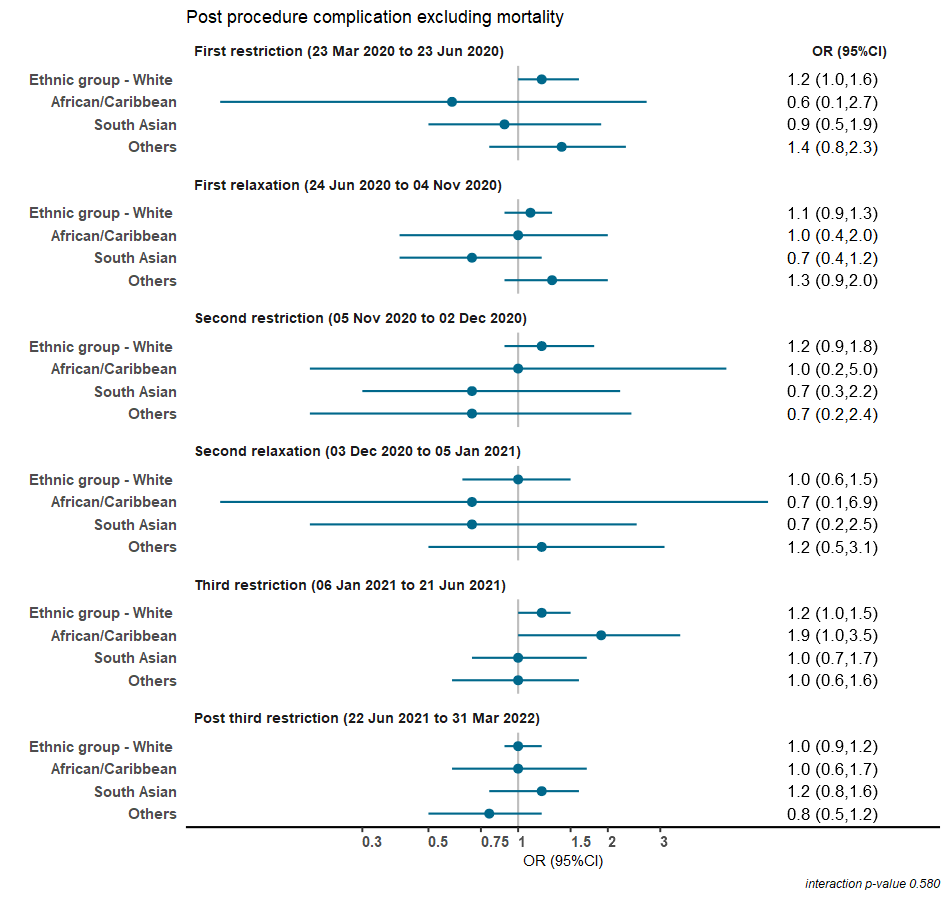

*Results show the odds of post-procedure complications (yes vs no) among children who had congenital heart disease procedures during different periods of restriction or relaxation compared with the pre-pandemic periods among different ethnic groups. Interaction p-value for differences = 0.580*

### Supplementary Figure S8: Odds ratios of mortality within 30 days of a pediatric congenital heart disease procedure during pandemic among different ethnic groups

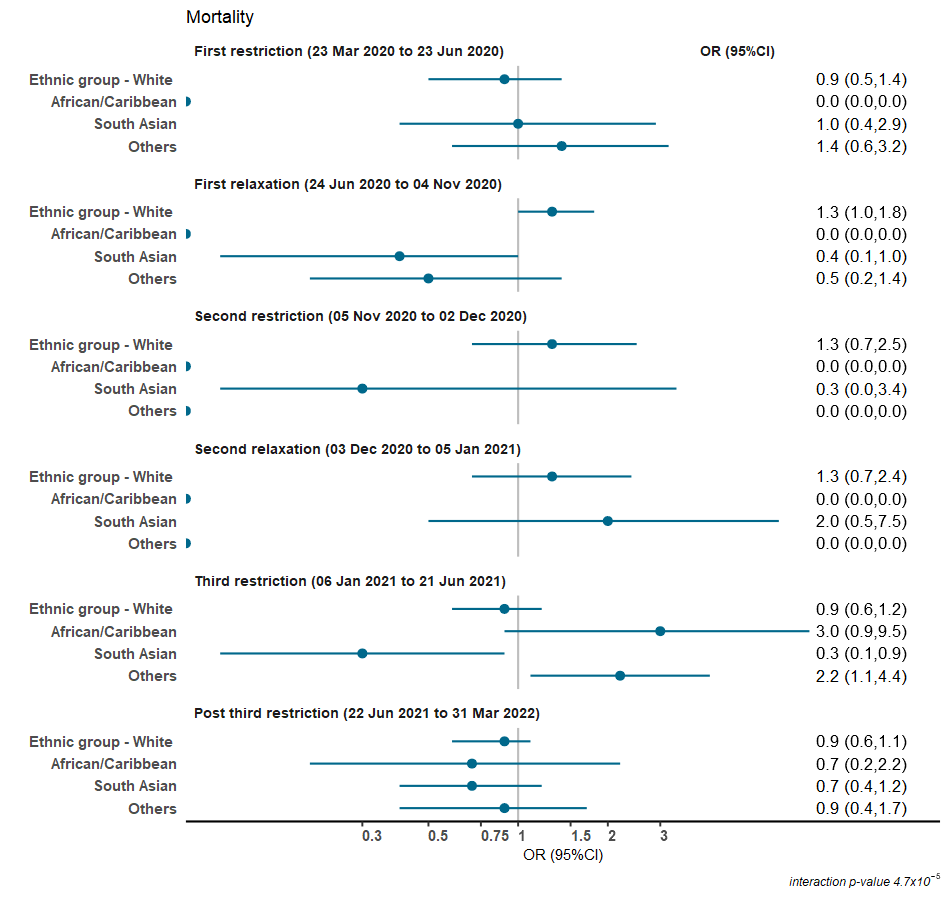

*Results show the odds of mortality within 30-days of a procedure among children who had congenital heart disease procedures during different periods of restriction or relaxation compared with the pre-pandemic periods among different ethnic groups. Interaction p-value for differences = 4.7x10^-5^*

### Supplementary Figure S9: Odds of urgency of a pediatric congenital heart disease procedure during pandemic among different levels of area deprivation

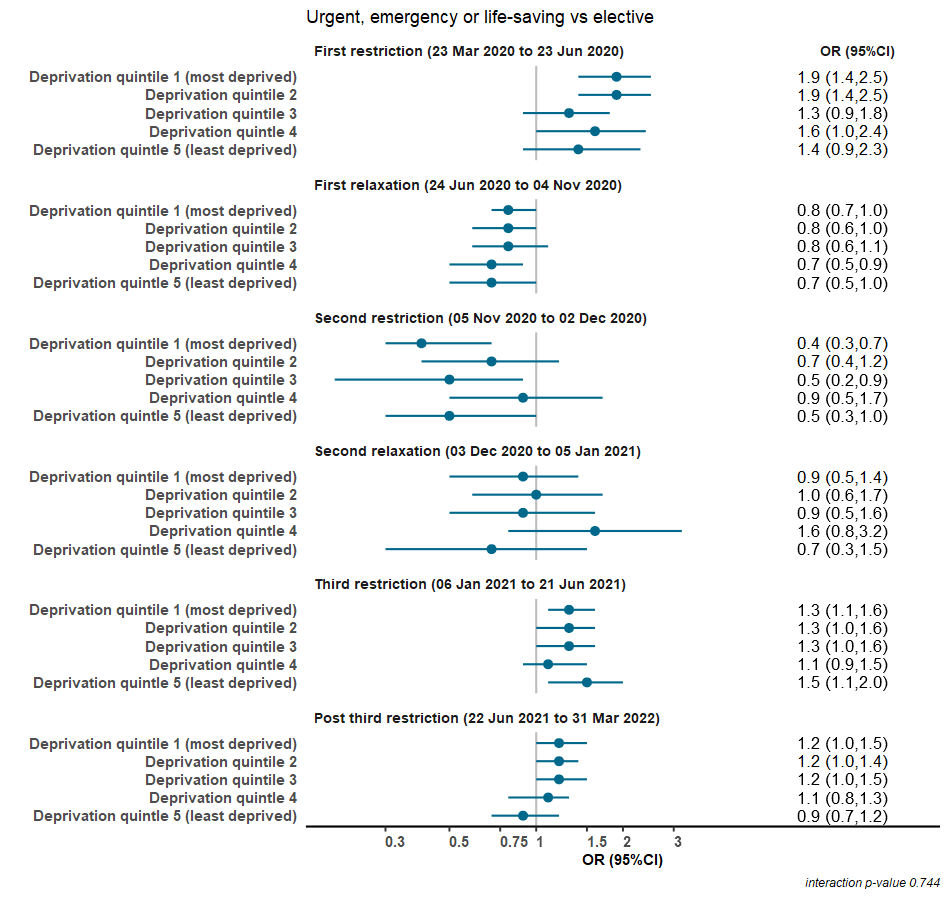

*Results show the odds of urgent/emergency/life-saving procedure among children who had congenital heart disease procedures during different periods of restriction or relaxation compared with the pre-pandemic periods among different levels of deprivation*. *Interaction p-value for differences = 0.744*

### Supplementary Figure S10: Odds of post procedure complication of a pediatric congenital heart disease procedure during pandemic among different levels of area deprivation

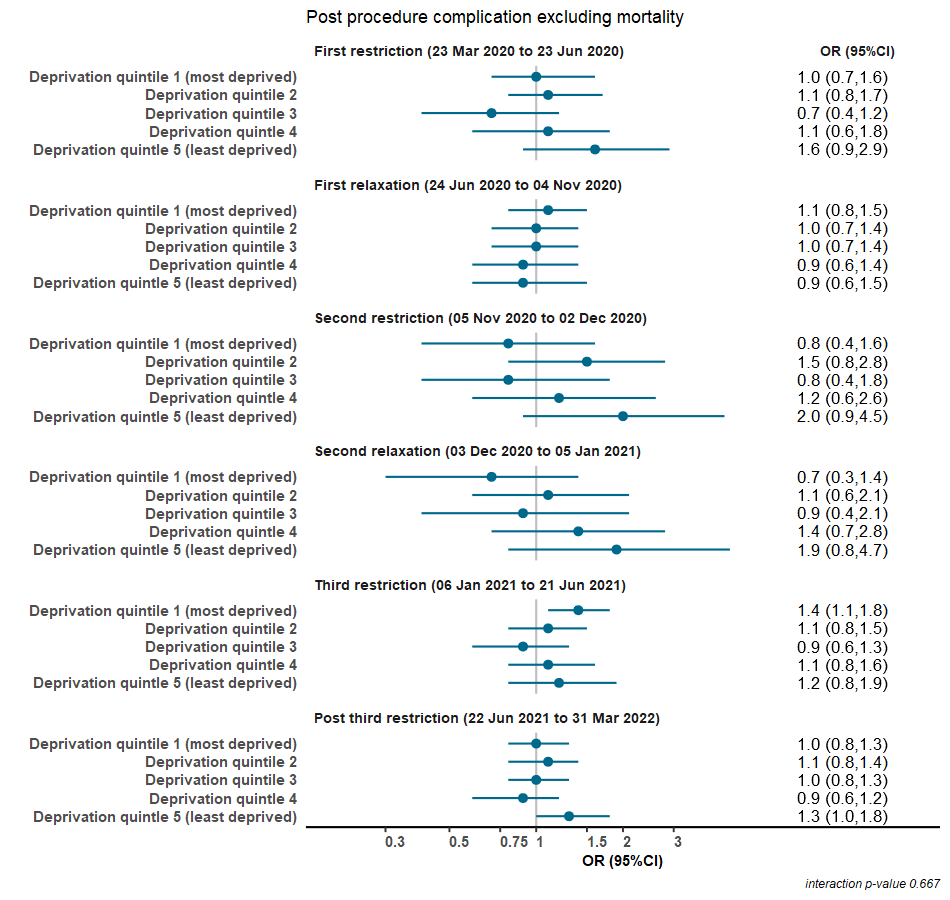

*Results show the odds of post- procedure complications among children who had congenital heart disease procedures during different periods of restriction or relaxation compared with the pre-pandemic periods among different levels of deprivation. Interaction p-value for differences = 0.667*

### Supplementary Figure S11: Odds of mortality within 30 days of a pediatric congenital heart disease procedure during pandemic among different levels of area deprivation

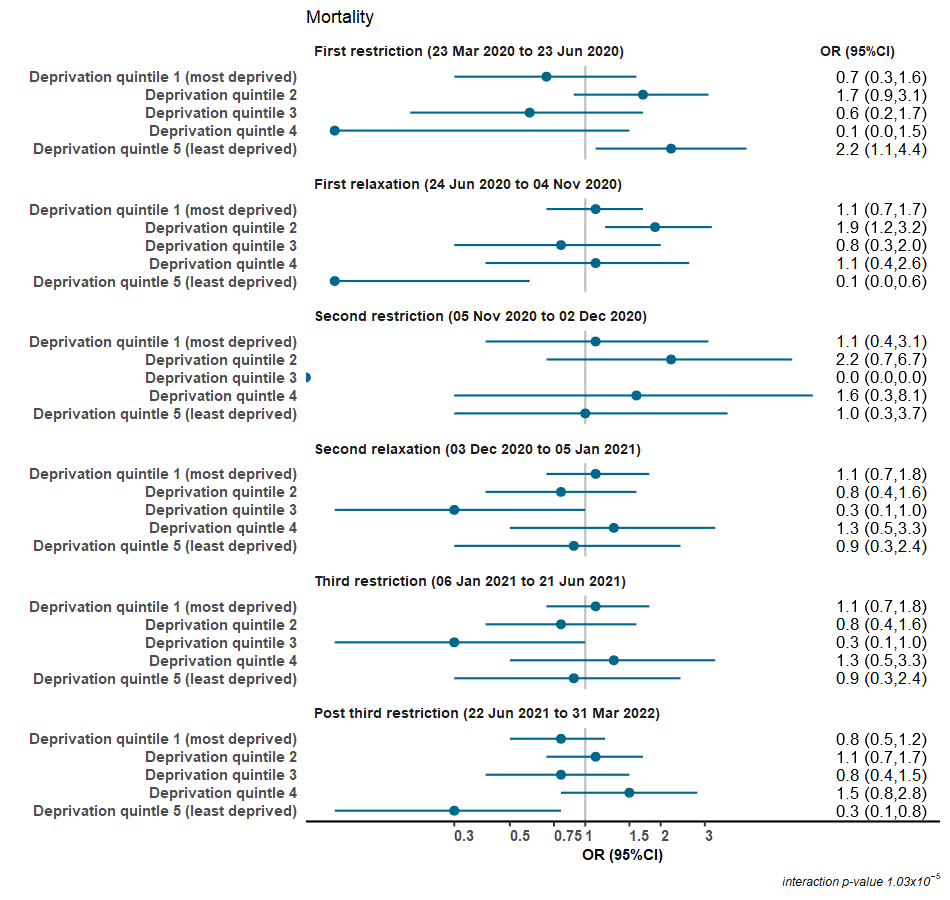

*Results show the odds of mortality within 30-days of a procedure among children who had congenital heart disease procedures during different periods of restriction or relaxation compared with the pre-pandemic periods among different levels of deprivation****.*** *Interaction p-value for differences = 1.03x10^-5^*
